# False-Positive Calls of Reimplemented Published Analyses for Covert Command-Following on Electroencephalograms Recorded Without Study Commands

**DOI:** 10.64898/2026.11.03.26367200

**Authors:** Alon Gorenshtein, Yosef Adiniaev, Yiftach Barash, Eyal Klang, Oved Daniel

## Abstract

**Objective:** To measure how often published electroencephalographic analyses for covert command-following call patients positive without commands, and their detection of genuine responses.

**Methods:** This cross-sectional study applied 11 reimplemented published analyses to electroencephalograms without commands: 974 clinical electroencephalography patients and, with rules fixed, 730 new patients; 38 with prolonged disorders of consciousness; and 544 comatose patients after cardiac arrest. Outcomes: false calls (share called positive) against a 5% benchmark, and genuine-response detection.

**Results:** Three of 10 command-following analyses called significantly more than 5% of clinical electroencephalography patients positive, replicated in new patients: Goldfine 2011, a spectral test, 23.3% (95% CI, 20.2%-26.6%), above its stated 5%; du Bois 2026, a criterion for entry to brain-computer interface training, 22.2% (95% CI, 19.2%-25.5%); and Guger 2018, a P300 accuracy threshold, 13.8% (95% CI, 11.5%-16.3%), near its approximate 12% chance rate (24.5% over 2 runs). The other 7 called 0.0% to 6.9%. After cardiac arrest: 11.5% to 27.3%. All 3 detected genuine responses. Identified contributors: a variance ignoring within-command dependence (Goldfine 2011), selection over configurations and times (du Bois 2026) and a permissive threshold (Guger 2018). Goldfine 2011’s command-level correction, fixed before testing, called 0.8% of new patients and 2.4% after arrest, detecting 93.4% of stroke imagery-versus-rest sessions but few between imagery types.

**Interpretation:** Reimplemented published analyses called up to 27% of patients positive without commands, more with repeated runs. A positive result needs its complete call rule’s false-call rate and detection at that rate; imagery-versus-rest detection may reflect task engagement, not imagery.

## Introduction

An unresponsive patient is asked during EEG to imagine moving a hand, and the analysis returns a positive result. Between 15% and 25% of patients without a behavioral response show such responses, termed cognitive motor dissociation (CMD),^1,2^ which has been associated with earlier and more complete recovery.^1,3^ European and US guidelines both make weak recommendations: the European to consider task-based EEG or functional MRI in multimodal assessment, the US to allow such tests when bedside findings remain ambiguous.^4,5^ A positive result can shape prognostic counseling and decisions about life-sustaining treatment.^6^

Statistical significance alone does not make a positive result trustworthy. A widely cited motor imagery result depended on treating trials within a block as independent,^7^ the binomial test of cross-validated accuracy overstates significance,^8^ and publications often leave such choices unstated.^1,9–11^ A "ghost response check", running the analysis on recordings in which no task was presented, has been proposed to estimate how often current methods give false-positive results.^6^ To our knowledge, it has not been carried out across published analyses or in patients.

In this cross-sectional study, we ran published EEG analyses for covert command-following on recordings from patients who received no study command. We measured how often each called patients positive, alone and over repeated runs, in clinical EEG and in coma after cardiac arrest, and whether randomizing trial order prevented it. We then tested the analyses with excess calls on genuine responses and sham labels, identified analytical contributors to the excess and measured detection at a patient-level false-call rate targeted within each design.

## Methods

### Study Design and Data

This cross-sectional study used de-identified EEG recorded without study commands; reporting follows STARD 2015 and STROBE. Clinical EEG came from the SpikeNet 2.0 corpus of the Brain Data Science Platform (BDSP), routine, epilepsy-monitoring and long-term recordings from 3 institutions.^12,13^ A development set of 974 patients was analyzed first; after every analysis rule had been fixed, 730 further patients never analyzed in this study were drawn from the same corpus as a replication set (Supplementary Methods 15). A validated EEG foundation model assigned each window a background.^14,15^ Overnight polysomnography of patients with prolonged disorders of consciousness (DoC), diagnosed with the Coma Recovery Scale-Revised (CRS-R), came from Mendeley Data^16–18^ and was analyzed on its 6 EEG channels; post-arrest EEG came from I-CARE, as one 30-minute stretch nearest 24 hours after return of circulation (Table 1).^19^ Detection was measured on healthy participants’ motor imagery^20,21^ and auditory counting,^22^ and sensory responses on passive oddball recordings^23,24^ (Table S1 in Supplementary Data S1).

**Table 1.** Cohorts.

| **Characteristic** | **Clinical EEG (n = 974)** | **Prolonged DoC (n = 38)** | **Post-arrest coma, complete data (n = 490)** |
|---|---|---|---|
| Recording | Routine, epilepsy-monitoring or long-term; 19 channels | Overnight polysomnography; 6 EEG channels | ICU monitoring; 19 channels |
| Analyzed duration, median (IQR) | 30 min (30-53) | 6.0 h (6.0-6.0) | 30 min (fixed) |
| First-window background, %<sup>a</sup> | Awake 27-34; sleep 42-47; ICU-typical ≤4 | Not available | Continuous 27; discontinuous 22; burst suppression 36; suppressed 16 |
| Diagnosis, No. | Not available | VS/UWS 28; MCS 10 | Coma after cardiac arrest 490 |
| Age, median (IQR), y<sup>a</sup> | Not available | 42 (37-57) | 62 (52-72) |
| Women, No. (%)<sup>a</sup> | Not available | 14 (47) | 148 (30) |
| Etiology or arrest, No.<sup>a</sup> | Not available | Traumatic 14; anoxic 10; vascular 5; inflammatory 1 | Out-of-hospital arrest 382; shockable rhythm 258 |
| Time since injury, median (IQR)<sup>a</sup> | Not applicable | 4 mo (3-5) | 24 h (24-24) |
| CRS-R total, median (IQR)<sup>a</sup> | Not applicable | 6.5 (5-8) | Not applicable |

Abbreviations: CRS-R, Coma Recovery Scale-Revised; DoC, disorders of consciousness; MCS, minimally conscious state; VS/UWS, vegetative state/unresponsive wakefulness syndrome. <sup>a</sup> DoC: the 30 patients with linked clinical data. Post-arrest coma: background is the American Clinical Neurophysiology Society continuity grade; 1 patient had no recorded sex. Race and ethnicity were not available in any dataset.

### Published Analyses

We reimplemented 11 published analyses from their publications and public code, 10 for command-following and 1 for covert processing, which is reported separately (Table 2; Tables S2 and S4). Claassen 2019 reproduces the authors’ public code. The Goldfine 2011, Chennu 2013 and Sokoliuk 2021 analyses reproduce the core statistic of a public implementation, with manual independent component analysis and visual rejection replaced by automated rules; the Goldfine 2011 statistic agreed with a translation of the public toolbox’s routine to within 4 × 10^-15^ on 30 random inputs, and the toolbox’s variance changed none of its 79 calls on the 342 development recordings held locally. The others were reimplemented from published descriptions; for the mindBEAGLE-type analyses, the vendor’s software was not tested. The du Bois 2026 reimplementation departs from several stated choices, among them fewer spatial filters, a coarser time step and no peak refinement (Table S4). A second implementation of the du Bois 2026 and Guger 2018 analyses, written from the publication texts alone without access to ours, was run on the same development windows (Supplementary Methods 16).

**Table 2.** Published Analyses, Their Call Rules and the Reimplementations.

| **Analysis (reimplementation)<sup>a</sup>** | **Positive call used for; original population** | **Call rule** | **Stated or implied error rate** |
|---|---|---|---|
| Claassen 2019, support vector machine<sup>1</sup> (authors' code) | Command-following; acute brain injury | AUC above 0.5, *p* < 0.05 | 5% |
| Goldfine 2011, spectral test<sup>9,25</sup> (public toolbox statistic) | Command-following; severe brain injury | Spectral difference *p* ≤ 0.05 (jackknife), false discovery rate, contiguity | 5% |
| Cruse 2011, motor imagery classifier<sup>10</sup> (description) | Command-following; vegetative state | Accuracy above the 99% binomial interval | 0.5% |
| Cruse 2012, motor imagery classifier<sup>26</sup> (description) | Command-following; vegetative state | Either of 2 permutation tests *p* < 0.025 | 5% |
| Edlow 2017, motor command classifier<sup>11</sup> (description) | Command-following; acute brain injury | Permutation *p* < 0.05 | 5% |
| Guger 2017, motor imagery, mindBEAGLE-type<sup>27-29</sup> (description) | Command-following; locked-in, DoC, intensive care | Accuracy above the 95% binomial limit | 2.5% |
| du Bois 2026, configuration search<sup>30</sup> (description; departures, Table S4) | Entry to brain-computer interface training; prolonged DoC, locked-in | Peak accuracy above 70% or paired t test *p* < 0.05 | 6% for one configuration<sup>b</sup> |
| Guger 2018, vibrotactile P300, mindBEAGLE-type<sup>31,32</sup> (description) | Discriminable response to commands; prolonged DoC | Accuracy above 23% | Not stated; about 12% by chance<sup>c</sup> |
| Gibson 2016, event-related potentials<sup>33</sup> (description) | Command-following; prolonged DoC | Cluster permutation *p* < 0.05 | 5% |
| Chennu 2013, event-related potentials<sup>34</sup> (authors' code, core statistic) | Command-following; prolonged DoC | All 3 cluster tests *p* ≤ 0.05<sup>d</sup> | 5% |

Abbreviations: AUC, area under the receiver operating characteristic curve; DoC, disorders of consciousness. <sup>a</sup> Authors' code: the authors' public code; public toolbox statistic: the core statistic of a public toolbox, checked against it on test inputs, not the full pipeline (Supplementary Methods 2); description: reimplemented from the published description. Analyses are named by the first author and year of the publication reimplemented; mindBEAGLE-type analyses reimplement published descriptions of the vendor's methods, not its software. The covert-processing analysis<sup>35</sup> is reported in Supplementary Results 2. <sup>b</sup> A simplified reference: 5% for the t test plus at most 1% for the 70% threshold, which the publication places above the 99% binomial bound for 90 trials; the reference assumes independent trials, one configuration and one time point, and does not cover the search over configurations and peak times (Table S19). <sup>c</sup> More than 23% of 10 split-half predictions correct, each with a 1 in 8 chance, if the splits were independent (Table S19). <sup>d</sup> The target-word P3b against baseline, its temporal cluster test and the explicit-against-implicit contrast, the criteria the publication's positive patient met.

### Running the Analyses Without Commands

Each analysis’s own schedule of commands or stimuli was placed on a seeded window of each recording, so every positive call was false with respect to commands. Schedules longer than 30 minutes were also run later on longer windows where recordings held them, and Claassen 2019 on the 6 DoC channels (Supplementary Methods 18). Per-patient rules used each patient’s first windows at the published number of assessments: any of 2 recordings for Claassen 2019 (bounded with other patients’ windows; Table S9B),^1,3^ either of 2 sessions for du Bois 2026, either of 2 runs for Guger 2018 (with up to 7 runs reported descriptively) and 1 for the others.

### Choices the Publications Left Open

Where a publication left a choice open, such as the cross-validation unit, each option consistent with it was run. The primary result used the consistent option with the fewest positive calls, so primary rates are the lowest among the consistent options run; the option was chosen on each half of the patients and tested on the other; full-sample rates are reported (Table S17). This rule and corrections to 2 reimplementations were set after initial rates had been seen; results under every consistent option, the originally planned option and the chronology are in Supplementary Methods 3, 4 and 12. The rules were then fixed and applied unchanged to the replication set.

### Randomized Trial Order and Detection

Each analysis was also run with trial order randomized trial by trial and tested by randomization at that unit (200 draws). Detection was measured on simulated responses added to the no-command EEG, against the published analysis size-corrected to 5%, on healthy participants’ sessions analyzed with true and shuffled labels (Supplementary Methods 7-9); Guger 2017 was also run in 47 instructed patients with acute stroke (Supplementary Methods 16).

### Genuine-Task Controls and Calibration

The 3 analyses with excess calls were then run on open recordings with genuine tasks; code, sham labels and pass criteria were committed before these recordings were analyzed (Supplementary Methods 17). The Guger 2018 rule was run on a tactile task in which 19 blind or sighted volunteers counted touches on 1 of 8 fingertips in 2 sessions each,^36^ with an unattended finger as the sham label. The Goldfine 2011 and du Bois 2026 rules were run on the recorded design of a stroke motor imagery dataset, 61 sessions of 23 patients alternating rest with imagery of the affected hand or of a robotic sixth finger;^37^ Goldfine 2011 compared imagery with rest and du Bois 2026 the 2 imagery types, with sham labels from rest periods only (Goldfine 2011) or redrawn at random (du Bois 2026), 20 times per session. On 200 development windows, a second du Bois 2026 implementation following the publication’s stated choices was varied one choice at a time, and its permutation evaluation was run literally or with the configuration search repeated inside each permutation. The Goldfine 2011 jackknife was recomputed with whole commands, rather than single tapered estimates, as the deleted unit; this variance was then fixed in code and applied unchanged to the replication and post-arrest windows, and to left-against right-hand imagery at the same post-cue time in 47 patients with acute stroke.^38^ Finally, each statistic was thresholded on null recordings of its own design (command-free clinical EEG or sham labels), cross-fitted by patient, targeting a 5% chance of any false call over k assessments (1, 2, 5 or 10) under independence, and applied to genuine responses; where patients had repeated null runs, the patient-level rate was also measured directly.

### Statistical Analysis

The primary outcome was the share of patients called positive (the false-call rate), with exact Clopper-Pearson 95% CIs. In the replication set, the 3 analyses above 5% in development were tested against 5% by one-sided exact binomial tests with Holm adjustment; no option was selected on these patients. The other analyses were tested for rates below 10%. Five percent was a common benchmark, not every rule’s stated error rate; each rate was also compared with the rate its rule states or implies and with the same rule’s rate on white noise (Table 2; Table S19).

Prolonged-DoC and post-arrest rates were estimated, not tested. Only the replication tests and the frozen test of the command-level Goldfine 2011 variance (one-sided exact binomial against 5%, Holm over the replication and post-arrest sets) were confirmatory; all other analyses were exploratory, including comparisons with published cohorts that delivered commands (Supplementary Methods 16).

Real-task rates carry participant-or patient-cluster bootstrap CIs. Analyses were run in October 2026 with Python 3.9.6 (real-task analyses, Python 3.12); code is at https://github.com/Alon-Gorenshtein/False-Positive-Calls-Covert-Command-Following-EEG.

### Ethics

BDSP recordings were collected under a Mass General Brigham Institutional Review Board protocol (2012P001929) with a waiver of consent and used under the platform’s data use agreement; the other datasets are public. This secondary analysis of de-identified data required no further review or consent.

## Results

### Cohorts

The development set comprised 974 clinical EEG patients and the replication set 730 patients; the DoC set comprised 38 patients and the post-arrest set 544 comatose patients with a usable window, 490 of them with complete clinical data, of whom 185 (37.8%) had a good outcome (Table 1; Tables S6 and S16).

### Positive Calls Without Study Commands

#### Without any study command, 3 of 10 analyses called 14% to 23% of clinical EEG patients positive

Goldfine 2011 called 164 of 703 patients (23.3%; 95% CI, 20.2%-26.6%), du Bois 2026 157 of 707 (22.2%; 95% CI, 19.2%-25.5%) and Guger 2018 117 of 850 (13.8%; 95% CI, 11.5%-16.3%) (Fig 1A); the other 7 called 0.0% to 6.9%. These positive calls differ in kind: command-following (Goldfine 2011), entry to brain-computer interface training (du Bois 2026) and a discriminable response (Guger 2018) (Table 2). In the replication set, the fixed rules called 22.9% (95% CI, 19.8%-26.1%; Goldfine 2011), 24.8% (95% CI, 21.7%-28.1%; du Bois 2026) and 12.5% (95% CI, 10.1%-15.3%; Guger 2018) (Holm-adjusted *p* < 0.001 for each), each inside its 95% prediction interval from development; the other 7 called 0.0% to 5.2%, Cruse 2011, Cruse 2012 and Chennu 2013 on longer windows (Table S39). Claassen 2019’s 3.4% lay below the 95% prediction interval around its development rate of 6.9% (4.0%-9.8%; Table S17). The Goldfine 2011 rate exceeded the 5% its rule states and the du Bois 2026 rate a simplified 6% reference, and the Guger 2018 rate was close to its threshold’s chance rate, about 12% (15.2% on white noise) (Table S19). When a du Bois 2026 call in our implementation also required the label-permutation test that its publication reports as an evaluation against chance, the rate fell from 22.2% to 2.3% on clinical EEG, from 24.8% to 1.8% in the replication set and from 27.3% to 1.0% in post-arrest coma (Table S18). Depending on the implementation, 1 to 4 of 10 analyses called significantly more than 5% (Tables S7 and S21). A second implementation written from the same publication texts disagreed with ours, calling 96.0% vs 26.5% (du Bois 2026; κ, 0.02) and 17.4% vs 8.4% (Guger 2018; κ, 0.26) of the same development windows, below the agreement bar of 0.6 (Table S24); for du Bois 2026, 1 unstated choice accounted for most of the difference in rates (below).

**Figure 1.**
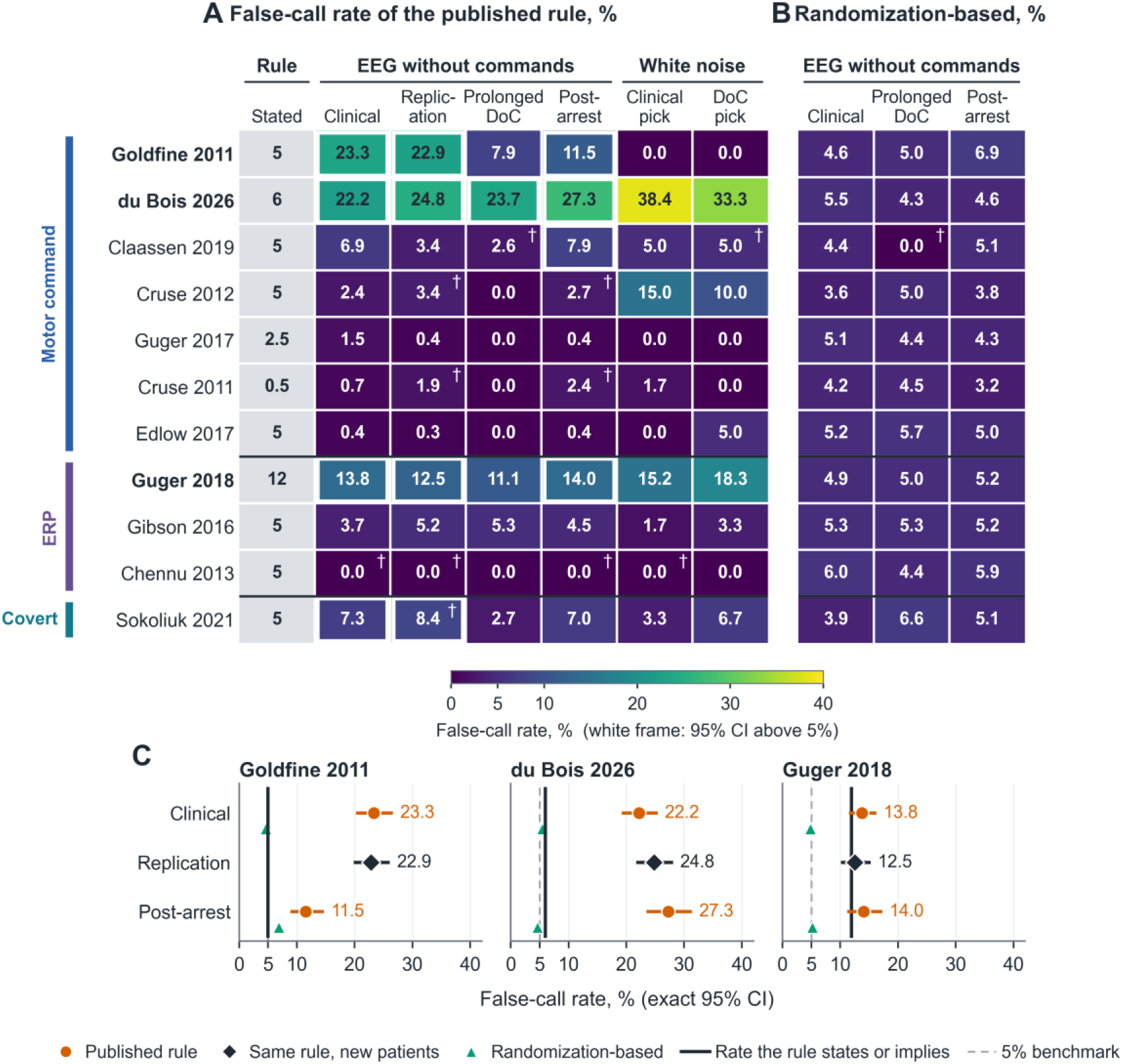
False-Call Rates of Reimplemented Published Analyses on EEG Recorded Without Study Commands. A, false-call rate (%) of the primary reading of each analysis, printed in each cell and coloured on one scale; rows grouped by method (motor command; event-related potential, ERP; covert processing); bold names, the 3 analyses whose excess was confirmed in the replication set. Columns: the rate each call rule states or implies (Guger 2018: about 12%, its threshold’s chance rate; du Bois 2026: 6% for one configuration and time point, not its full search, Table 2); clinical EEG (development set); the same readings, fixed, in the replication set; prolonged disorders of consciousness (DoC); coma 24 hours after cardiac arrest (all 544 patients with a usable 30-minute window); and white noise, for the reading chosen on clinical EEG and the reading chosen on DoC EEG (500 recordings for Goldfine 2011, du Bois 2026 and Guger 2018, 60 for the others; Cruse 2012, 500 further recordings, Table S11). White frame: exact 95% CI entirely above 5%, unadjusted, EEG columns only. Dagger: run later (Supplementary Methods 18; Table S39), the full-length schedules on longer windows of the patients whose recordings held them (Chennu 2013 in clinical EEG, 89 recordings; after arrest, 231 patients), Claassen 2019 on the 6 DoC channels and Sokoliuk 2021 in the replication set. B, randomization-based version of each analysis, a point estimate pooled over overlays (post-arrest, the 490 patients with complete data). C, the 3 analyses above 5% in the replication set, with exact 95% CIs: circles, primary reading; diamonds, replication set; triangles, randomization-based version; solid line, the stated or implied rate; dashed line, 5% benchmark. Originally planned readings are in Table S7B in Supplementary Data S2.

### Repeated Runs

#### Allowing more runs raised the share of patients called positive

Under the published two-run rule of Guger 2018, 206 of 841 patients (24.5%; 95% CI, 21.6%-27.5%) had at least 1 positive run (23.3%; 95% CI, 20.1%-26.8% in the replication set). In a fixed cohort of 639 patients with 7 runs, the share rose from 14.1% with 1 run to 62.9% with 7 (Fig 2; Table S9C).

**Figure 2.**
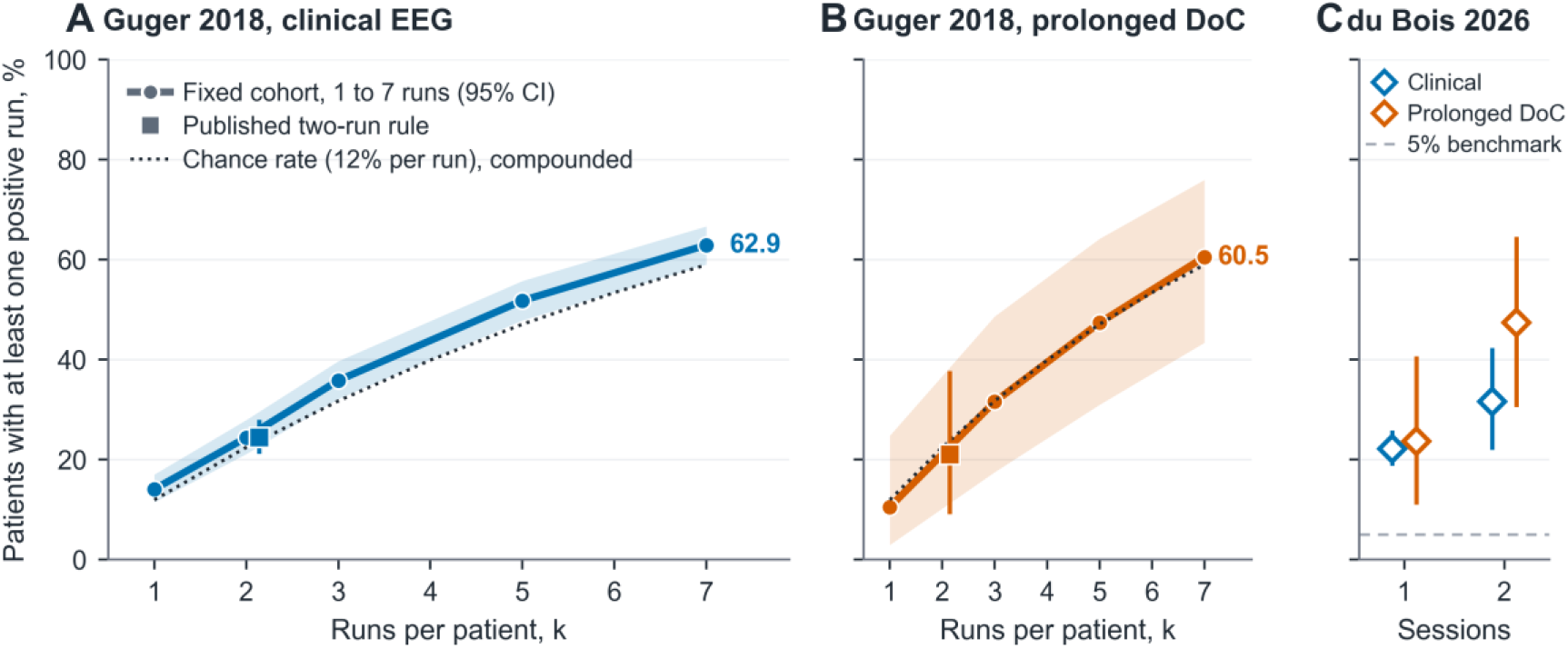
Patients Called Positive Over Repeated Runs. Patients with at least 1 positive run, with exact 95% CIs (bands and bars). A, the Guger 2018 rule on clinical EEG in a fixed cohort of 639 patients with 7 runs, by runs per patient (1 to 7, the published range; Table S9C); square, the published two-run rule in all 841 patients with 2 runs. B, the same rule in the 38 patients with prolonged DoC. Dotted line in A and B, the threshold’s 12% chance rate compounded over independent runs. C, du Bois 2026 single-session call (1) and two-session criterion (2) on clinical EEG (blue; for 2 sessions, the 95 patients with 2 eligible windows) and prolonged DoC EEG (orange); dashed line, 5% benchmark. Clinical EEG by patients with at least k runs and prolonged-DoC values, Table S9A in Supplementary Data S2.

### Coma After Cardiac Arrest

#### The same rules called 12% to 27% of comatose patients 24 hours after cardiac arrest

Applied unchanged from clinical EEG to the 544 patients with a usable window, du Bois 2026 called 140 of 513 (27.3%; 95% CI, 23.5%-31.4%), including 29 of 83 with a suppressed background, Guger 2018 75 of 534 (14.0%; 95% CI, 11.2%-17.3%) and Goldfine 2011 57 of 494 (11.5%; 95% CI, 8.9%-14.7%) (Fig 1C; Supplementary Results 10; Table S16H). On longer windows of 231 patients, Cruse 2011, Cruse 2012 and Chennu 2013 called 0.0% to 2.7% (Table S39). In the exploratory prolonged-DoC set, du Bois 2026 called 9 of 38 patients (23.7%; 95% CI, 11.4%-40.2%). Claassen 2019 on the 6 recorded channels called 1 (2.6%); on clinical EEG, the same 6 channels called 8.3% and all 19 called 6.9% (Table S39).

### Where the Positive Calls Came From

du Bois 2026 and Guger 2018 called 38.4% and 15.2% of 500 white-noise recordings; Goldfine 2011 called none, but its calls rose as the amplitude envelope of synthetic EEG fluctuated more (Tables S11D and S19). On sessions assembled from healthy volunteers’ motor imagery, the Goldfine 2011 and du Bois 2026 readings detected 51 and 41 of 51 participants but also called 14 and 15 of 51 label-shuffled sessions (Supplementary Figure S1). Guger 2017 called 9 of 47 patients with acute stroke instructed to imagine hand movements (19.1%; 95% CI, 9.1%-33.3%) and 3.2% of label-shuffled sessions (Table S28).

### Randomized Trial Order

Randomizing trial order gave false-call rates of 3.6% to 6.0% on clinical EEG and 3.2% to 6.9% in the 490 comatose patients with complete data (one Goldfine 2011 timing, 8.0%, Holm-significant). Detection of simulated responses was noninferior at 17 of 34 evaluation points and lower at every point for Goldfine 2011 (by 17.3-57.2 percentage points) and Edlow 2017 (by 8.4-19.9 points) (Table S14).

### Genuine Tasks and Sham Labels

**Each analysis with excess calls detected genuine responses, and each excess had identified contributors (Fig 3**).

**Figure 3.**
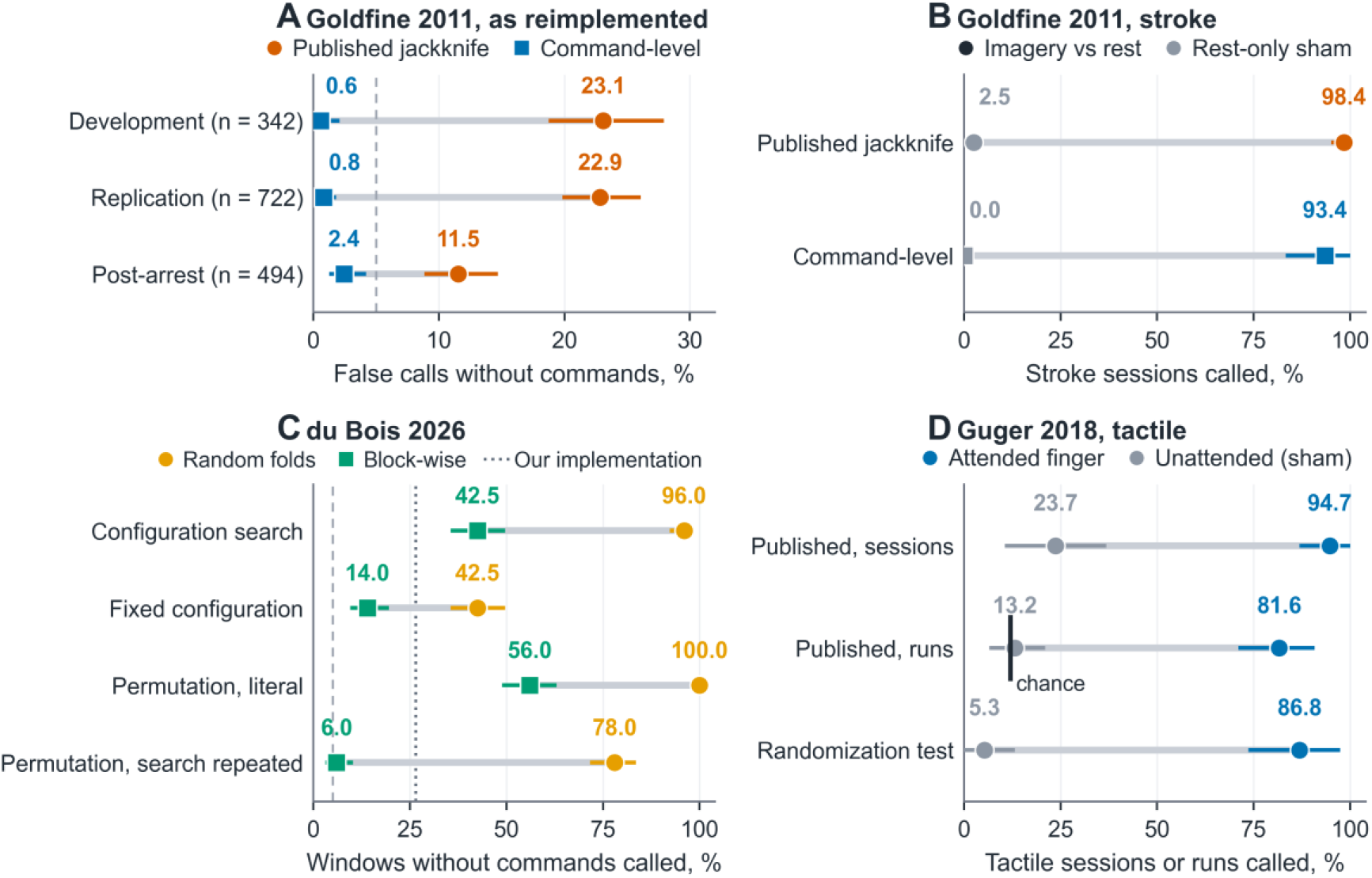
Genuine Tasks, Sham Labels and Contributors to Excess Calls. Each row joins two estimates with a grey line; the value is printed above each point. A, Goldfine 2011 false calls without commands with the published jackknife over tapered estimates, as reimplemented here (circles) and the command-level jackknife (squares): development windows held locally and, fixed in advance, the replication set and coma after cardiac arrest; exact 95% CIs; dashed line, 5% benchmark. B, Goldfine 2011 in stroke motor imagery sessions, imagery against rest with true labels (coloured by reading) and with rest-only sham labels (grey); patient-cluster 95% CIs. C, du Bois 2026 calls by the second implementation on 200 development windows without commands, by fold rule (circles, random folds; squares, block-wise folds), with the configuration search or a configuration fixed in advance, and under the publication’s permutation evaluation read literally or with the search repeated in each permutation; dotted line, our implementation; dashed line, 5% benchmark; exact 95% CIs. D, Guger 2018 on the tactile P300 task with the attended finger (blue) and an unattended finger (grey, sham) as the label: sessions called (either of 2 runs) by the published threshold, single runs by the published threshold, and sessions called by a randomization test; participant-cluster 95% CIs; black bar, the threshold’s chance rate (about 12%).

On the tactile task, the Guger 2018 rule called 36 of 38 sessions labeled with the attended finger (94.7%; participant-cluster 95% CI, 86.8%-100%), including all 19 volunteers, narrowly missing its preset bar (Table S32); with an unattended finger as the label, it called 13.2% of runs, near its chance rate, and under its 2-run rule 9 of 38 sessions (23.7%; 95% CI, 10.5%-36.8%).

Seventeen volunteers had more true than sham calls and none the reverse (*p* < 0.001); a randomization test called 86.8% and 5.3% of sessions (Tables S32 and S36). In the stroke sessions, Goldfine 2011 called 98.4% (95% CI, 95.1%-100%) with true labels and 2.5% (95% CI, 1.2%-4.4%) with rest-only sham labels. On the 342 development recordings held locally, deleting whole commands rather than single tapered estimates in its jackknife gave a variance a median 3.9 times that expected for independent estimates and lowered false calls from 79 (23.1%) to 2 (0.6%; 95% CI, 0.1%-2.1%); with this variance, the test called 93.4% of stroke sessions (4.9 points lower; 95% CI, 0 to 12.3; 3 discordant sessions, exact McNemar *p* = 0.25) and none with sham labels (Tables S31, S33 and S36). The second du Bois 2026 implementation called 96.0% (95% CI, 92.3%-98.3%) of 200 development windows with random folds and 42.5% with block-wise folds, which the publication does not state and our implementation uses; with a configuration fixed in advance, 14.0%. The publication’s permutation evaluation passed 100% and 56.0% of windows read literally, and 78.0% (71.6%-83.5%) and 6.0% (3.1%-10.2%; median *p*, 1.0) with the search repeated in each permutation (Fig 3C). In the stroke sessions, our du Bois 2026 implementation was positive in 86.9% of sessions with true labels and 46.7% with labels redrawn (Tables S30 and S33).

### The Corrected Goldfine 2011 Variance in New Patients

**Fixed before testing, the command-level variance kept false calls below 5% in patients not used to develop it (Fig 3A**; **Table S35).** It called 6 of 722 replication patients (0.8%; 95% CI, 0.3%-1.8%; published variance, 22.9%) and 12 of 494 comatose patients after cardiac arrest (2.4%; 1.3%-4.2%; published, 11.5%) (Holm-adjusted *p* < 0.001 and *p* = 0.003 for rates below 5%), with point estimates of at most 4.5% in any continuity grade (upper 95% limits up to 10.3%). Contrasting left-with right-hand imagery at the same post-cue time in 47 patients with acute stroke, the published variance called 10.6% of patients, no more than its 7.7% rate with pair-swapped labels, and the command-level variance 4.3% (null, 0.5%) (Table S37).

### Detection at a Targeted False-Call Rate

**Detection at thresholds set within each design depended on the analysis and contrast; achieved false-call rates often exceeded their targets (Fig 4**; **Table S34).** With thresholds targeting a 5% chance of any false call over 5 assessments under independence, a single assessment detected 100% of 51 healthy volunteers’ assembled sessions and 95.1% of 61 stroke sessions with the Goldfine 2011 statistic and 2.0% and 16.4% with the du Bois 2026 paired t test, at held-out false-call rates of 1.0% to 1.1% per assessment (target, 1.0%); the Guger 2018 statistic detected 7.7% of 13 volunteers and 30.3% of 76 tactile runs at 1.8% and 2.6%, above target, so these figures are optimistic. On up to 10 null runs per patient in clinical EEG, these independence-based Guger 2018 thresholds gave measured patient-level false-call rates of 6.2% (95% CI, 4.5%-8.3%) over 5 runs and 6.4% (3.8%-10.0%) over 10, slightly above 5% (Table S38).

**Figure 4.**
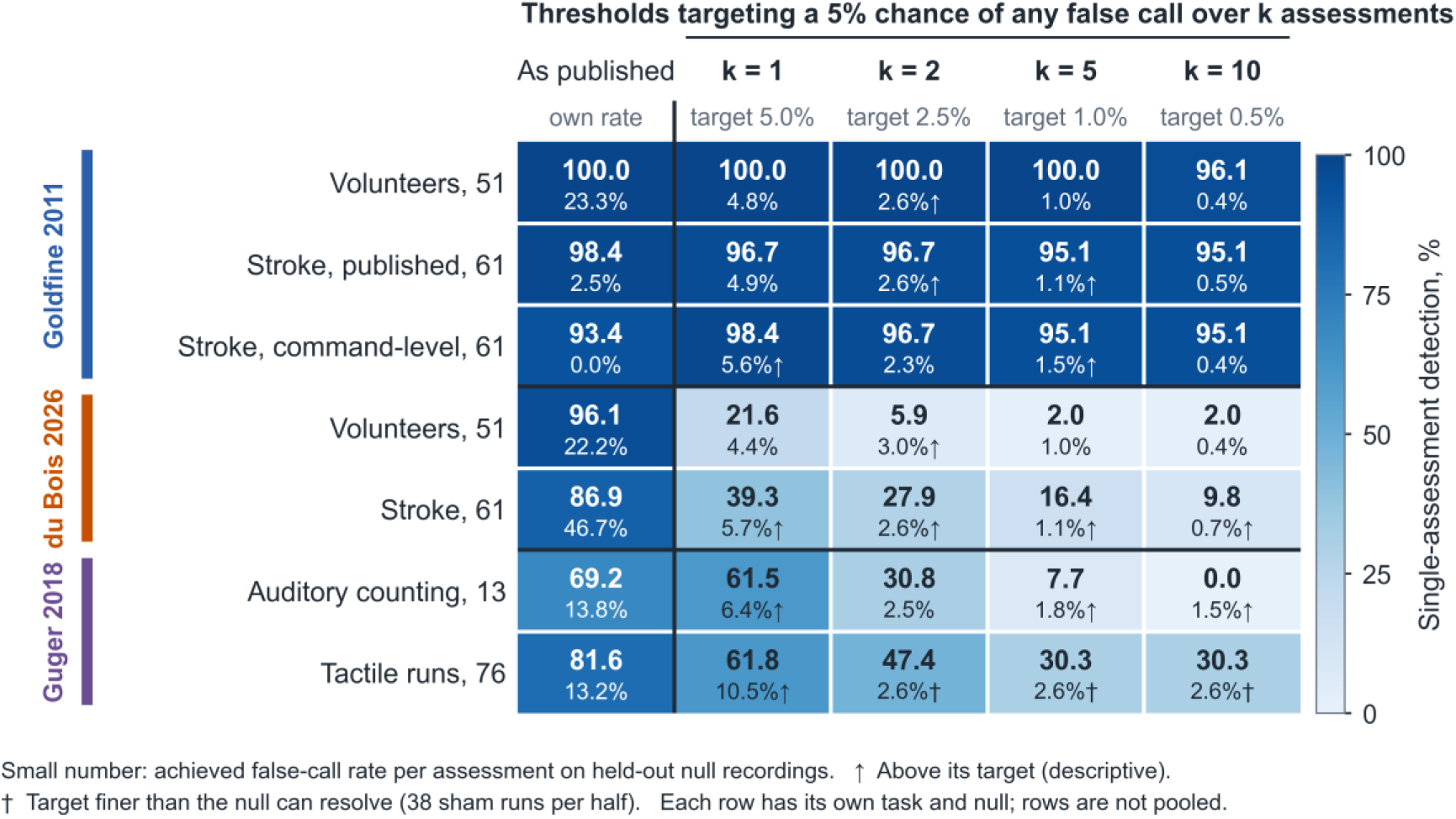
Task-Specific Detection at a Targeted False-Call Rate. Large number and colour: single-assessment detection of genuine responses (%). Small number: achieved false-call rate per assessment on held-out null recordings of the same design, pooled over null units; its 95% CI in Table S34 comes from 2,000 resamples of participants or patients, with both thresholds re-estimated in each, so it reflects runs and draws clustered within a person. Arrow: achieved rate above its target, descriptive (not tested); dagger: target finer than the null can resolve (38 sham runs per half); detection in these cells is optimistic. Measured directly over repeated null runs, the patient-level rates of the Guger 2018 thresholds were 6.2% over 5 runs and 6.4% over 10 (Table S38). Rows: designs, grouped by analysis, with the number of participants, sessions or runs; each has its own task and null; rows are not pooled or ranked, and detection is a task-specific operating point, not clinical sensitivity. Columns: the published threshold, with its rate on the design’s own null (for volunteer designs, the clinical EEG rate of Figure 1), and thresholds set on null recordings, cross-fitted by patient, targeting a 5% chance of any false call over k = 1, 2, 5 or 10 assessments under independence (per-assessment targets 5.0%, 2.5%, 1.0% and 0.5%). Goldfine 2011: healthy volunteers’ assembled motor imagery sessions; stroke sessions with the published variance and the command-level variance (t reference). du Bois 2026: paired t test. Guger 2018: auditory counting; tactile runs.

## Discussion

Run on patients given no study command, 3 of 10 reimplemented published EEG analyses for covert command-following called about 1 in 7 to 1 in 4 patients positive, and more when repeated runs were allowed. The same rules called 12% to 27% of comatose patients 24 hours after cardiac arrest, a population in which task-based testing is studied^39^ before guideline-based prognostication,^40^ and the rates held in patients analyzed after the rules were fixed. The other 7 analyses called few patients, although Cruse 2011’s 2% in new windows exceeded the 0.5% its rule implies (Table S39). All 3 analyses with excess calls detected genuine responses, so their false calls do not reflect implementations that cannot detect.

What makes a positive result interpretable is the complete patient-level rule, including the runs allowed, its false-call rate on recordings without commands and its detection of genuine responders. The three analyses with excess calls behaved differently; only for Goldfine 2011 was a correction fixed in advance and tested in new patients. Goldfine 2011 called no white-noise recordings, so its calls arise from properties of EEG: its jackknife treats tapered estimates within a command as independent, and on clinical EEG a variance that respects this dependence was a median 3.9 times that expected for independent estimates. With whole commands as the jackknife unit, fixed in advance, false calls fell below 5% in new patients and after cardiac arrest while stroke imagery against rest was detected in 93.4% of sessions. Left-against right-hand imagery at the same time after the cue was detected in few acute-stroke patients by either variance; detection against rest therefore shows task-associated EEG differences, not that the test isolates motor imagery, and the corrected variance, at 0.6% to 2.4%, may be conservative. In our implementation, the du Bois 2026 rule compares a long task window’s peak accuracy with a short baseline’s; a longer window’s maximum is larger by chance, and the rule called more white-noise recordings than EEG recordings. An implementation following more of the publication’s stated choices called more windows, mostly through 1 unstated choice, whether cross-validation folds respect the task blocks, and its permutation evaluation did not hold 5% with random folds. The complete decision pathway, more than the classifier, sets the rate, and the 22% describes the advancement step as implemented, an entry criterion for training rather than the publication’s full evidence of task decoding.^30^ The Guger 2018 threshold would be met about 12% of the time by chance if its 10 split halves were independent,^31^ close to its rates on EEG and white noise, a permissive threshold rather than a failed error guarantee: the implementation detected 95% of tactile attention sessions, and with real touch to an unattended finger it was met in 13% of runs, near that chance rate. Implementations written independently from the same texts disagreed with ours, so a publication does not fix a single executable analysis. Detecting healthy volunteers, the usual check that an analysis works,^25,31^ does not establish specificity: readings that detected most healthy responders also called over a quarter of shuffled sessions, and the du Bois 2026 criterion was met in about half to two thirds of stroke sessions with random labels.

Thresholds set on null recordings of each design show how much detection survives, although achieved false-call rates often exceeded their targets, by most for Guger 2018. At thresholds for 5 assessments, the Goldfine 2011 statistic detected 95% to 100% of imagery against rest, and the du Bois 2026 and Guger 2018 statistics 2% to 30%. The contrasts differ, so these results do not rank the methods; they show that detection must be read at the achieved false-call rate of its rule.

These are false calls without commands, not clinical false-positive rates in patients who hear commands without following them. The original studies delivered commands to different cohorts with different preprocessing, and these results do not show that their patients were false positives. The analyses come from publications of 2011 to 2026; whether laboratories using current, locally validated protocols have these rates was not tested. Missed responses also cause harm, and a negative result does not exclude awareness:^2^ Guger 2017 called only 1 in 5 awake patients with acute stroke positive when asked to imagine hand movements, without verified task performance. Because bedside responsiveness fluctuates, repeated assessment is recommended,^41,42^ and each added EEG run compounds the chance of a false call unless the threshold accounts for it. Randomizing trial order cut false calls but lowered detection for some analyses, and its feasibility for patients was not tested.

Extending calls for standard reporting,^6,43^ we recommend that a positive EEG command-following result be reported with the false-call rate of its exact call rule on command-free recordings, including how many runs may count, and its detection rate in known responders at that false-call rate. That rate describes chance behavior, not diagnostic specificity, which requires delivered commands in the target population.

This study has limitations. First, the analyses were reimplemented from publications and public code, without vendor software, the original authors’ review or review by an independent EEG expert; independent implementations of 2 analyses disagreed with ours, and our du Bois 2026 implementation departs from several stated choices, which moved its rate in both directions (Table S30), so the rates describe the published rules as implemented here (positive-control evidence for each, Table S23). Second, the real-task controls used healthy volunteers and patients after stroke, not patients with disorders of consciousness, and a tactile Braille task rather than the vendor’s vibrotactile paradigm; no recordings of the original studies’ patients were available, the repeated-search permutation evaluation used development windows only, and the corrected Goldfine 2011 variance was tested with commands only in stroke recordings, high against rest and low between imagery types. Third, the choice rule and 2 corrections were set after development rates had been seen; the replication was internal to one corpus and had no background labels, and the longest schedules ran later, only where recordings held them (after arrest, 231 of 544). Fourth, recordings without study commands may contain unannotated activation procedures, although rates were similar in awake windows without detected activation. Fifth, the DoC set was small, had 6 channels (Claassen 2019 ran on 6 of its 19) and was exploratory; post-arrest windows were short, sedation was not recorded, and associations with outcome do not validate consciousness detection; no open EEG with delivered commands in disorders of consciousness was available. Sixth, assembled healthy sessions lack slow drift, so leave-block-out detection is optimistic. Seventh, repeated runs were windows of one recording.

A positive EEG command-following result should not be interpreted from the algorithm name or nominal threshold alone. Clinicians need the complete patient-level call rule, its false-call rate in a negative control relevant to the paradigm, and its ability to detect genuine responses under matching conditions at that false-call rate.

## Data Availability

The SpikeNet 2.0 and MORGOTH datasets are available through the Brain Data Science Platform to credentialed users under its data use agreement. The I-CARE database was obtained from PhysioNet and is now distributed through the Brain Data Science Platform under its data use agreement. The prolonged-DoC polysomnography is openly available on Mendeley Data, and the healthy-participant and stroke datasets are openly available from their repositories (Table S1 in Supplementary Data S1). The analysis code and the classifications are available at https://github.com/Alon-Gorenshtein/False-Positive-Calls-Covert-Command-Following-EEG.

## Acknowledgment

A.G. and E.K. were supported by the Clinical and Translational Science Awards (CTSA) grant UL1TR002541 from the National Center for Advancing Translational Sciences, through the Harvard Catalyst | The Harvard Clinical and Translational Science Center Pilot Award Program. The content is solely the responsibility of the authors and does not necessarily represent the official views of the National Institutes of Health.

## Author Contributions

A.G., Y.B., E.K. and O.D. contributed to the conception and design of the study; A.G., Y.A. and Y.B. contributed to the acquisition and analysis of data; A.G. contributed to drafting the text and preparing the figures. All authors critically reviewed the manuscript and approved the final version.

## Data Availability

The SpikeNet 2.0 and MORGOTH datasets are available through the Brain Data Science Platform to credentialed users under its data use agreement.^13,15^ The I-CARE database was obtained from PhysioNet^19^ and is now distributed through the Brain Data Science Platform under its data use agreement. The prolonged-DoC polysomnography is openly available on Mendeley Data,^16–18^ and the healthy-participant and stroke datasets are openly available from their repositories (Table S1 in Supplementary Data S1). The analysis code and the classifications are available at https://github.com/Alon-Gorenshtein/False-Positive-Calls-Covert-Command-Following-EEG.

## Use of Artificial Intelligence

During the preparation of this work the authors used Claude (Anthropic) and ChatGPT and Codex (OpenAI) to assist with writing the simulation and analysis code, drafting and editing text, and proofreading. All AI-assisted code, analyses and text were validated, manually checked and audited by the authors at each phase of the study. The authors take full responsibility for the content of this Article.

## Supporting Information

Supplementary Data S1: Supplementary methods, results, tables, figure and references. Supplementary Data S2: Supplementary tables (one worksheet per table).

