## Supplementary Data S1 for "False-Positive Calls of Reimplemented Published Analyses for Covert Command-Following on Electroencephalograms Recorded Without Study Commands"

This supplement accompanies: False-Positive Calls of Reimplemented Published Analyses for Covert Command-Following on Electroencephalograms Recorded Without Study Commands.

Authors: Alon Gorenshtein, MD; Yosef Adiniaev; Yiftach Barash, MD; Eyal Klang, MD; Oved Daniel, MD.

#### Contents

- Supplementary Methods 1. Datasets, licences and splits
- Supplementary Methods 2. Published analyses and their reimplementation
- Supplementary Methods 3. Blind classifications and eligibility
- Supplementary Methods 4. Reading selection, cross-fitting and statistical inference
- Supplementary Methods 5. Overlays, artifact rules and activation-window exclusion
- Supplementary Methods 6. Per-patient rules and the first-k definition
- Supplementary Methods 7. Randomization-based design
- Supplementary Methods 8. Simulated responses and the size-corrected comparator
- Supplementary Methods 9. Healthy-volunteer sessions

- Supplementary Methods 10. White noise and the synthetic envelope atlas
- Supplementary Methods 11. Mechanism analyses
- Supplementary Methods 12. Order of analyses and corrections
- Supplementary Methods 13. Reporting, software and code availability
- Supplementary Methods 14. Post-arrest coma arm
- Supplementary Methods 15. Replication set
- Supplementary Methods 16. Independent implementation, repeated recordings, outcome exposure, published cohorts and patient positive controls
- Supplementary Methods 17. Genuine tasks, sham labels, the du Bois 2026 implementations and calibration within each design
- Supplementary Methods 18. Full-length schedules and Claassen 2019 on six sites
- Supplementary Results 1. Sensory responses read as volition
- Supplementary Results 2. Cohorts and false calls by analysis in clinical EEG
- Supplementary Results 3. Per-patient rules over repeated assessments
- Supplementary Results 4. Prolonged disorders of consciousness and EEG background
- Supplementary Results 5. Synthetic noise and below-chance accuracy
- Supplementary Results 6. Healthy volunteers
- Supplementary Results 7. Randomization-based design
- Supplementary Results 8. Mechanism analyses
- Supplementary Results 9. Sensitivity analyses and classification agreement

- Supplementary Results 10. Post-arrest coma
- Supplementary Results 11. Replication
- Supplementary Results 12. Independent implementation, repeated recordings, outcome exposure, published cohorts and patient positive controls
- Supplementary Results 13. Genuine tasks, sham labels, the du Bois 2026 implementations and calibration within each design
- Supplementary Results 14. Full-length schedules and Claassen 2019 on six sites
- Table S1. Datasets, licences and splits
- Table S2. Published analyses and their reimplementation
- Table S3 (Supplementary Data S2). Every classified choice, with both blind classifications and the verbatim quotation
- Table S4. Departures from the publications shared by every reading of an analysis
- Table S5. Sensory responses read as volition
- Table S6. Cohort assembly, first-window background and the prolonged-DoC cohort
- Table S7. False-call rates by analysis and dataset (primary, originally planned, generous, strict, cross-fitted, eligible range)
- Table S8. Shortened-to-fit readings and the covert-processing analysis
- Table S9. Per-patient rules
- Table S10 (Supplementary Data S2). False-call rates by EEG background

- Table S11. White noise, the synthetic envelope atlas and the Cruse 2012 diagnostic
- Table S12 (Supplementary Data S2). Below-chance accuracy
- Table S13. Healthy-participant sessions
- Table S14. False-call control and detection of the randomization-based versions
- Table S15. Mechanism analyses
- Table S16. Post-arrest coma: false calls, readings, outcome, non-stationarity and background
- Table S17. Replication of the fixed readings
- Table S18. The du Bois 2026 decision pathway
- Table S19. Stated rates, white-noise rates and clinical EEG rates of each call rule
- Table S20. Clinical EEG readings applied to post-arrest coma; usability bounds
- Table S21. Ranges of rates by reading category (Table S21B in Supplementary Data S2)
- Table S22. Illustrative share of positive calls that would be false
- Table S23. Patient-level decision summary
- Table S24. Independent implementation of the du Bois 2026 and Guger 2018 analyses
- Table S24B (Supplementary Data S2). Choices recorded by the independent implementation

- Table S25 (Supplementary Data S2). Repeated recordings per patient: Claassen 2019 and Edlow 2017
- Table S26. Post-arrest coma: patients called positive by outcome
- Table S27. Published positive rates against the no-command rate of the same rule
- Table S28. Positive control in acute stroke
- Table S30. The second du Bois 2026 implementation, our departures from it and the publication's permutation evaluation
- Table S31. Goldfine 2011 with a variance that respects dependence within commands
- Table S32. Tactile P300 positive control with genuine attention
- Table S33. Stroke motor imagery on its recorded design
- Table S34. Detection at thresholds targeting a patient-level false-call rate, set within each design
- Table S35. The command-level Goldfine 2011 variance, fixed before testing, in new windows
- Table S36. Cluster-aware inference for the genuine-task controls
- Table S37. Time-matched specificity of Goldfine 2011
- Table S38. Patient-level calibration measured directly on repeated null runs
- Table S39. Analyses run later at full length, and Claassen 2019 on six sites
- Supplementary Figure S1. Detection and false calls in healthy volunteers

- Tables S3, S7B, S7C, S7D, S7E, S9A, S10, S11A, S11B, S11C, S11F, S12, S13A, S13B, S13C, S13F, S14B, S15A, S16B, S21B, S24B, S25 and S29 are in Supplementary Data S2 (spreadsheet), one sheet per table.
- Supplementary References

### Supplementary Methods

#### Supplementary Methods 1. Datasets, licences and splits

Clinical EEG came from the SpikeNet 2.0 corpus of the Brain Data Science Platform (BDSP), a collection of de-identified routine, epilepsy-monitoring and long-term recordings from three institutions [12,13]. The recordings were staged in the standard 19-channel 10-20 montage at 128 Hz and were analyzed as stored. The files carry no care-setting, medication or behavioral data, so the setting of a recording (intensive care, routine or epilepsy monitoring) is unknown. A validated EEG foundation model assigned each window one background label from a fixed hierarchy (burst suppression, then periodic patterns, then sleep, then diffuse slowing, then normal awake, then other awake) [14,15]. The background of an analysis window was the label held by at least 60% of its valid model windows; a window without such a majority was labelled mixed, a window with fewer than half valid model windows was labelled unlabelled, and a window whose first and second halves had different majority labels was flagged as transitioning. Diffuse slowing, burst suppression and periodic patterns are called ICU-typical backgrounds. The label is a model-assigned proxy for the pattern of the EEG and says nothing about where the recording was made.

Prolonged disorders of consciousness were represented by overnight polysomnograms released on Mendeley Data, with diagnoses of vegetative state or unresponsive wakefulness syndrome and of minimally conscious state made with the Coma Recovery Scale-Revised [17,18]. The files carry six scalp EEG channels (F3, F4, C3, C4, O1, O2), so each analysis ran on this reduced montage, and Claassen 2019, whose released pipeline uses 19 sites, was run there later on the 6 recorded sites (Supplementary Methods 18). One file was a byte-identical duplicate of another and was removed before the development split was frozen. The files carry no sleep staging, and the staging model has not been validated on this montage, so a wake-like subset was defined within each recording as the windows whose C3 and C4 log delta/alpha power ratio fell below the recording's median. This subset is called lower-delta (wake-like), never wake. The clinical table released with the recordings was linked to recordings only by a deterministic match of the signal samples (33 of 40 cases matched byte for byte, none ambiguous; diagnosis agreed in all 33 matched cases); header identity fields were never read, and no outcome column was used before the mapping was committed. A preliminary inspection of the table had displayed its columns before the analysis was designed; no outcome or Coma Recovery Scale-Revised value was recorded or used in the design.

Healthy-participant data were used to measure detection of true responses (Supplementary Methods 9) and sensory responses read as volition (Supplementary Results 1). Motor imagery came from OpenBMI [20] and from the EEG Motor Movement/Imagery Database [21], auditory counting from the Kojima recordings [22], and passive oddball recordings from ERP CORE [23] and OpenNeuro ds003061 [24]. A third passive dataset (ds003522) was dropped because its raw files did not fit the available storage. The post-arrest coma arm (I-CARE, version 2.1 [19]) is a secondary outcome of this study (Supplementary Methods 14).

Splits were fixed before any analysis-set result was read. Development recordings were used to build and debug every analysis and were excluded from every reported number: the last 20 recordings in a seeded order of the clinical EEG design sample, 3 prolonged-DoC patients drawn with a fixed seed, 3 participants of each of the two motor imagery datasets and 2 participants of each passive dataset (including the Kojima recordings). Six clinical EEG recordings on which an analysis had computed an outcome while they were still provisional development recordings, before the split was frozen, were excluded from every analysis; four further recordings had only been loaded and were retained. The analysis set added 500 recordings that had not been pulled before the freeze and whose patients are disjoint from the design and training samples (Table S6). Licences, access terms and sample sizes are in Table S1. The clinical EEG recordings were collected under a protocol approved by the Mass General Brigham Institutional Review Board (2012P001929) with a waiver of consent and accessed under the platform's data use agreement; the other datasets are public and de-identified.

#### Supplementary Methods 2. Published analyses and their reimplementation

Eleven published analyses were implemented (Table S2): seven motor-command analyses (Claassen 2019 [1], Goldfine 2011 [9,25], Cruse 2011 [10], Cruse 2012 [26], Edlow

2017 [11], Guger 2017 [27-29] and du Bois 2026 [30]); three event-related potential (ERP) command analyses (Guger 2018 [31,32], Chennu 2013 [34] and Gibson 2016 [33]); and one covert-processing analysis [35] that does not test command-following and is therefore reported separately and never pooled with the others. Each analysis was implemented from the publication's text, supplementary material and, where public, the authors' code. Three grades describe the fidelity of the reimplementation. Claassen 2019 reproduces the authors' public code, including its event-shifting and epoching rules. The Goldfine 2011, Chennu 2013 and Sokoliuk 2021 analyses reproduce the core statistic of a public implementation, with their manual ICA and visual rejection replaced by automated rules. The remaining analyses were reimplemented from the published descriptions, and the mindBEAGLE-type analyses are reimplementations of published descriptions of the vendor's methods; the vendor's software itself was not tested.

The Goldfine 2011 two-group statistic was checked against a line-by-line translation of the public toolbox's two-group test routine (Chronux 2.12) on 30 random inputs: the test statistic agreed to within $4 \times 10^{-15}$ and the theoretical-variance decision was identical. The reimplementation's jackknife variance divides each leave-one-out statistic by the full-sample variance, where the toolbox uses the leave-one-out variance, so the reimplementation's variance was 0.4% to 2.5% larger and its calls can only be fewer. Recomputing the primary Goldfine 2011 reading with the toolbox's variance changed none of the calls on the 342 development recordings held locally (79 calls either way).

For every analysis the paradigm and window, the statistic, the stated call rule, the published number of assessments per patient (k) and the published detection rate in healthy participants are in Table S2. Where a publication left a choice unstated, each defensible reading was implemented as a separate variant (called a reading below); the number of readings per analysis is in Table S7E in Supplementary Data S2. For example, the Goldfine 2011 publications cite both Benjamini-Hochberg [e1] and Benjamini-Yekutieli [e2] false discovery rate control without saying which was applied, so both were run, in the multitaper spectral test of the Chronux toolbox [e3,e4] on a Hjorth Laplacian montage [e5]. The common-spatial-pattern analyses [e6] used the chance bounds that their publications state for the number of trials kept [e7], and the ERP cluster tests are cluster-mass permutation tests [e8]. All readings of an analysis share a set of departures from the publication, chiefly fewer EEG channels, no manual artifact review and a simplified filtering order (Table S4). Some follow from the available recordings; others, such as the du Bois 2026 filter pairs, mutual-information estimator, time step and peak refinement, were implementation choices, whose effect is measured in Table S30. Their expected direction is stated where it could be argued. They apply to every reading and cannot be separated from the readings, so they could move all rates up or down together.

Each analysis's own trial schedule (number of trials, timing, block structure and paradigm) defines the window it needs. For example, the Claassen 2019 schedule needs 1,505 s, the Goldfine 2011 1,065 s, Cruse 2011 3,244 s and Chennu 2013 2,530 s. Where the native length did not fit a 30-minute clinical window, a shortened twin was run on the same overlay (3 instead of 4 blocks for Cruse 2011, 4 instead of

5 blocks for Cruse 2012 and 12 instead of 20 blocks for Chennu 2013). Shortened readings are never eligible for the primary reading and are reported in Table S8A. The full-length schedules were run later on longer windows of the same patients (Supplementary Methods 18).

#### Supplementary Methods 3. Blind classifications and eligibility

Every varied choice of every analysis (117 classified choices; Table S3 in Supplementary Data S2) was compared with the publications' text and assigned one of four classes: stated, when the publication fixes the choice; not stated, when the publication is silent or ambiguous about it; contradicting, when the reading departs from what the publication states; and shortened to fit, when the only departure is a length reduced to fit the recording. Each class was tied to a verbatim quotation. Where a publication was silent and the authors' public code fixed the choice, the code counted as stating it. Classifications were produced on October 18, 2026, in sessions of a large language model that were barred from result files and, by their tool records, opened none. The first classification was made in the two sessions that had compiled the analysis catalogues from the publications since October 13 (motor and event-related potential analyses). A third session spot-checked at least 3 of its quotations per analysis against the sources and proposed corrections, all applied, 4 of which changed whether a reading was eligible. The second classification was made in one further session that could not see the first classification's classes. The checker's proposals, the convention that public code states a choice the publication leaves silent, the classes of constants shared by all readings, of shortened readings and of the Claassen 2019 call, the top-down endpoint convention below (which set 4 choices of Chennu 2013 and Gibson 2016 to contradicting: 1 in the first classification only, on a point the checker had left open and that the second classification also called contradicting, and 3 in both classifications) and the either-classifier rule, fixed before the second classification was returned, were decided in the session that coordinated the study, which ran the same model and had access to the false-call rates; other disagreements between the two classifications were not adjudicated but resolved by the either-classifier rule. No EEG expert reviewed the classifications independently. The instructions are summarized by session in Table S29 in Supplementary Data S2; their fixed elements were as follows.

*Both classifications.* Assign each varied choice one class (stated; not stated, including ambiguous; contradicting; shortened to fit), with a verbatim quotation and its source, without looking at any false-call rate.

*First classification.* Classify every value on which a variant differs from the other variants of its analysis, from the analysis catalogues and, where they are silent, the publications; mark variants that cannot call. Do not open the output tables, intermediate outputs or any results log. For Guger 2018, accuracy above 23% is stated and the number of trials behind it is not.

*Second classification.* Classify every (analysis, choice, value) item from the publications' full texts (verbatim excerpts for 2 paywalled publications). Do not open any output, rate or results file, or the first classification, and run no analysis code. Public code states a choice only where the publication is silent. The publication an analysis is named after is primary; a companion publication fills gaps only where the primary is silent, and a conflict between companions makes the choice not stated. A rule that also accepts bottom-up or nondiscriminative responses contradicts a publication that says such responses do not show top-down engagement. The two classifications agreed on whether a choice was consistent with the publications (stated or not stated) or contradicted them for 88.9% of the 117 classified choices (Cohen κ [e9], 0.73; 95% CI, 0.59-0.86) and agreed on the class for 77.8% (κ, 0.67; 95% CI, 0.56-0.77) across the four classes; the intervals are percentile bootstrap intervals over choices (2,000 resamples). A choice whose first class differed across variants counts once per class (Table S3 in Supplementary Data S2 lists each).

The endpoint of the study is command-following, a top-down act. A reading that accepts a response the publication itself describes as bottom-up, involuntary or nondiscriminative (an orienting P3a, a steady-state sensory response, or any target-versus-baseline response that is the same for all stimuli) was therefore classed as contradicting, even when the publication reports that response. This top-down endpoint convention was applied to Chennu 2013 and Gibson 2016. It was not applied to the Sokoliuk 2021 analysis, which tests covert processing rather than command-following.

A variant was eligible for the primary reading if every one of its choices was stated or not stated under at least one of the two classifications (the either-classifier rule) and it was neither a shortened twin nor unable to call. A variant is unable to call when its minimum attainable P value exceeds the alpha it uses. Union-eligible variants numbered 68 (50 motor and 18 ERP, including 4 for Sokoliuk 2021). A stricter sensitivity rule required both classifications to judge every choice stated or not stated; 56 variants met it (45 motor and 11 ERP), all within the 68. Shortened twins were listed when a variant had a shortened choice and no choice that both classifications called contradicting.

The call rule is a separate question from eligibility, and the following principle was applied to it: "A variant has a STATED call rule only when EVERY axis that sets the call criterion (which test or contrast decides) or its threshold (alpha, CI level, cut-off, correction) is STATED. Axes that define the statistic, the null's construction (permutation unit), cross-validation, or preprocessing are analysis choices; they stay in the STATED/NOT_STATED eligibility rule, as now." The choices that set the call criterion or its threshold are, by analysis: the patient-level call (Claassen 2019 and du Bois 2026), the accuracy threshold (mindBEAGLE-type analyses), the second outcome criterion and the false discovery rate procedure (Goldfine 2011), the method, baseline check and interval level (Cruse 2011), the tails and cluster alpha (Gibson 2016), and the patient-level rule (Chennu 2013 and Sokoliuk 2021). For Cruse 2012 and Edlow 2017 the call is common to all readings and stated. Among eligible readings, only those with a stated call rule were candidates for the primary reading wherever any had one. Where none had one, all eligible readings were candidates, and the analysis is labelled "call rule not stated" or "not fully stated" in Table S7A.

#### Supplementary Methods 4. Reading selection, cross-fitting and statistical inference

Within each analysis and dataset, the primary reading was the eligible reading with the fewest false calls on the recordings where every eligible candidate ran (the common recordings), among readings usable on at least 70% of the recordings they were attempted on (attempted recordings are those with a result, no usable trial or an error; a window that does not fit is not attempted). Without the usability floor, "fewest false calls" could select a reading whose artifact rule leaves a small, selected subset. A recording in which no trial survived an analysis's own artifact rule left that analysis's denominator and was counted by stratum.

Selecting the lowest-rate reading biases its rate downward, so rates were cross-fitted. Patients were split into two halves by the parity of a checksum of the patient identifier; the reading was selected on one half and its rate estimated on the other, in both directions, and the pooled held-out counts were tested. The cross-fitted rate is the average of the two held-out rates. The significance of an analysis was the one-sided exact binomial test of its pooled held-out count against 5%, with Holm adjustment [e10] across the call-rule analyses of the same clinical EEG null set (7 motor-command analyses on the no-command overlays and 2 ERP analyses on the no-stimulus overlays). An analysis was inflated if its Holm-adjusted P was below .05. A single family of 9 would give Claassen 2019 a Holm-adjusted P of .26 (primary), .30 (generous) and .22 (strict) in place of .22, .26 and .17; no conclusion changes. The selected variant's own full-sample rate is shown with an exact

Clopper-Pearson interval [e11] but was not tested, because selection makes its P value uninterpretable. The prolonged-DoC data were used for estimation only, with no tests and no multiplicity adjustment. Bootstrap intervals [e12] for cross-fitted rates and for the Claassen 2019 two-recording bracket came from patient-level resampling with relabelled draws (2,000 resamples), because keeping the original identifiers of repeated draws would merge them and narrow the interval.

The range across eligible readings describes how far a rate depends on unstated choices. Its endpoints are selected readings (the lowest and the highest rate among eligible readings), not estimates of any quantity, and each endpoint is therefore shown with its own exact interval and labelled "selected, not an estimate" (Table S7E in Supplementary Data S2). The range is not a confidence interval. The originally planned reading (the planned lowest-rate reading among all defensible readings, on the original rows) and the a priori default reading (the default of the implementation, fixed before any rate was read, with a fidelity grade of the default) are shown beside every primary rate (Table S7A; Table S7B in Supplementary Data S2).

All other proportions carry exact Clopper-Pearson 95% CIs. Unless a one-sided test was stated, tests were two-sided at .05. Two-group comparisons of detection used the patient-paired bootstrap difference with the margin stated in Supplementary Methods 8.

The usability floor of 70% excluded no eligible reading: the lowest usable share of any clinical EEG reading was 78.1% (motor imagery), and the three inflated primary readings were usable in 99.3% (Goldfine 2011, 703 of 708), 99.9% (du Bois 2026, 707 of 708) and 93.9% (Guger 2018, 850 of 905) of attempted recordings. Because the two held-out counts of a cross-fitted rate are not independent, each half was also tested alone: for the 3 inflated analyses every half-specific one-sided exact P was below $10^{-13}$ (Goldfine 2011, 91 of 369 and 73 of 334; du Bois 2026, 73 of 372 and 84 of 335; Guger 2018, 68 of 427 and 61 of 414), so no conclusion rests on pooling the halves. If every unusable recording had been negative, or positive, the three rates would lie between 23.2% and 23.9% (Goldfine 2011), 22.2% and 22.3% (du Bois 2026) and 12.9% and 19.0% (Guger 2018) (Table S20).

#### Supplementary Methods 5. Overlays, artifact rules and activation-window exclusion

For every patient and analysis, the analysis's own schedule was placed on a window of the recording long enough to hold it. Motor-command analyses received command labels on EEG with no commands and ERP analyses received stimulus markers on EEG with no stimuli, so every positive call is false by construction. Windows were drawn from a seeded permutation of the clean starts of each recording. A start was clean when the run and a 30-s margin on each side contained no padding, defined as seconds with exact zeros on any overlay channel and stretches of at least 60 s in which every overlay channel stayed below 2 μV. Large artifacts were kept on purpose: no artifact rejection was applied beyond each analysis's own rule. Overlays on one recording were accepted in order if they did not overlap, one overlay per patient was the primary unit, and a further overlay of a recording could overlap an earlier one only after all non-overlapping windows were used; those overlays were labelled. The artifact rule of each analysis was applied inside the analysis (for example, trials beyond ±100 μV for Guger 2017 and columns above 200 μV for Edlow 2017), and a recording in which no trial survived left that analysis's denominator. Denominators therefore differ by analysis, from 147 patients for the four-block Cruse 2011 schedule to 850 for Guger 2018 (Table S6B).

Routine and epilepsy-monitoring EEG contain eye opening and closure, photic stimulation, hyperventilation and reactivity testing, which are external block-structured drivers. Because the clinical EEG carries no event annotations, windows overlapping photic stimulation (stepped narrow-band lines at the flash frequencies and their harmonics at O1 and O2) or regular eye opening and closure (blink transients at Fp1 and Fp2 paired with an alpha change at O1 and O2) were identified by automated detectors whose thresholds were set on synthetic signals and the development recordings only. Windows flagged by either detector were excluded from the awake-background and ICU-typical rates (Table S10 in Supplementary Data S2) and from the activation-clean sensitivity analysis of the mechanism model (Table S15B). In the prolonged-DoC recordings only the five annotations in four files were used, and a "lights on" annotation was treated as stimulation. Hyperventilation and reactivity testing have no detector, and the recording type is unknown; both are limitations. The detectors are conservative: a photic session under strong ECG pick-up can be missed, and spontaneous drowsy eye closure is not flagged and remains in the background labels.

#### Supplementary Methods 6. Per-patient rules and the first-k definition

Each analysis's per-patient rule was applied at its published number of assessments k (Table S2): k = 1 for most analyses, k = 2 for the du Bois 2026 criterion (sessions 1 and 2; the publication uses it to advance patients to brain-computer interface training, and a pass is counted here as a positive call because it requires task decoding above baseline) and for Guger 2018 (two runs), and a bracket for Claassen 2019, whose publication reports 1 to 8 recordings per patient (mean 2.3). The patient's first k usable windows were taken in the order of a seeded random numbering of the recording's windows, so the first k are the k windows with the lowest overlay indices and not the k earliest in time. A patient was positive when at least one of the k windows was called. Patients with fewer than k usable windows left the denominator and are counted in Table S9A in Supplementary Data S2. For du Bois 2026, which governs advancement to further training rather than a diagnosis, the rule is a pass in either of the first 2 sessions. For Guger 2018 only the 3-stimulus paradigm had an eligible reading, so both runs were of that paradigm, and the best-of-k accumulation is reported descriptively up to 7 runs, the most reported per patient, and beyond that range only for the prolonged-DoC nights (Table S9C).

The du Bois 2026 publication advances a patient "when DA exceeded 70% or when task-related DAs were statistically greater than baseline (one-tailed paired t-test, $\alpha = 0.05$)", calls the t test alone a significant run, and describes a label-permutation test (100 permutations) as an evaluation against chance whose curves "were plotted together for visual comparison", without stating that the permutation test enters advancement [30]. The primary call is therefore the advancement rule. Table S18 reports the pathway four ways: advancement, significant run, permutation alone, and advancement together with permutation. The permutation P came from the originally planned permutation reading of the same window; its leave-one-block-out folds give the same configuration and fold-mean peak time as the primary reading, and every pair matched on window start and peak time.

The BDSP files carry no dates, so two recordings of a patient from different days cannot be built. The Claassen 2019 rule was therefore bracketed. The lower bound used two windows of one recording (14 recordings were long enough); the upper bound used all pairs of different patients' first windows, with Benjamini-Hochberg control within the pair and a bootstrap that resamples patients and recomputes all pairs. The true rate for a patient assessed twice is expected to lie between the two when calls are positively associated within a patient; this is not guaranteed for a rule that corrects across recordings, so the bracket is approximate (Table S9B).

#### Supplementary Methods 7. Randomization-based design

For each analysis the randomization-based version kept the analysis's features, classifier and statistic but randomized the order of trials trial by trial and tested the statistic by a randomization test at that unit: the trial labels were redrawn under the same scheme and the statistic recomputed (200 draws, with early stopping that is exact for the decision). A redraw below an analysis's minimum trial count used rejection sampling (a conditional randomization test capped at 10,000 draws), never a zero statistic; a non-finite null draw counted as an exceedance, and a non-finite observed statistic made the recording unusable. The version was run once per timing group, that is, per set of readings that share trial count, onsets, response duration and paradigm. For sensory-response data a label permutation on physically different stimuli is not a volition null, so the family has no randomized arm; a sham-instruction randomization test, exact by construction, is reported only as a construction check (Supplementary Results 1).

The false-call bar for the randomization-based version was a point estimate of 5% or less with no Holm-significant one-sided excess in any dataset, background stratum or recording unit (Table S14A). A point estimate above 5% with no significant excess is labelled "not met (point estimate above 5%)" and is not described as a failure of validity.

#### Supplementary Methods 8. Simulated responses and the size-corrected comparator

Detection by the randomization-based version was compared with detection by the published analysis on simulated source-level responses added to the no-command EEG. A hand-area source with a fixed template lead field was given a smooth amplitude ramp. Because the response is added at the source level, it spreads to neighbouring channels as volume conduction would, so analyses that use a current-source-density or Laplacian transform are not favoured. The response was a desynchronization or synchronization of relative depth 0.2, 0.4 or 0.6 for motor analyses, and a template waveform with latency jitter and template topography of 1.0, 2.5 or 5.0 μV for ERP analyses. Onset jitter was 0 to 1 s and the response probability was 1 or 0.5. No responses were added in suppression or burst suppression. The grid was fixed before analysis and never used for tuning.

The comparator was the published analysis size-corrected to a 5% false-call rate: the threshold on its continuous statistic was the 95th percentile of its null statistic, taken from one half of the patients and applied to the other half, then swapped and averaged. A stratum's own threshold was used only when the calibrating half held at least 100 null draws in that stratum; otherwise the pooled threshold of that half was used and the rows were flagged. For each analysis the evaluation point was fixed on a seeded 60-patient sample used to locate the evaluation point (the locator sample) as the smallest grid effect at which the corrected comparator reached 50% pooled sensitivity (the largest grid effect, labelled "50% not reached", if none did) and sensitivity was measured on patients outside the locator. The comparator of a timing group was its reading with the highest mean size-corrected sensitivity over the grid on the locator. Noninferiority required the lower bound of the 95% interval of (randomization-based minus size-corrected comparator), by patient-paired bootstrap, to exceed -5 percentage points. An uncorrected comparison, run on the next 100 seeded patients and labelled inflated, and a comparison of P-based against statistic-based size correction (flagged when P-based correction was more than 5 points more sensitive; none were) were also run. The achieved size of the corrected comparators on the locator was 3.8% to 11.5% (4.0% to 5.9% excluding Cruse 2011). Results are in Table S14C and in Table S14B in Supplementary Data S2.

#### Supplementary Methods 9. Healthy-volunteer sessions

Detection was measured on healthy participants' own motor imagery and auditory counting, not on simulated responses. Each analysis's schedule was assembled as a pseudo-session from separate trial excerpts of a participant's runs. Excerpts were cross-faded at splices, because splice discontinuities are detectable features, and each excerpt was used once. The same sessions were then analyzed with the class labels permuted over source units (label-shuffled controls), which leaves no task contrast, so every call is false. Participants and data: OpenBMI session 1 for Claassen 2019, du Bois 2026 and Edlow 2017; OpenBMI sessions 1 and 2 for Goldfine 2011, the motor imagery analysis and Cruse 2012; Claassen 2019 on a seeded sample of 20 participants; Kojima attended counting for the three ERP analyses; Cruse 2011 had no dataset with hand versus toe movement and was not graded, because every available dataset reuses source trials. The verdict for each analysis, on its default reading, was "fidelity concern" if the upper 95% Clopper-Pearson bound of detection fell below the published healthy-participant rate, and "consistent with the published rate" otherwise; there is no pass or fail label, and a reuse-confounded analysis is never graded.

The original assembly packed disjoint excerpts so that no sample was reused, but a long source trial could host several excerpts that filled slots of the same pseudo-class. In the label-shuffled controls this gave 1.07 to 1.48 slots per source trial, which clusters trials within a class and can inflate the false calls of analyses that treat trials as independent. After an adversarial review noticed this, the assembly was rebuilt with one source trial per slot: the same seeds, schedules, splice grid, cross-fade, shuffle control, tests and verdict logic, with one change, that no source fills two slots (a guard checks this for every session before any analysis runs, and the guard fails on a deliberately broken allocation). The rebuilt sessions cover Claassen 2019 (the same 20 participants on sessions 1 and 2, because one source per slot needs 341 rest slots against about 200 rest stretches in a session), Goldfine 2011 and the motor imagery analysis (51 participants, sessions 1 and 2), and du Bois 2026 and Edlow 2017 (51 participants, session 1). Chennu 2013 and Gibson 2016 were carried over from the original assembly because each unit is already one whole trial. Cruse 2012 (the published session needs 443 to 453 rest slots per session against about 400 rest stretches, with no third session) and Guger 2018 (60 groups per session against 12 trials per counted identity) cannot be constructed with one source per slot. Both assemblies are reported (Table S13).

The 27 stated-rule eligible readings of the Goldfine 2011 (9), du Bois 2026 (14) and Edlow 2017 (4) analyses were also run on the one-source sessions, with true and shuffled versions for each participant (Table S13A in Supplementary Data S2), so that the readings selected for the primary rates can be set against their own detection and shuffled-session false-call rate. The pseudo-sessions contain no slow drift across blocks, so detection by block-wise cross-validation is optimistic on them, and a block-wise reading meets no drift between blocks that a recorded session would contain.

#### Supplementary Methods 10. White noise and the synthetic envelope atlas

A synthetic calibration atlas (C1) tested the implementations and a possible mechanism with no patient data. The noise types were independent Gaussian white noise (SD 20 μV), pink noise with spectral slope 1/f and 1/f squared, stationary alpha-band noise, and EEG-like noise made of spatially mixed pink sources plus alpha, multiplied by a log-normal amplitude envelope with log-scale SD 0, 0.25, 0.5 or 0.75 and correlation time 1, 5, 10, 30 or 60 s. All noise had 19 channels at 128 Hz and was seeded. Each analysis's own run function was applied to its default, planned lowest-rate and randomization-based readings, 60 replicates per cell (30 for Claassen 2019 and du Bois 2026, which were run on 3 cells only: white, pink 1/f and log-scale SD 0.5 at 10 s). The white-noise rate of every stated-rule eligible reading (68) was measured separately on 60 replicates of white noise on the clinical EEG specifications (Table S11A in Supplementary Data S2); readings that need a longer window than the atlas unit (Chennu 2013 with 20 blocks and the six-block Cruse 2011 readings) ran on their own longer draw.

Four criteria were fixed in advance. The implementation bar required the planned lowest-rate reading of each analysis to lie within the binomial 95% range of 5% on white noise (Guger 2018 and du Bois 2026 analyses were declared exceptions in advance). The mechanism bar required a monotone rise of false calls with envelope fluctuation for readings not immune by design, tested by a Cochran-Armitage trend test [e13] per analysis with Holm adjustment. The construction bar required that the randomization-based versions show no trend. A stopping rule specified that if no planned lowest-rate reading of a non-immune analysis exceeded 10% at log-scale SD 0.25 to 0.5, the non-stationarity explanation would be dropped (Table S11E; Table S11F in Supplementary Data S2).

A diagnostic examined the Cruse 2012 white-noise excess (9 of 60 draws). It used 500 fresh white-noise draws, and the same 500 draws shaped to a 1/f spectrum, on the clinical EEG specification, for the two Cruse 2012 cross-validation readings: groups drawn at random within each class and contiguous groups. The same schedules and seeds were shared across cells and arms, so comparisons are paired (exact McNemar test). On the first 353 draws (a one-hour cap on the run), three variants of the permutation null were compared with the analysis's own null, which keeps the cross-validation groups fixed and permutes labels across all trials: a null that permutes labels within each cross-validation fold; groups re-stratified for each permutation; and trials dropped to the number kept in the clinical recordings, with or without re-stratification. The calibration of the analysis's permutation P values was checked on 200 white-noise draws with 1,000 permutations per comparison. Spectrum shape, artifact rejection and epoch overlap were also checked. The within-fold arm was added after the interim rows of the first step had been seen, and the run reached its memory and one-hour limits, so the drop arms rest on 353 draws (347 evaluable) and the calibration check on 200 draws. The diagnostic used synthetic noise only; no clinical recording was read. It does not enter any rate in the main text. A noise spectrum shaped like the clinical recordings was not tested, because no median clinical power-spectrum aggregate was available, so the lower rate in the clinical recordings (2.4%; Fisher P = .006 against the white-noise rate) remains unexplained (Table S11G and 11H).

#### Supplementary Methods 11. Mechanism analyses

The planned mechanism hypothesis (the alignment test, M1) was that false calls arise when the spontaneous slow fluctuations of the analysis's feature align with the rhythm of the design's labels, so that a trial or epoch permutation, which assumes exchangeable trials, is not calibrated. For each analysis the predictor X was the predicted variance inflation of the label contrast relative to a flat spectrum: the aperiodic fit of the feature series' periodogram [e14] weighted by the design-expected periodogram of the centred label sequence, with the alternation frequency and its harmonics excluded so that X never uses the in-band power that is the statistic. X was computed on each analysis's own channels and spatial transform. M1 was a logistic regression of the call on z-scored X with reading fixed effects over all 233 readings of the clinical EEG rows, each analysis weighted equally and inference by patient-cluster bootstrap (2,000 draws); the bar was a one-sided P below .05 for an odds ratio per SD above 1. Sensitivities used a random intercept, X from the whole recording, X from each analysis's own bands, X from the first principal component across channels, exclusion of the Claassen 2019 rows, activation-clean windows, and a mask for flat channels and artifacts (Table S15B).

Designs were typed from the predictor on a reference 1/f series as inflated (X above 0.1), immune (absolute X at most 0.1) or deflated (X below -0.1). The planned immunity check asked whether the slope of false calls on the aperiodic exponent was null in immune designs (the upper 95% bound of the odds ratio per SD at most 1.25, and a larger slope in block designs than in immune designs); only the du Bois 2026 training readings and the motor-imagery analysis typed as immune. A pre-stated prediction for deflated designs was a false-call rate at or below the nominal 5% and a negative slope on the exponent. A secondary family tested, with Holm adjustment across the four tests, (S1, the concentration test) concentration of false calls on a subset of patients (a dispersion statistic against a patient-label permutation within analysis), (S2, the background test) organized (normal awake and sleep) versus disorganized (burst suppression, periodic patterns, diffuse slowing) backgrounds, (S3, the artifact test) artifact share per 10 percentage points, and (S4, the level-of-consciousness contrast) the patient-median X of minimally conscious versus vegetative state patients on the reference 12-block schedule (Hodges-Lehmann shift [e15] and Wilcoxon test), and whether X adds to resting markers for the classification (change in AUROC). The background test confounds background with care setting in this corpus, so organized backgrounds include non-ICU recordings, and it supports no statement about patients with disorders of consciousness. Estimates without a bar were the transport of the fitted model to the prolonged-DoC recordings (A2, the transport estimate: calibration intercept and slope) and, for the 30 patients with linked clinical data, the Spearman correlation of X with the Coma Recovery Scale-Revised arousal subscale and the AUROC of X for recovery (A3, the arousal and recovery estimates; exploratory, not powered for prognosis, with no inference about prognosis). A confirmatory replication (A4, the spectrum replication) tested the stopping rule of the earlier design-phase test on patient-disjoint recordings: the share of the block-design excess explained by the recording's own spectrum (R, the excess false-call rate above the nominal 5% in surrogates that keep each recording's power spectrum and randomize its phases [e16], divided by the excess in the recordings themselves) had to be at least 0.5, with a support check that AR-prewhitened series produce at most 6% [e17,e18]. Spindle-like density and slow sigma modulation (B5, the spindle-like activity test) were tested within inflated designs with Holm adjustment across the two tests.

After M1 was not supported, the following were added and are labelled as added wherever reported. A non-stationarity index (C2; the variance over epochs of the analysis's log feature divided by its mean in 10 multivariate phase-randomized surrogates of the same window) was related to false calls in the same model; the Phase-1 rows suggested it and are hypothesis-generating. Its confirmation used a fresh seeded draw of 400 patients disjoint from every earlier sample; the bar was an odds ratio per SD of at least 1.20 with the lower 95% bound above 1. A within-recording check asked whether the index tracks the statistic inside a recording (stop if the pooled slope was at most 0 or the within-recording correlation below 0.10). An alpha-rhythm route related alpha-peak prominence (a periodic-peak fit above the aperiodic background) to false calls (stop if the lower 95% bound of the odds ratio was at most 1; go if the odds ratio was at least 1.3 with a lower bound above 1), with a second step for a route-specific interaction. A non-EEG arousal marker (the log SD of the 0.01 to 0.1 Hz heart-rate fluctuation from the ECG of the same file) was related to the log index on the fresh 400 patients (standardized β of at least 0.15 with a one-sided lower 95% bound above 0, with HC3 robust errors [e19] and adjustment for background and artifact share), after a go/no-go test on design data (Spearman correlation of at least 0.10 with an interval excluding 0). Label-aligned heart-rate changes were tested against 40 circularly shifted placebo schedules, with exposed, immune and randomized designs, recording-stratified and artifact-controlled versions. Prolonged-DoC values of the index by diagnosis and outcome were described without tests.

#### Supplementary Methods 12. Order of analyses and corrections

The items below changed or were added after earlier results had been seen, in the order they arose. The stated-rule framework and the analyses that followed it (Supplementary Methods 3, 4 and 9) came after the first rates had been seen.

First, the healthy-participant positive control was reinstated after having been omitted at one stage of the design. The published healthy-participant rate of Edlow 2017 was corrected to 12 of 16 (75.0%), as stated in the paper; the earlier 11 of 16 (68.8%) does not appear in the publication.

Second, after the first results were read, an adversarial review found that the originally planned reading (the reading with the fewest false calls among all defensible readings) could contradict a publication's stated rules; the originally planned du Bois 2026 reading, for example, replaced the publication's stated pass criterion with a label permutation and fell to 5.1%. This prompted the stated-rule framework of Supplementary Methods 3 and 4: the classification of each choice, the either-classifier eligibility rule and a strict sensitivity rule. The originally planned reading is shown beside every primary rate (Table S7A; Table S7B in Supplementary

Data S2), and the planned false-call bar, noninferiority margin and Holm families were not changed.

Third, the same review found that the most favorable reading of one analysis could replace its stated call criterion, and the call-rule restriction of Supplementary Methods 3 was added. Where an analysis has a reading with a stated call rule, only such readings are eligible for the primary reading; this restriction removed the du Bois 2026 reading with a permutation call from the primary reading and made it a generous-reading result.

Fourth, the default readings of the Goldfine 2011 and du Bois 2026 analyses produced rates of 98.2% and 90.0% in the clinical EEG, which were implausibly high. A fidelity check on synthetic noise, run before the headline merge was read, graded the defaults and led to the corrected readings (a jackknife variance with the contiguous-difference criterion for Goldfine 2011, and fold-mean peak times with per-fold values at them for du Bois 2026). They were added after the excess of the uncorrected readings had been seen. They did not enter the original selection; they entered the stated-rule framework as eligible readings. Under that framework they became the primary Goldfine 2011 and du Bois 2026 readings (23.3% and 22.2%, against 38.1% and 5.1% under the originally planned readings).

Fifth, the top-down endpoint convention of Supplementary Methods 3 was ruled after rates had been seen. Its first use, on a point the checker had left open, set the call criterion of the 8 Chennu 2013 readings that call on an explicit-target response against baseline in the P3a window from not stated to contradicting in the first classification (the second classification called them contradicting on its own). Applied later in both classifications, it changed the class of the call criterion of 16 Chennu 2013 readings (from not stated to contradicting) and of the contrast of 6 Gibson 2016 readings (from stated to contradicting). It changed no call test, because neither axis set the call at the earlier classification, but it changed which readings were eligible. The convention was not applied to the covert-processing analysis; two of its readings that the second classification called contradicting on the same argument are excluded from the strict set, and the strict covert-processing rows are flagged.

Sixth, the healthy-volunteer assembly was rebuilt with one source trial per slot (Supplementary Methods 9), and the stated-rule readings were run on those sessions, after the label-shuffled controls of the original assembly had been seen. The result on the original assembly stays as reported.

Seventh, the fresh-patient confirmation of the non-stationarity index was first run with the rows of Cruse 2012 incompletely constructed (355 instead of 710 rows, so 4,269 instead of 4,624 rows in 11 instead of 12 analysis-readings) and met its bar (odds ratio per SD, 1.20; 95% CI, 1.00-1.42). The row construction was corrected and the confirmation rerun on the full rows, where the bar was not met (odds ratio per SD, 1.17; 95% CI, 0.99-1.37). The corrected run is the result; both are in Table S15A in Supplementary Data S2.

Eighth, all mechanism analyses after M1 (the non-stationarity index, the within-recording check, the alpha route, the heart-rate analyses and the synthetic atlas) were added after the planned M1 test was not supported (one-sided P = .06) and the rates of default readings were seen. Their results are exploratory unless a fresh-patient bar was stated and met. Among them, only the coupling of heart-rate fluctuation to the non-stationarity index met a fresh-patient bar (Table S15A in Supplementary Data S2); the association of the index with false calls did not. The confirmatory replication of the planned stopping-rule test (A4) was itself planned.

Ninth, the four-block readings of Cruse 2011 were rerun on four-block windows (148 recordings instead of 26) before the final table, because the seeded start depends on the window size; the interim cells are not used. The per-patient values at the published number of assessments replaced earlier expected-false-calls-per-100 values that used a different set of readings and k.

Tenth, the diagnostic of the Cruse 2012 white-noise excess (Supplementary Methods 10) was run after the excess had been seen. It used synthetic noise only, and was stopped at 353 draws for some arms by a one-hour cap; it does not enter any rate in the main text. Its within-fold arm was added after the interim rows of the first step had been seen.

Eleventh, the white-noise rate of all 68 eligible readings (Supplementary Methods 10, Table S11A in Supplementary Data S2) was measured after the primary readings had been chosen, to describe how far the rates of eligible readings depend on the reading; it was not used for selection.

Twelfth, the post-arrest coma code was corrected twice, both times before any I-CARE outcome or aggregate was computed or viewed (Supplementary Methods 14).

The original code tested for recording padding (an exact-zero second, or every channel below 2 μV for 60 s or more) after the median re-reference, where the median channel is exactly zero by construction, so real data could be read as padding and single-channel dropouts were missed. The first correction applied the rule to the recording as read, skipping a channel flat over the whole segment (the recording reference); the second also treated a channel flat over the window as read as the reference, after 3 patients failed under the first correction and produced no output. Every patient processed under earlier code was rechecked under the final code: 29 patients previously excluded for having no clean window became eligible, 3 were processed for the first time, 121 windows were reprocessed (104 moved and 17 with a changed padding mask only), 266 were unchanged and 18 exclusions stood; no included patient was lost. The old rule had marked 80,005 s as padding in 118 chosen windows; the final rule marked none of them. The 34 exclusions that the original code had decided from the file index or header alone (no segment starting 12 to 36 hours after return of circulation, or none long enough) were rerun under the final code and were unchanged. Because a correction might have changed a patient's window after it was first stored, the 66 windows kept for the arousal analysis (Supplementary Results 10) that no longer matched the rechecked windows were set aside and the rechecked windows read again; the main post-arrest analyses did not use stored windows.

Thirteenth, the stated-rule framework was applied to the post-arrest coma rows after the first post-arrest rates had been computed. The corrected Goldfine 2011 and du Bois 2026 readings were rerun on each patient's stored window; each rerun reproduced the arm's stored result for its base reading. The arousal analysis in post-arrest coma (Supplementary Results 10) is secondary; its code was fixed before the post-arrest outcomes were read. Two further post-arrest analyses were added after the post-arrest outcomes had been read: false calls of each primary reading by background grade (descriptive, Table S16G), and the primary readings' false calls against good outcome with the planned adjustment (Table S16D).

Fourteenth, the replication set (Supplementary Methods 15) was listed, extracted and analyzed after every reading and test of the 7 replicated analyses had been fixed and committed; nothing about these analyses was chosen after any of their rows had been computed. Fifteenth, two exploratory analyses of the replication set were added later: a brain-computer interface test, which is outside this report, and a rerun of the Claassen 2019 development windows. After the Claassen 2019 replication rate fell below its prediction interval, the Claassen 2019 development windows were rerun with the replication's window placement; the paired comparison with the stored calls (two-sided exact McNemar test), the comparison with the replication rate (two-sided Fisher exact test) and the rule for reading the result were fixed in code before the rerun, and windows without a fitting overlay left the paired denominator (Supplementary Results 11). Sixteenth, after review, the main-text post-arrest rates were moved to all 544 patients with a usable window, and their split by background was computed (Table S16H); no reading was reselected and no outcome was read. Seventeenth, after review, the genuine-task controls, the second du Bois 2026 implementation, the Goldfine 2011 command-level variance and the calibration within each design were added (Supplementary

Methods 17); none changes a reading, a rate or a test reported above. Eighteenth, after a second review, the command-level variance was fixed in code and tested in the replication and post-arrest windows, and the time-matched left-right test, the cluster-aware summaries and the direct patient-level calibration were added (Supplementary Methods 17). Nineteenth, after the figures had been drafted, the schedules longer than 30 minutes were run at full length on longer windows, Chennu 2013's 20-block reading was chosen on the development recordings that hold it, and Claassen 2019 was run on the 6 prolonged-DoC sites (Supplementary Methods 18); each reading was fixed before any longer window was read, and none changes a reading, a rate or a test reported above.

#### Supplementary Methods 13. Reporting, software and code availability

Reporting follows the Standards for Reporting of Diagnostic Accuracy Studies (STARD 2015) [e20], adapted to a study with no positive reference cases, and the Strengthening the Reporting of Observational Studies in Epidemiology (STROBE) guideline [e21]. Analyses used Python 3.9.6 (NumPy 2.0.2, SciPy 1.13.1, scikit-learn 1.6.1, pandas 2.3.3); the classifications and analysis code are at https://github.com/Alon-Gorenshtein/False-Positive-Calls-Covert-Command-Following-EEG. Author code of the published analyses is not redistributed.

#### Supplementary Methods 14. Post-arrest coma arm

The post-arrest coma arm used the 607 public patients of I-CARE version 2.1 [19], comatose adults after cardiac arrest at five hospitals; 3 were used for development and excluded. Each patient contributed one continuous 30-minute window chosen without reference to outcome: in the EEG segment whose start was nearest 24 hours after return of spontaneous circulation, within 12 to 36 hours, the first 30-minute stretch free of recording padding (an exact-zero second, or every channel below 2 μV for 60 s or more) that passed the clinical EEG noise-floor rule; if none passed, the next-nearest segment within 12 to 36 hours was used, and otherwise the patient was excluded with the reason recorded. The files differ by hospital (200 to 2,048 Hz, 19 to 21 channels). The 19 sites of the 10-20 montage were read by name over the needed bytes, resampled to 128 Hz and re-referenced to the per-sample median across channels, because some recordings use C3 as the recording reference. Each of the 11 analyses ran with the overlays and call rules used on clinical EEG, and its randomization-based version ran on the same windows. False-call rates use each patient's first overlay.

Background was graded by American Clinical Neurophysiology Society continuity [e22] on 18 longitudinal bipolar derivations filtered at 0.5 to 30 Hz. A 1-s epoch was suppressed when the median amplitude across derivations was below 10 μV; the grade was continuous (below 10% suppressed epochs), discontinuous (10% to 49%), burst suppression (50% to 99%) or suppressed (above 99%). A periodic-pattern flag (the EEG foundation model's periodic label in at least 10% of the window) and the log median amplitude of non-suppressed epochs were also recorded. The foundation model is not validated in post-arrest coma, so its labels entered only a sensitivity analysis.

The planned test related the mechanism measure X (Supplementary Methods 11), computed on a fixed 12-block reference schedule from 8 to 30 Hz power at C3 and C4 and taken as the median over consecutive 8-minute windows, to good outcome. Good outcome was a Cerebral Performance Category of 1 or 2, assessed by the contributing centers at 3 to 6 months [19]. The logistic model adjusted for the background grade, the periodic flag, the amplitude, hours from return of circulation to the window, age, out-of-hospital arrest, shockable rhythm, temperature target (33 °C, 36 °C or unknown) and hospital, with a weak L2 penalty. The odds ratio per SD of X carries a patient-bootstrap 95% CI (2,000 resamples) and a two-sided test at $\alpha = .05$. Patients without complete covariates or without a value of X were left out of the model and compared with those included on outcome (Fisher exact test). Secondary estimates were the change in leave-one-hospital-out AUROC when X was added to the background-only model; the call of each analysis's a priori default reading against outcome, with the same adjustment and never ranked; leave-one-hospital-out estimates; and sensitivity analyses excluding the 40 patients used in an earlier null set, replacing the continuity grade by the foundation model's background labels, and excluding patients with any flat channel. The calibration of the clinical EEG mechanism model in coma was also estimated. Outcome and covariate files stayed on one local computer; the windows were processed on a separate compute server without them.

The stated-rule readings (Supplementary Methods 3 and 4) were applied to the post-arrest rows and use the 490 patients of the outcome model, the denominators of the rates of the originally planned readings. The 30-minute window holds only shortened-to-fit versions of the Cruse 2011, Cruse 2012 and Chennu 2013 schedules, so these analyses have no stated-rule reading in post-arrest coma. Their full-length readings were run later on longer windows (Supplementary Methods 18). Post-arrest coma data were used for estimation only.

#### Supplementary Methods 15. Replication set

After every reading, both corrections of Supplementary Methods 12 and the selection rule of Supplementary Methods 4 had been applied to the development patients, the primary reading of each command-following analysis whose schedule fits a 30-minute window (Claassen 2019, Goldfine 2011, du Bois 2026, Edlow 2017, Guger 2017, Gibson 2016 and Guger 2018) was fixed, together with the tests below, in a record committed to the code repository before any replication patient was listed. The record holds each reading, its development rates, the checksum of the patient listing, the random seed and the code version. Cruse 2011 and Cruse 2012 were not replicated: their primary readings need 2,186 and 1,891 seconds of clean EEG, more than a 30-minute window. Chennu 2013 had no eligible reading.

Patients were drawn from the same seeded listing of the SpikeNet 2.0 corpus as the 400-patient set of Supplementary Methods 11, continuing in rank order after it. Any patient analyzed in this study before was refused. Each patient contributed the first recording segment holding a clean 30-minute window, at a start drawn uniformly from its eligible starts with a rank-specific seed, until 730 patients were listed (listing ranks 1,129 to 3,246). The 30-minute windows were extracted on a compute server holding the corpus and copied to the analysis computer with matching checksums. Background labels (Supplementary Methods 5) were not computed for the replication windows: the background model would have needed about 30 hours on the processors available, and no call uses the labels. The replication therefore has no background strata.

Each analysis ran on the first overlay of each window at its fixed reading, with the trial schedule placed by the same planner as in the development run, and without randomization-based rows. The Claassen 2019 development windows were the exception: they came from that analysis's own runner, with its own seeded window draw, whereas its replication windows used the common planner. Windows with an error, no fitting overlay or no usable trial left the denominator. Guger 2018 also ran its second run, for the published two-run rule. The du Bois 2026 permutation reading ran beside its primary reading, for the decision pathway (Table S18).

Tests, fixed before the draw. The 3 analyses above 5% in development (Goldfine 2011, du Bois 2026 and Guger 2018) were each tested for a first-overlay rate above 5% by a one-sided exact binomial test, with Holm adjustment over the 3. The other 4 were tested for a rate below 10% (one-sided exact binomial, Holm over the 4). A rate was called reproduced when it lay inside the 95% prediction interval around the development cross-fitted rate, $p \pm 1.96 \times \sqrt{p(1 - p)(1/N + 1/M)}$, where p is the development cross-fitted rate and N and M are the development and replication denominators. No reading was chosen on these patients. The replication is internal: it uses the same corpus and sampling frame as the development set, and recording site was not available to stratify by.

#### Supplementary Methods 16. Independent implementation, repeated recordings, outcome exposure, published cohorts and patient positive controls

Independent implementation. The du Bois 2026 and Guger 2018 analyses were written a second time from the publication texts alone [30-32] in a separate session of a large language model (October 22, 2026) given only the publication texts and no access to this study's code. The fixed elements of its instructions were: implement each analysis from the named publications only; consult nothing else; return a call, the statistic and its P value through a stated function interface; and record every choice the texts leave open with the quotation it rests on (Table S24B in Supplementary Data S2). The instructions named each call criterion (du Bois 2026: peak decoding accuracy above 70%, or task above baseline by a one-tailed paired t test; Guger 2018: accuracy above 23%); the instruction is summarized in Table S29 in Supplementary Data S2. Choices the texts left open were made independently and recorded with the text each was based on. Both versions were run on the same 200 clinical EEG development windows (a seeded sample of development patients with a stored primary du Bois 2026 row) with the same sham schedules as this study's primary readings. The Guger 2018 schedule switches the target wrist between instructions; each stimulus was passed as target, non-target wrist or distractor, without changing the independent method. Bars, set before the run: the independent rate inside the range of readings consistent with the publication (Table S21), and call agreement kappa of at least 0.6 (Table S24).

Repeated recordings. The Claassen 2019 group assesses patients with 1 to 8 recordings (mean 2.3) and calls a patient positive when the Benjamini-Hochberg procedure at q = .05 across the patient's recordings rejects a recording with AUC above 0.5 [1]; Edlow 2017 was evaluated for any positive among 1 to 5 sessions. The clinical EEG files carry no dates, and the post-arrest windows are one per patient, so each additional recording was another patient's first window. For the any-positive rule of Edlow 2017 this gives an upper bound under non-negative association of calls within a patient. For the Claassen 2019 rule it does not: the correction across recordings depends on their number, and identical recordings would give the 1-recording rate at every number of recordings (Table S25 in Supplementary Data S2).

Outcome exposure. In post-arrest coma, the share of patients with poor outcome (Cerebral Performance Category 3 to 5) called positive was computed for each primary reading, and for any of the 3 analyses above 5% among patients assessed by all 3 (Table S26).

Published cohorts. Published positive rates with delivered commands were compared with the rate of the same rule on recordings without commands, at the published number of assessments: du Bois 2026 advancement after Phase I (either of 2 sessions) by diagnosis, against the two-session criterion on clinical EEG (primary) and on prolonged-DoC EEG (secondary); and Claassen 2019 EEG brain activation (16 of 104), against the 2-recording rate of the Claassen 2019 rule assuming independent recordings, in post-arrest coma (primary) and on clinical EEG (secondary). One-sided exact tests: Fisher for du Bois 2026, comparing two counts; binomial for Claassen, against an estimated rate, so P is also given at the ends of that rate's 95% CI and against the 1-recording rate (identical recordings). Holm adjustment over the 4 primary comparisons (Table S27). The published counts were taken verbatim from the publications. No open EEG dataset with delivered commands in disorders of consciousness was available; the one found, from the du Bois 2026 group, is access-restricted.

Stroke positive control. Open stroke motor imagery data were screened for a positive control in brain-injured patients who were instructed to imagine movements. Each candidate set was first checked on a few patients. The lower-limb set (27 patients) was set aside because its idle trials desynchronized like imagery trials. The acute hand set (Liu et al.,[38] 50 patients 1 to 30 days after stroke, 40 left and right hand imagery trials) passed a check that occipital channels did not decode better than sensorimotor channels, because a hand-specific video played during imagery. No published schedule fits 40 trials, so only Guger 2017 was run, on each patient's own trials (40-trial variant), with 20 label permutations per patient as the false-call control (Table S28). The dataset's authors reported a mean accuracy of 55.57% with the same method (common spatial patterns with linear discriminant analysis, 8 to 30 Hz, 0 to 4 s, 10-fold cross-validation, all channels); here it was 54.6% on sensorimotor channels and 52.2% on all channels. Their best method, which chose the time window and frequency band by search, reported 72.21%, and they found the expected contralateral power decrease at C3 and C4 in 23 of 50 patients.[38] Task performance was not verified, so the rate with true labels is the share called positive after the instruction, not sensitivity in verified responders.

P300 positive control in amyotrophic lateral sclerosis. Visual P300 speller recordings of 48 patients with amyotrophic lateral sclerosis (bigP3BCI cohorts B, F, L and N; CC BY 4.0)[e23] were screened as a positive control for the Guger 2018 rule, using groups of real consecutive flashes so that no session was assembled. A check fixed before the analysis required a target-minus-non-target difference at Pz from 250 to 600 ms with $P < .05$ (1,000 permutations of the target flags within each sequence; non-targets at least 1 s from any target) in at least 34 of 48 patients and a positive grand average. Results are in Supplementary Results 12.

#### Supplementary Methods 17. Genuine tasks, sham labels, the du Bois 2026 implementations and calibration within each design

These analyses were added after the analyses above were complete, to ask whether the 3 analyses with excess calls detect genuine responses, where their excess calls come from and how much detection remains at a patient-level false-call rate set within each design. For the tactile and stroke recordings, the code, the sham construction, the readings and the pass criteria were committed before any analysis recording was read; synthetic recordings and one development participant or patient per dataset were used only to build and test the code. The du Bois 2026 and Goldfine 2011 analyses of development windows reused windows already analyzed (the independent du Bois 2026 rate of 96.0% was known, Table S24), and the calibration within each design reused stored rows; their readings were fixed in code before they were run. A code review during the first runs found that three sham constructions were not valid nulls (a stroke sham that split whole rest periods across a run and so broke the within-trial pairing that cancels slow drift; tactile shams and a tactile randomization null that let a finger repeat within a run, so that finger-specific evoked responses became decodable; and a per-trial coin-flip label exchange that added drift). Those runs were stopped, their rows discarded unread, and the runs restarted with the constructions described below.

Tactile P300 (Table S32). Open Braille brain-computer interface recordings (G-Node, CC0 1.0) of 20 blind or sighted adults, each with two sessions (large and small pins), 45 scalp channels at 500 Hz with linked-ear reference.[36] One finger rests on each of 8 Braille cells; each selection cycle raises every cell 10 times in random order (stimulus onset asynchrony about 300 ms) while the participant counts the target cell's activations, 16 cycles per session. Guger 2018 ran unchanged on its own 8-stimulus layout: within a cycle, the k-th touch of each finger forms one group of 8 stimuli with exactly one on the attended finger, so the analysis's 8-way decision is the recording's own question of which finger was attended. Run A is cycles 1 to 6 (60 groups, the published run length) and run B cycles 7 to 12; a session is called if either run is, the per-patient rule of the replication. True: the attended finger as the label. Sham: each cycle's target mapped through one random derangement of the 8 fingers per session, so the sham label is never the attended finger and a run's sham targets are distinct fingers, as the true ones are. Readings: the primary reading (median split-half accuracy above 23%), the default reading, the independent implementation and a randomization test (the analysis's LDA score margin against the run's targets permuted across its cycles, 200 draws). Bars: the positive control passes if the primary reading's exact upper 95% limit for true-label detection reaches this analysis's bar for healthy detection (the published 3 of 3, Table S2) and true-label calls exceed sham-label calls by an exact McNemar test ($P < .05$); the sham rate was tested against 5% (one-sided exact binomial) and reported beside its split by whether the sham label mostly stayed on the attended hand. One participant (sub-6) was used only to build the loader. Session CIs here are exact binomial and do not account for the 2 sessions of each participant; participant-cluster CIs are in Table S36. Fingertip pins differ from the vendor's wrist and back vibrators.

Stroke motor imagery (Table S33). Open recordings of 24 patients 3 to 14 months after stroke, each at 3 time points (pre, post and follow-up; 5 patients have no follow-up) with 4 runs per session, 59 EEG channels at 250 Hz.[37] Each trial is 7 s of rest with a fixation cross, 1 s of preparation and 6 s of imagery of the affected hand or of a robotic sixth finger, the imagery type in random order; a standard run has 20 trials. Unit: the session (patient x time point), with patients clustered in every CI (patient-cluster bootstrap, 2,000 draws). Goldfine 2011: per trial, two 3-s epochs from the imagery period (0.5 to 6.5 s after the cue) as task and two from the preceding rest period (0.5 to 6.5 s after rest onset) as rest, through the analysis's own feature code with its timing set for this design, run by run, and the primary decision applied to the merged runs. Sham: rest periods only, each period's two epochs labeled task and rest in random order, with half of each run's rest periods in each order. Exchanged: the recorded trials with task and rest labels swapped in exactly half of each run's trials, a conservative comparator that keeps imagery in both classes and does not keep the recording's cancellation of slow drift, because rest always precedes imagery. du Bois 2026: affected-hand against sixth-finger imagery, cue onsets as recorded, the session's runs demeaned and joined with 0.25-s equal-power cross-fades; the second implementation (Supplementary Methods 16) and our implementation with random folds, which are legitimate here because the imagery type was randomized; sham: the trial labels redrawn at random, balanced. Twenty draws of each null per session. Readings: detection is the share of sessions called with true labels and the null rate the mean over sessions of the share of draws called; a reading separates true from null labels if the lower 95% limit of their difference is above 0, and a Goldfine 2011 reading is calibrated if the upper 95% limit of its rest-only sham rate is at most 10%. Byte-identical sessions, found by file hash, were excluded when the rows were merged, before any merged result was read: two sessions of different patients (both excluded) and one repeated session of one patient (the later excluded). Trials without room for the windows were dropped. One patient was used only to build the loader. Task performance was not verified.

The second du Bois 2026 implementation (Table S30). The independent implementation of Supplementary Methods 16 follows more of the choices the publication states (3 filter pairs per band, mutual information with 3 quantization levels, a 40-ms step, accuracy at each time point and the raw-curve peak refinement; Table S4 lists where our implementation departs from them); it uses every recorded channel and random stratified folds, a rule the publication does not state. It was moved toward our primary reading one choice at a time by changing its settings for the duration of a call, never its code: first the fold rule (leave one block out, the six blocks of the run) and then, cumulatively, our channel set, our peak rule, a 100-ms step, our band edges, 2 filter pairs, Gaussian mutual information and Ledoit-Wolf shrinkage; each change was also applied alone. A factorial crossed folds (random or block-wise), peak rule (the published refinement or ours) and configuration (the 84-configuration search or one configuration fixed before any window was run). The publication's 100-permutation evaluation was run two ways: literally, with the configuration selected on the true labels held fixed and P computed at the selected peak time, and with the whole 84-configuration search and peak selection repeated inside each permutation, so that a selected maximum is compared with selected maxima; P = (number of permuted accuracies at or above the observed + 1) / 101, an unusable permuted draw counting as at or above. Random folds were redrawn in each permutation; block-wise folds stay the blocks. Units: the 200 development windows of the independent implementation (Supplementary Methods 16), first overlay. The repeated-search version, about 40 processor-hours for 200 windows, ran first on the first 60 of these windows, a subset fixed before the run, and was extended to the other 140 after those finished and before any of them ran; both are reported. Synthetic checks used 30 command-free 1/f recordings per drift level (Supplementary Methods 10, slow amplitude drift added).

The Goldfine 2011 variance (Table S31). The primary reading's jackknife deletes one tapered estimate at a time (5 tapers x 3-s epochs) and treats the estimates as independent. Estimates from one command share its slow fluctuations, so the command-level jackknife deletes all estimates of one command at a time; its variance replaced the sample-level one in an otherwise unchanged decision, with a normal or t reference (degrees of freedom: the commands of both conditions - 2), and with the frequency grid decimated to one bin per resolution element (2 Hz at time-bandwidth 3 on 3-s epochs, the width of the >2 Hz contiguity criterion). Variance inflation: the command-level jackknife variance of the standardized spectral difference, whose expected value for independent estimates is 1, the median over channels of the band mean. Units: every development window with a stored first-overlay primary Goldfine 2011 row whose recording is held locally (342 windows; the primary reading was rerun and compared with the stored call). Bars: a reading is calibrated if its upper 95% limit is at most 10%, and dependence is the mechanism if the median variance inflation is at least 2.

Calibration within each design (Table S34). Each analysis's evidence statistic, the one its size correction uses (Supplementary Methods 8), was thresholded on recordings without commands from the same design, targeting a 5% chance that a patient assessed k times is called positive by chance: per-assessment level 1 - $0.95^{(1/k)}$ (exact for independent assessments; positive association between a patient's assessments lowers the patient-level rate, but sampling error in a threshold at a small level can raise it, so Table S38 measures it where repeated runs exist), the threshold the (1 - level) quantile of the null statistic and a call a statistic strictly above it. Thresholds were cross-fitted by patient: the null was split into halves by a hash of the patient identifier, each half's false calls were judged by the other half's threshold, and each genuine-task unit was judged by the threshold of the half its own participant or patient does not fall in under the same hash. A threshold is never carried across designs, because the statistic's scale depends on trial count, runs and channels. Designs: the published schedules, with the clinical EEG development windows as null and healthy volunteers as genuine responders (motor imagery: the sessions of the original assembly of Supplementary Methods 9, the only volunteer rows that carry the fixed readings' statistics, because the one-source rebuild ran the randomization-based variants only; auditory counting for Guger 2018); the tactile runs, with sham-labeled runs as null; and the stroke sessions, with each analysis's sham as null. Missing statistics left the denominator; infinite ones were capped. A null of n units cannot place a threshold below its largest value, so small nulls (76 tactile runs) cannot reach the lowest levels; the achieved held-out false calls, pooled over the two halves, are reported beside each detection with 95% CIs from a bootstrap of participants or patients (2,000 resamples; in each, both thresholds were re-estimated on the resampled rows and a participant drawn more than once stayed in its half); a cell whose achieved rate is above its target is marked, descriptively, as is a cell whose target is finer than half the null can resolve. These intervals reflect runs and sessions clustered within participants (76 tactile runs from 19 participants; 1,220 stroke null draws from 23 patients) and the estimation of each threshold; achieved rates were not tested against their targets. k = 1, 2 (a median of 2 recordings per patient in acute intensive care[3]), 5 (repeated assessment in prolonged disorders of consciousness[41]) and 10 (daily sessions[32^]). Beside each: detection by the published rule at its own false-call rate and by the randomization-based version at P at or below the per-assessment level.

Frozen test of the command-level Goldfine 2011 variance (Table S35). After the development results above, the command-level jackknife (normal reference) was fixed as the primary reading, with the t reference and the decimated grid as secondary readings, and the test fixed as a one-sided exact binomial of the count against 5% (rate below 5%), Holm over the replication and post-arrest sets, calibrated if the Holm-adjusted P is below .05. The frozen record and code were committed before any replication or post-arrest window was run. Two later code changes leave the test unchanged: windows without usable trials are now counted under that name instead of as not reproduced (the same windows leave the denominator), and the development strata use the window's MORGOTH background without the activation filter of Supplementary Methods 5, which needs another pipeline. Each unit is the patient's first published Goldfine 2011 overlay: the 730 replication windows (Supplementary Methods 15) and the 544 post-arrest windows

(Supplementary Methods 14), rebuilt from the kept windows with the arm's own padding context. On the same features, the published (sample-level) reading had to reproduce the stored call of that window; a window that did not would have left every denominator. McNemar compared the published and command-level readings on the same windows. Strata, descriptive: the post-arrest ACNS continuity grade and the development windows' MORGOTH background.

Time-matched specificity (Table S37). In the acute-stroke recordings of Table S28 (47 distinct patients), left- and right-hand imagery were contrasted at the same time after the cue: one 3-s epoch from 0.5 to 3.5 s after imagery onset, with the Goldfine 2011 features, Laplacian and decision unchanged and 10 s of reflected signal added at each end of the recording (the first cue lies about 2 s into each file, inside the window margin the features need). The cues are hand-specific videos, so the contrast is matched in time, not in stimulus. The 40 strictly alternating trials formed 4 pseudo-runs of 10. Null: the labels of exactly half of the 20 consecutive left / right pairs swapped, 20 draws per patient (seeded). Readouts and the 'sensorimotor-specific' rule were committed before any patient was run.

Cluster-aware inference (Table S36). Stroke rates were resampled by patient and tactile rates by participant (2,000 resamples, percentile intervals); the change in stroke detection between the published and command-level Goldfine 2011 jackknife was computed on the same sessions; a sign test over participants compared true and sham sessions called. These descriptive summaries were computed once from the stored rows.

Direct patient-level calibration (Table S38). Where the clinical null had repeated runs per patient (Guger 2018, up to 10 runs; du Bois 2026, 2 for 95 patients), each half's threshold was set on its patients' maximum statistic over the first k runs, so that the patient-level false-call rate is calibrated without assuming independent assessments, and measured on the other half; the independence-based threshold of Table S34 was measured the same way. The runs are successive windows of one recording, not separate days.

#### Supplementary Methods 18. Full-length schedules and Claassen 2019 on six sites

Three schedules are longer than the 30-minute windows of the replication set and the post-arrest arm: Chennu 2013 at its stated 20 blocks (2,530 s), the primary 4-block reading of Cruse 2011 (2,166 s) and the primary reading of Cruse 2012 (1,871 s); each window also carries a 10-s lead and tail. In development, Cruse 2011 and Cruse 2012 ran on the clinical recordings long enough to hold them, and Chennu 2013 only as a 12-block twin, never eligible for the primary reading (Table S8A). After the other results had been reported, each schedule was run at full length on one longer window per patient. The readings were fixed and committed before any longer window was read: the development primary readings of Cruse 2011 and Cruse 2012 and, for Chennu 2013, the reading chosen by the rule of Supplementary Methods 4 on the development recordings that hold 20 blocks in a clean stretch. Of 364 development recordings at least 2,550 s long, 89 held such a stretch; padding inside the others broke every stretch of that length. Both eligible 20-block readings ran on all 89, and the one with fewer calls was chosen. Each run used the benchmark engine unchanged, on the first overlay. These rates are descriptive and outside the Holm families; P values are one-sided exact binomial tests against 5%.

Replication set. In each patient's listed segment, the clean stretches found when the 30-minute windows were placed (the rule of Supplementary Methods 5, second by second) were searched for the longest need they hold (2,550, 2,186 or 1,891 s), and the start was drawn uniformly from the eligible starts with a seed for each patient. Of the 730 patients, 540 held one (231 at 2,550 s, 167 at 2,186 s and 142 at 1,891 s); the listing was hashed before any window was read. Each window was read from the corpus over a read-only connection and checked again against the clean rule on the whole recording; where it overlapped the stored 30-minute window, the samples were identical. Sokoliuk 2021 (1,347 s) fits 30 minutes and ran on the replication set's own windows.

Post-arrest arm. In the segment each patient's 30-minute window came from, the arm's own selection (Supplementary Methods 14), unchanged except for the window length, chose the first clean window of 2,550 s, else 2,186 s, else 1,891 s. Segments were read from PhysioNet by byte range, in patient-identifier order, for the 236 patients whose files could be obtained (identifiers 0284 to 0621); 231 held a long clean window and 5 did not. The other 308 patients were not read; Table S39C compares the hospitals of the two groups. The 30-minute windows of all 544 patients were read earlier (Supplementary Methods 14).

Claassen 2019 on six sites. The prolonged-DoC files carry 6 scalp channels (F3, F4, C3, C4, O1 and O2), and every other whole-head analysis ran on these 6 sites there (Supplementary Methods 1). The released Claassen 2019 pipeline uses 19 sites. It was run on the 6 with two module settings changed for the run: the channel list, and the spherical-spline current source density matrices rebuilt for the 6 sites with the pipeline's own routine (24 features instead of 76). The classifier, leave-one-trial-out cross-validation, permutation test, early stopping and call were unchanged. The 13 missing sites were not interpolated, because an interpolated channel is a weighted sum of recorded channels and adds no information. To measure what the reduced montage alone changes, the 6-site pipeline also ran on the 481 clinical EEG recordings of the Claassen 2019 development analysis at the same window starts, schedules and permutation seeds as the 19-site results, for the published test and the randomization-based test; the pairs differ only in montage and were compared with exact McNemar tests. On the 38 prolonged-DoC patients it ran as the other analyses did there: first overlay, with its randomization-based version. Its white-noise rate is that of its one reading, measured on the 19-site white noise of Supplementary Methods 10.

### Supplementary Results

#### Supplementary Results 1. Sensory responses read as volition

Passive recordings contain real stimuli and no task intent, so an ERP analysis can call a sensory response a response to a command. These calls form a separate family. They are not pooled with the false calls on recordings without stimuli, and the family has no randomization-based arm, because a label permutation across physically different stimuli is not a volition null. The participants were conscious healthy adults, so this measures responses without task intent, not without consciousness, and healthy responses are larger than those of most patients. Each analysis was run on its planned lowest-rate reading; the stated-rule framework was not applied to this family, so some readings (for example the Gibson 2016 P3a and steady-state contrasts) are ones that the top-down convention classes as contradicting. The reading used in each row is given in Table S5.

On the ERP CORE mismatch negativity data (38 participants), the Gibson 2016 reading called 23.7% of participants (95% CI, 10.5%-36.8%) and the Guger 2018 reading 34.2% (95% CI, 18.4%-50.0%), both above 5% after Holm adjustment; Chennu 2013 called 2.6% (95% CI, 0.0%-7.9%). On the unattended stream of the Kojima recordings (13 participants), the Guger 2018 reading called 17.9% of runs (95% CI, 10.3%-25.6%), above 5% after Holm adjustment, and Chennu 2013 and Gibson 2016 called 1.3% and 2.6%. On ds003061 (11 participants) no analysis was above 5% (Chennu 2013, 9.4%; Guger 2018, 16.7%; Gibson 2016 steady-state test, 0 of 10 runs in 5 participants). As a construction check, sham instructions drawn independently of the EEG gave per-participant rates of 0.0% to 15.4% across the nine cells, none significantly above 5% (smallest P = .14, Guger 2018 on the Kojima recordings, 2 of 13 participants). That check is exact by construction because stimulation is identical across the sham conditions; it shows that the machinery is exact and does not test real attended-versus-ignored effects, which need instructed data.

#### Supplementary Results 2. Cohorts and false calls by analysis in clinical EEG

The clinical EEG analysis set comprised 974 patients with one recording each (median analyzed duration, 30 minutes; IQR, 30-53 minutes; Table S6). In the first analysis window the model-assigned background was normal awake in 27% to 34% of recordings, sleep in 42% to 47% and ICU-typical in 4% or fewer, depending on the analysis. Denominators differed by analysis because each schedule needs a clean window of its own length, from 147 patients for the four-block Cruse 2011 schedule to 850 for Guger 2018. The prolonged-DoC set comprised 38 patients (28 VS/UWS, 10 MCS) with overnight recordings (median, 6.0 hours). Among the 30 with linked clinical data, the median age was 42 years (IQR, 37-57), 14 (47%) were women, the etiology was traumatic in 14, anoxic in 10, vascular in 5 and inflammatory in 1, the median time since injury was 4 months (IQR, 3-5), and the median Coma Recovery Scale-Revised total score was 6.5 (IQR, 5-8). The 490 post-arrest patients with complete data came from 5 hospitals, with windows near 24 hours after return of circulation (median age, 62 years; IQR, 52-72; 148 women, 30%; 382 out-of-hospital arrests; 258 shockable rhythms), a continuous background in 27%, discontinuous in 22%, burst suppression in 36% and suppression in 16%; 185 (37.8%) had a good outcome (Supplementary Results 10).

Under the primary reading Goldfine 2011 called 164 of 703 patients (23.3%; 95% CI, 20.2%-26.6%), du Bois 2026 157 of 707 (22.2%; 95% CI, 19.2%-25.5%) and Guger 2018 117 of 850 (13.8%; 95% CI, 11.5%-16.3%), each with a Holm-adjusted P below .001. Claassen 2019 called 33 of 481 (6.9%; 95% CI, 4.8%-9.5%; Holm-adjusted P = .22). The other analyses called Gibson 2016 25 of 683 (3.7%; 95% CI, 2.4%-5.4%), Cruse 2012 5 of 210 (2.4%; 95% CI, 0.8%-5.5%), Guger 2017 8 of 550 (1.5%; 95% CI, 0.6%-2.8%), Cruse 2011 1 of 147 (0.7%; 95% CI, 0.0%-3.7%) and Edlow 2017 3 of 796 (0.4%; 95% CI, 0.1%-1.1%). Chennu 2013's stated 20 blocks need 2,530 s, more than the analysis windows held; on the 89 development recordings that hold them in a clean stretch, its chosen reading called 0 (0.0%; 95% CI, 0.0%-4.1%) and the other eligible reading 2 (2.2%; 95% CI, 0.3%-7.9%) (Supplementary Methods 18; Table S39A), and its shortened 12-block twin called 2 of 467 (0.4%; 95% CI, 0.1%-1.5%) (Table S8A). Cross-fitted rates differed from the selected rates by 1.6 percentage points or less in every clinical EEG analysis (Table S7A).

Under the originally planned reading only Goldfine 2011 exceeded 5% after Holm adjustment (268 of 703; 38.1%; 95% CI, 34.5%-41.8%). The originally planned du Bois 2026 rate (36 of 707; 5.1%; 95% CI, 3.6%-7.0%) came from a reading whose call is a label permutation, a criterion the publication does not state, and the originally planned Guger 2018 rate (40 of 838; 4.8%; 95% CI, 3.4%-6.4%) from a 2-stimulus paradigm that contradicts the publications. The a priori default readings gave 98.2% for Goldfine 2011 (a liberal departure from the publication), 90.0% for du Bois 2026 (liberal departure), 30.4% for Edlow 2017 and 17.3% for Guger 2018 (faithful literal readings of unstated items) (Table S7B in Supplementary Data S2). The fidelity grade is descriptive and no rate of a liberal departure is called the published method's rate.

The covert-processing analysis called 41 of 565 recordings (7.3%; 95% CI, 5.3%-9.7%) by its surrogate test, with a cross-fitted rate of 8.1% (95% CI, 5.1%-11.2%). Run later on the replication set's windows, it called 58 of 693 (8.4%; 95% CI, 6.4%-10.7%; one-sided $P < .001$ against 5%) (Table S39A). Its published prognostic cut-off (inter-trial phase coherence of at least 0.116) was exceeded in 136 of 595 recordings (22.9%; 95% CI, 19.5%-26.4%); that cut-off was published as a prognostic threshold and is not a per-recording call rule, so this figure is described only as the share of no-stimulus recordings that exceed it (Table S8B). Rates of covert-processing analyses are never pooled with those of command-following analyses.

#### Supplementary Results 3. Per-patient rules over repeated assessments

Under the Guger 2018's two-run rule, 206 of 841 patients (24.5%; 95% CI, 21.6%-27.5%) had at least 1 positive run; 42 patients had fewer than 2 usable runs. When any positive run counted, the share of patients with at least that many runs rose from 13.8% with 1 run (883 patients; 95% CI, 11.6%-16.3%) to 36.1% with 3 (790 patients; 95% CI, 32.7%-39.5%), 52.6% with 5 (698 patients; 95% CI, 48.8%-56.3%) and 62.9% with 7 runs, the most per patient reported in the publications (639 patients; 95% CI, 59.0%-66.7%); with 10 runs, beyond the published range, 75.2% (266 patients) (Table S9C). In a fixed cohort of the 639 patients with at least 7 runs, the share was 14.1%, 24.4%, 35.8%, 51.8% and 62.9% with 1, 2, 3, 5 and 7 runs, so the rise does not come from a changing population. The du Bois 2026 criterion for advancing to further training was met by 30 of the 95 patients with 2 eligible windows (31.6%; 95% CI, 22.4%-41.9%); 612 patients had fewer than 2 eligible windows. The Claassen 2019 two-recording rule called between 7.1% (1 of 14; 95% CI, 0.2%-33.9%), for two windows of one session, and 8.6% (95% CI, 5.4%-12.2%), for two independent recordings (Table S9B). The k = 1 rates of the other analyses equal their primary rates (the Guger 2017 composite uses 559 patients with at least one usable window, against 550 with a first-window result; 8 patients called in both).

In the 38 prolonged-DoC patients the du Bois 2026 criterion was met by 18 (47.4%; 95% CI, 31.0%-64.2%) and the two-run Guger 2018 rule by 8 (21.1%; 95% CI, 9.6%-37.3%). Any positive run among 1, 3, 5 or 7 runs called 10.5%, 31.6%, 47.4% and 60.5% of the 38 patients, and with 10, 20 and 40 runs, beyond the published range, 71.1%, 84.2% and 100.0% (37 patients at 40 runs). Under the generous reading the du Bois 2026 two-session criterion was met by 9 of 95 clinical EEG patients (9.5%), because the generous reading uses the permutation call; under the strict reading the du Bois 2026 and vibrotactile results equal those of the primary reading (Table S9D).

#### Supplementary Results 4. Prolonged disorders of consciousness and EEG background

In the 38 patients with prolonged DoC du Bois 2026 called 9 (23.7%; 95% CI, 11.4%-40.2%), Guger 2018 called 4 of 36 (11.1%; 95% CI, 3.1%-26.1%) and Goldfine 2011 called 3 of 38 (7.9%; 95% CI, 1.7%-21.4%). With the reading selected on 19 patients per half, the cross-fitted estimates were 18.4% (95% CI, 0.0%-34.2%)

for Goldfine 2011 and 25.0% (95% CI, 2.3%-39.6%) for Guger 2018. Gibson 2016 called 2 of 38 (5.3%), the covert-processing analysis 1 of 37 (2.7%), and the remaining analyses 0 of 36 to 38 (upper 95% bounds, 9.3% to 9.7%). Claassen 2019, run later on the 6 recorded sites, called 1 of 38 (2.6%; 95% CI, 0.1%-13.8%) and its randomization-based version none (Supplementary Results 14). BDSP rates are not DoC false-positive rates: the model fitted to clinical EEG over-predicted calls in the DoC recordings (calibration intercept -0.58; Table S15A in Supplementary Data S2), and the six-channel montage and overnight recordings differ from the clinical EEG.

False calls were not confined to one background (Table S10 in Supplementary Data S2). In model-assigned normal awake recordings Goldfine 2011 called 34 of 177 (19.2%; 95% CI, 13.7%-25.8%), du Bois 2026 39 of 179 (21.8%; 95% CI, 16.0%-28.6%) and Guger 2018 37 of 273 (13.6%; 95% CI, 9.7%-18.2%); their randomization-based versions called 7 of 177 (4.0%), 7 of 179 (3.9%) and 15 of 273 (5.5%) of the same recordings. In ICU-typical backgrounds the counts were 2 of 25, 6 of 24 and 2 of 29, and in the DoC wake-like windows 0 of 19, 6 of 20 and 3 of 16. These windows are those with automated activation-procedure exclusion (Supplementary Methods 5) and without a transitioning flag. The recordings are not from a known care setting: a model-assigned background is a proxy for the pattern of the EEG, and these rates are not rates in patients of any setting.

#### Supplementary Results 5. Synthetic noise and below-chance accuracy

On white noise the du Bois 2026 corrected reading called 20 of 60 recordings (33.3%; 95% CI, 21.7%-46.7%) and Guger 2018 9 of 60 (15.0%; 95% CI, 7.1%-

26.6%). Goldfine 2011 called none of 60 (95% CI, 0.0%-6.0%) but called 2.3%, 18.3% and 24.3% of 300 synthetic recordings at amplitude-fluctuation log-scale SD 0.25, 0.5 and 0.75, so its calls rise as the amplitude envelope fluctuates more. Claassen 2019 called 3 of 60 (5.0%), Cruse 2012 9 of 60 (15.0%; 95% CI, 7.1%-26.6%), and the other primary readings selected on clinical EEG 0 or 1 of 60 (Cruse 2011, 1; Edlow 2017, 0; motor imagery, 0; Chennu 2013, 0; Gibson 2016, 1) and 2 of 60 for the covert-processing analysis. The readings selected on the prolonged-DoC data called 11 of 60 (18.3%; 95% CI, 9.5%-30.4%) for Guger 2018 (Annen 2018 timing), 6 of 60 for Cruse 2012 (contiguous groups), 3 of 60 for Edlow 2017, 2 of 60 for Gibson 2016, 4 of 60 for the covert-processing analysis and 0 of 60 for Chennu 2013, Cruse 2011 and the motor imagery analysis. Of the 68 eligible readings, 13 called at least half of 60 white-noise recordings: 9 du Bois 2026 readings, 2 motor-imagery readings with common spatial patterns fitted once on all trials, and 2 Guger 2018 readings that summarize the accuracy curve by its maximum (Table S11A in Supplementary Data S2). The Cruse 2012 excess did not hold at 500 further draws: the random-group reading called 38 of 500 (7.6%; 95% CI, 5.4%-10.3%; exact P = .008 against 5%) and the contiguous-group reading 35 of 500 (7.0%; 95% CI, 4.9%-9.6%), with the same rates on pink noise (paired P = 1.00 and .82). In a diagnostic analysis the excess disappeared when the permutation null kept each cross-validation fold's class balance: on the same 353 draws the random-group reading fell from 7.1% to 3.4% (95% CI, 1.8%-5.9%; 14 against 1 discordant calls, P = .001) and the contiguous-group reading from 6.5% to 4.2% (8 against 0, P = .008) (Table S11G). The analysis's permutation null keeps the cross-validation groups fixed and permutes labels across all trials, so its permuted labellings give less class-balanced test folds than the observed labelling: the mean absolute deviation of a test fold's class share from the overall share was 0.000 for the observed labels and 0.107 for permuted labels in the random-group reading (0.071 and 0.107 in the contiguous-group reading). Its P values were accordingly not uniform under white noise: on 200 draws (400 per-comparison P values) the Kolmogorov-Smirnov D was 0.185, 8.0%, 0.75% and 0.25% of P values were below .05, .01 and .001, and observed accuracy sat 0.0097 (SE, 0.0010) above the mean of its permutation null (Table S11H). The analysis's own P, the smaller of two, fell below .05 in 15.5% of draws, which is not uniform even under a calibrated null. Artifact rejection never fired, epochs did not overlap (minimum inter-onset interval 6.00002 s against a 6-s epoch), all 180 of 180 trials were kept in every draw, and white and pink noise gave the same rates, so spectrum shape was not the cause. Cutting the noise to the clinical trial counts did not lower the rate (6.1% against 7.1%; P = .70). The clinical rate of the random-group reading (5 of 210; 2.4%) remained below its white-noise rate (Fisher P = .006), and why is not identified. The implementation bar was met for every analysis (the Guger 2018 and du Bois 2026 analyses being declared exceptions), the mechanism bar was met by 1 of 7 tested analyses (Goldfine 2011, 0, 30, 82 and 96 calls of 300 at log-scale SD 0, 0.25, 0.5 and 0.75; Holm-adjusted P < .001), the construction bar was met (no randomization-based version rose; smallest raw P = .27), and the stopping rule was not triggered, because the originally planned Goldfine 2011 reading called 112 of 600 (18.7%) at log-scale SD 0.25 to 0.5 (Tables S11D to S11H; Tables S11B and S11C in Supplementary Data S2).

Under cross-validation by whole blocks, accuracy fell below chance in most no-command recordings (Table S12 in Supplementary Data S2). The Edlow 2017 reading had a median accuracy of 0.40, with 96.7% of recordings below 0.5 (38 of 38 in DoC), and the du Bois 2026 reading had a median peak accuracy of 0.37, with 85.6% below 0.5 (73.7% in DoC). By comparison, the share below chance was 52.4% for Claassen 2019 (AUC), 52.4% for Cruse 2011, 46.9% for the motor imagery analysis and 0.5% for Cruse 2012, whose accuracy statistic is a maximum over time. Guger 2018 accuracy is eight-way (chance, 0.125), with a median of 0.10 and 64.7% of recordings below chance. Below-chance accuracy is the signature of slow drift across blocks combined with block-wise cross-validation [e24].

#### Supplementary Results 6. Healthy volunteers

On sessions assembled from healthy participants' motor imagery with one source trial per slot, the corrected Goldfine 2011 reading detected 51 of 51 participants (100%; 95% CI, 93.0%-100%) and called 14 of 51 label-shuffled sessions (27.5%; 95% CI, 15.9%-41.7%). The block-wise corrected du Bois 2026 reading detected 41 of 51 (80.4%; 95% CI, 66.9%-90.2%) and called 15 of 51 shuffled sessions (29.4%; 95% CI, 17.5%-43.8%) (Table S13A in Supplementary Data S2). The leave-block-out readings detected 50 of 51 (Edlow 2017, with permutation by 1-s column and cross-validation by whole blocks) and 41 of 51 (du Bois 2026, label-permutation call) and called none of 51 shuffled sessions (95% CI, 0.0%-7.0%); these assembled sessions contain no slow drift across blocks, so their detection is optimistic (Limitations of the main text). The randomization-based versions detected 20 of 20 (Claassen

2019), 50 of 51 (Goldfine 2011 and Edlow 2017) and 45 of 51 (du Bois 2026) participants and called 0% to 5.9% of shuffled sessions, none significantly above 5% (Table S13B in Supplementary Data S2). The motor imagery analysis detected few participants in either version (3 of 13 published, 5 of 17 randomization-based). On the auditory counting data, randomization lowered detection by the ERP analyses (Guger 2018, 4 of 13 against 10 of 13; Gibson 2016, 1 of 13 against 6 of 13; Chennu 2013, 5 of 13 against 6 of 13), and Guger 2018 called 10 of 13 label-shuffled sessions under its published default reading (Table S13C in Supplementary Data S2).

The rebuild with one source trial per slot lowered the label-shuffled false calls of the published default readings of Claassen 2019 (from 5 of 20, 25.0%, to 2 of 20, 10.0%), Edlow 2017 (from 14 of 51, 27.5%, to 3 of 51, 5.9%), the motor imagery analysis (from 6 of 15, 40.0%, to 2 of 15, 13.3%) and du Bois 2026 (from 26 of 51, 51.0%, to 18 of 51, 35.3%) and left Goldfine 2011 at 51 of 51 (Table S13D). Within-class clustering of excerpts therefore explains most of the excess shuffled calls of Claassen 2019, Edlow 2017 and the motor imagery analysis in the original assembly and does not explain those of du Bois 2026 and Goldfine 2011. On the original assembly the verdict was consistent with the published rate for five analyses (Claassen 2019, Goldfine 2011, Cruse 2012, Edlow 2017, Gibson 2016), a fidelity concern for four (Chennu 2013, du Bois 2026, the motor imagery analysis, Guger 2018), and not graded for Cruse 2011; the published default readings of 7 of 9 analyses called significantly more than 5% of shuffled sessions (all but Chennu 2013 and Gibson 2016). Among all readings of an analysis, detection ranged widely (Table S13E): for example, 15 of 16 motor-imagery readings and 110 of 120 vibrotactile readings had an upper bound below the published rate.

#### Supplementary Results 7. Randomization-based design

Randomizing trial order and testing at the trial unit gave pooled false-call rates of 3.9% to 5.5% across the 11 analyses, and no dataset, background stratum or recording unit showed a Holm-significant excess (Table S14A). Four analyses exceeded the 5% point-estimate bar (du Bois 2026, 5.1%; Edlow 2017, 5.3%; Gibson 2016, 5.3%; Chennu 2013, 5.5%), none with a significant excess (smallest Holm-adjusted P, .25 to .79). Detection of simulated responses was noninferior to the size-corrected published analysis at 17 of 34 evaluation points (Table S14B in Supplementary Data S2): Claassen 2019 2 of 4, Goldfine 2011 0 of 4, Cruse 2011 1 of 4, Cruse 2012 4 of 4, Edlow 2017 0 of 4, the motor imagery analysis 3 of 4, du Bois 2026 2 of 4, Guger 2018 2 of 2, Chennu 2013 2 of 2 and Gibson 2016 1 of 2. It was lower at every point for Goldfine 2011 (by 17.3 to 57.2 percentage points) and Edlow 2017 (by 8.4 to 19.9 points). Without size correction the Goldfine 2011 comparator detected 91.6% to 98.5% and the du Bois 2026 comparator 100.0%, against 42.0% to 82.5% and 81.1% to 99.5% after correction (Table S14C), so the uncorrected comparison is inflated.

In post-arrest coma, the randomization-based versions called 3.2% to 6.9% pooled over overlays, Goldfine 2011's 6.9% above the 5% point-estimate bar, with a Holm-significant excess for one Goldfine 2011 timing (Supplementary Results 10 and Table S16A).

#### Supplementary Results 8. Mechanism analyses

The planned alignment test of slow EEG fluctuations with the label rhythm (M1) was not supported (odds ratio per SD, 1.06; 95% CI, 0.98-1.15; one-sided P = .06); the point estimate was positive in every sensitivity analysis and ranged from 1.02 to 1.17 (Table S15B). In designs that are randomized trial by trial or otherwise unexposed, the slope of false calls on the aperiodic exponent was null (odds ratio per SD, 0.96; 95% CI, 0.90-1.02; upper bound within the 1.25 margin) and smaller than in block designs (ratio 1.18; 95% CI, 1.06-1.31; one-sided P = .002), which fits the explanation but rests on two analyses. The pre-stated prediction for deflated designs failed (pooled odds ratio per SD of the exponent, 1.21; 95% CI, 1.07-1.38), and the rates of the Claassen 2019, Goldfine 2011 and Guger 2018 default readings were above nominal. In the secondary family, concentration of false calls on a subset of patients was not rejected (Holm-adjusted P = 1.00), and neither was artifact share (odds ratio per 10 percentage points, 1.01; 95% CI, 0.99-1.03; Holm-adjusted P = .66) nor the minimally conscious versus vegetative state contrast of X (Hodges-Lehmann shift, -0.008; 95% CI, -0.244 to 0.199). Organized backgrounds carried higher false-call odds than disorganized ones (odds ratio, 1.50; 95% CI, 1.22-1.83; Holm-adjusted P = .008), unchanged after X enters the model, so X does not mediate it; in this corpus organized backgrounds include non-ICU recordings, and the background test (S2) supports no statement about patients with disorders of consciousness. The BDSP-fitted model over-predicted false calls in the prolonged-DoC recordings (calibration intercept -0.58; 95% CI, -0.80 to -0.38) and its predictions were too extreme (slope 0.67; 95% CI, 0.58-0.75). An exploratory estimate in 30 patients (10 recovered) was null for X against the Coma Recovery Scale-Revised arousal subscale (Spearman correlation, 0.06; 95% CI, -0.27 to 0.38) and for recovery (AUROC, 0.47; 95% CI, 0.25-0.68). The study was not powered for prognosis, and no inference about prognosis is drawn.

The confirmatory replication of the earlier stopping-rule test supported the spectral explanation of block-design false calls on 435 patient-disjoint recordings: in simple 8- and 12-block designs, surrogates that kept each recording's power spectrum and randomized its phases produced false calls at least as often as the recordings themselves (20.4% against 16.2%; R, 1.38; 95% CI, 1.27-1.50), and AR-prewhitening reduced the rate to 3.0% (95% CI, 2.5%-3.6%). That spectrum, and not a recording-level feature, therefore reproduces the false calls of simple block designs. The non-stationarity index did not meet its confirmation bar on 400 fresh patients (odds ratio per SD, 1.17; 95% CI, 0.99-1.37; bar, at least 1.20 with a lower bound above 1); an earlier run that omitted the Cruse 2012 rows in error had met it (odds ratio, 1.20; 95% CI, 1.00-1.42). Within recordings the index did not track the statistic (pooled slope, 0.04; 95% CI, 0.0001-0.09; within-recording Spearman correlation, 0.03; between-recording correlation, 0.27), so this explanation was not pursued. Heart-rate fluctuation, a marker of arousal that is not computed from the EEG, was associated with the index on the fresh patients (standardized β, 0.21; 95% CI, 0.05-0.37; n = 283; bar met), but the label-aligned heart-rate changes did not predict false calls beyond chance (aligned odds ratio, 1.15, against a circular-shift placebo median of 1.17; empirical P = .56). Alpha-peak prominence (odds ratio per SD, 0.99; 95% CI,

0.86-1.15) and spindle-like activity (odds ratio, 1.06; 95% CI, 0.96-1.14; Holm-adjusted P = .45) were not associated with false calls.

#### Supplementary Results 9. Sensitivity analyses and classification agreement

Under the generous reading, which also allows readings with an unstated call rule, 2 of 9 analyses were inflated (Goldfine 2011 and Guger 2018); the du Bois 2026 rate fell to 36 of 707 (5.1%; 95% CI, 3.6%-7.0%) through a permutation evaluation with cross-validation by whole blocks (Table S7C in Supplementary Data S2). Under the strict reading, in which both classifications must judge every choice stated or not stated, 4 of 9 were inflated, adding Edlow 2017 (242 of 796; 30.4%; 95% CI, 27.2%-33.7%) (Table S7D in Supplementary Data S2). Across the readings eligible for the primary analysis the rate ranged from 23.3% to 65.5% for Goldfine 2011, 22.2% to 84.6% for du Bois 2026 among corrected readings (90.0% for its a priori default, a liberal departure from the publication; Table S7B in Supplementary Data S2), 13.8% to 53.5% for Guger 2018, 0.4% to 30.4% for Edlow 2017, 1.5% to 84.2% for the motor imagery analysis, 3.7% to 9.1% for Gibson 2016, 0.0% to 8.3% for Cruse 2011 and 2.4% to 2.9% for Cruse 2012; the generous reading extended the du Bois 2026 range to 99.9%. The endpoints are selected readings, not estimates, and each is shown with its own interval (Table S7E in Supplementary Data S2). The two blind classifications agreed on whether a choice was consistent with or contradicted the publications for 88.9% of the 117 classified choices (Cohen κ, 0.73; 95% CI, 0.59-0.86; 77.8% and κ, 0.67; 95% CI, 0.56-0.77, across the four classes).

#### Supplementary Results 10. Post-arrest coma

Of 604 comatose patients in I-CARE, 60 had no usable window (37 with no EEG segment starting 12 to 36 hours after return of circulation, 19 with no clean 30-minute window and 4 with no segment long enough) and 544 were analyzed; 54 of them lacked a covariate or a value of the mechanism measure, leaving 490 with complete data (Table 1). Good outcome was as common outside the model as in it (40 of 114, 35.1%, against 185 of 490, 37.8%; Fisher exact P = .67).

Every positive call in these windows is false, because no command or stimulus was given. Under the primary reading (Supplementary Methods 12, thirteenth item), du Bois 2026 called 129 of 464 patients (27.8%; 95% CI, 23.8%-32.1%), Guger 2018 64 of 481 (13.3%; 95% CI, 10.4%-16.7%), Goldfine 2011 54 of 446 (12.1%; 95% CI, 9.2%-15.5%) and Claassen 2019 40 of 489 (8.2%; 95% CI, 5.9%-11.0%); Gibson 2016 called 4.6%, the motor imagery analysis 0.5% and Edlow 2017 0.4% (Table S16A; Table S16B in Supplementary Data S2). Cross-fitted rates differed from the selected rates by 0.5 percentage points or less. No consistent reading of Cruse 2011, Cruse 2012 or Chennu 2013 fit the 30-minute windows; their originally planned shortened-to-fit readings called 1.9%, 4.4% and 0.0%. Run later at full length on longer windows of 231 patients (Supplementary Methods 18), they called 5 of 205 (2.4%; 95% CI, 0.8%-5.6%), 6 of 223 (2.7%; 95% CI, 1.0%-5.8%) and 0 of 198 (0.0%; 95% CI, 0.0%-1.8%) (Table S39A). The covert-processing analysis called 32 of 439 (7.3%; 95% CI, 5.0%-10.1%). Under the originally planned reading, Goldfine 2011 called 79 of 428 (18.5%), Claassen 2019 8.2%, Guger 2018 4.2% and du Bois 2026

1.7%, the last two through readings with a paradigm contradicting the publication or an unstated call criterion, as on clinical EEG. Under the generous reading the du Bois 2026 rate fell to 1.7% through a permutation evaluation; under the strict reading Edlow 2017 called 50 of 484 (10.3%). Each patient contributed one window, so per-patient rules over repeated runs were not evaluated. Pooled over overlays, as on clinical EEG, the randomization-based versions called 3.2% to 6.9%; the highest rate, Goldfine 2011's (61 of 886 overlays, about 2 per patient), is above the 5% point-estimate bar. These rates are estimates; the Holm test computed as on clinical EEG found an excess for one Goldfine 2011 timing (35 of 440 overlays, 8.0%; Holm-adjusted P = .01), so the randomization-based design missed its bar for Goldfine 2011 in coma. False calls did not fall with the background: the du Bois 2026 reading called 26 of 72 patients with a suppressed background (36.1%; 95% CI, 25.1%-48.3%), Guger 2018 11 of 76 (14.5%) and Goldfine 2011 5 of 64 (7.8%) (Table S16G). Read on all 544 patients with a usable window, including the 54 without a complete covariate or mechanism value, the same primary readings called 140 of 513 patients (du Bois 2026; 27.3%; 95% CI, 23.5%-31.4%), 75 of 534 (Guger 2018; 14.0%; 95% CI, 11.2%-17.3%), 57 of 494 (Goldfine 2011; 11.5%; 95% CI, 8.9%-14.7%) and 43 of 543 (Claassen 2019; 7.9%; 95% CI, 5.8%-10.5%); no reading was reselected and no outcome was read. The main text reports these 544-patient rates, their split by background (Table S16H) and the du Bois 2026 pathway on the 544 (Table S18); Tables S16A to S16E (16B in Supplementary Data S2), 16G and 20 use the 490 with complete data, and Table S16F uses its own populations.

The planned outcome test found no association between the mechanism measure and good outcome (odds ratio per SD, 0.79; 95% CI, 0.59-1.03; P = .10; Table S16C), a null result under the planned wording; the interval does not exclude 41% lower odds per SD, and the point estimates were below 1 in nearly every analysis. The estimate was 0.75 (95% CI, 0.54-0.98; P = .04) without the 40 patients of an earlier null set, 0.91 (0.69-1.18) with foundation-model background labels, 0.92 (0.64-1.29) without patients with a flat channel and 0.68 to 1.01 when each hospital was left out in turn. Adding the measure to the background-only model changed the leave-one-hospital-out AUROC by 0.009 (95% CI, -0.002 to 0.020). The clinical EEG mechanism model transported with a calibration intercept of -0.31 (95% CI, -0.38 to -0.24) and a slope of 0.81 (95% CI, 0.77-0.85).

False calls related inconsistently to outcome (Table S16D). Under the a priori default readings, no analysis's false calls went with better outcome, and the du Bois 2026 default reading's false calls went with lower odds of good outcome (odds ratio, 0.53; 95% CI, 0.28-0.96). Under the primary readings, the du Bois 2026 false calls went with higher odds of good outcome (odds ratio, 1.81; 95% CI, 1.04-3.47; uncorrected P = .04, one of 7 estimable analyses). Every other interval included 1. These estimates are not ranked. Because a call that is false by construction could still go with recovery, an association between calls and recovery cannot by itself show that the calls reflect command-following. Pooled over every reading, including liberal departures, false calls were 19.9% in inflated designs, 54.9% in immune designs (dominated by the du Bois 2026 training readings) and 17.9% in deflated designs (Table S16E); these pooled rates are not rates of any published method.

The arousal analysis in coma was secondary, with its code fixed before the post-arrest outcomes were read (Table S16F). In continuous or discontinuous backgrounds, the heart-rate fluctuation was not coupled to slow EEG non-stationarity (standardized beta, -0.04; 95% CI, -0.22 to 0.14; 140 patients), so the coupling seen on clinical EEG was not shown in coma. Non-stationarity was not associated with good outcome (odds ratio per SD, 0.65; 95% CI, 0.37-1.01; P = .06; 245 patients; full sample, 0.77; 95% CI, 0.55-1.05). By the stopping rule fixed for this analysis, no claim is made that the non-stationarity carries meaning in coma. Sedation was not recorded, and outcome after cardiac arrest partly follows decisions to withdraw life-sustaining treatment that were informed by EEG.

#### Supplementary Results 11. Replication

All 730 listed patients had a 30-minute window. Windows with an error, no fitting overlay or no usable trial left the denominator: none for the Claassen 2019 and du Bois 2026 analyses, 8 for Goldfine 2011, 12 for Edlow 2017, 24 for Gibson 2016, 60 for Guger 2018 and 169 for Guger 2017 (Table S17).

The 3 analyses above 5% in development stayed above 5%, each with a Holm-adjusted P below .001 and each inside its 95% prediction interval: Goldfine 2011 called 165 of 722 patients (22.9%; 95% CI, 19.8%-26.1%), du Bois 2026 181 of 730 (24.8%; 95% CI, 21.7%-28.1%) and Guger 2018 84 of 670 (12.5%; 95% CI, 10.1%-15.3%). Under the published two-run rule, Guger 2018 called 152 of 652 (23.3%; 95% CI, 20.1%-26.8%), against 24.5% in development.

The other 4 stayed below 10% (Holm-adjusted $P < .001$ for each): Edlow 2017 called 2 of 718 (0.3%; 95% CI, 0.0%-1.0%), Guger 2017 2 of 561 (0.4%; 95% CI, 0.0%-1.3%), Gibson 2016 37 of 706 (5.2%; 95% CI, 3.7%-7.2%; not above 5%, one-sided $P = .41$) and Claassen 2019 25 of 730 (3.4%; 95% CI, 2.2%-5.0%). Six of the 7 rates lay inside their prediction intervals. The Claassen 2019 rate lay below its interval (4.0%-9.8%) and below its development rate of 33 of 481 (6.9%; Fisher exact $P = .008$). The development rate came from a 481-patient subset, its windows were placed by the analysis's own seeded draw rather than the common planner (Supplementary Methods 15), and background labels were not computed for the replication windows. Rerun with the replication's window placement (14 of the 481 windows had no fitting overlay), the Claassen 2019 development windows were called in 26/467 (5.6%; 95% CI, 3.7%-8.1%) against 33/467 at the original placement (exact McNemar $P = .40$) and 25/730 in the replication (Fisher $P = .08$); these data cannot tell whether window placement explains the difference.

Run later at full length on longer windows of the replication patients whose recordings held one (Supplementary Methods 18), the three analyses left out of the replication also stayed below 5%: Cruse 2011 called 7 of 365 (1.9%; 95% CI, 0.8%-3.9%), Cruse 2012 17 of 495 (3.4%; 95% CI, 2.0%-5.4%) and Chennu 2013 0 of 231 (0.0%; 95% CI, 0.0%-1.6%) (Table S39A). The Cruse 2011 rate is above the 0.5% its 99% binomial interval implies (one-sided exact $P = .003$), though not above 5%.

In the du Bois 2026 decision pathway on the replication set (Table S18), 181 of 730 patients (24.8%) met the advancement criterion, 177 (24.2%) had a significant run,

43 (5.9%; 95% CI, 4.3%-7.9%) were positive under the permutation reading and 13 (1.8%; 95% CI, 1.0%-3.0%) under advancement and the permutation reading together, close to the development shares (22.2%, 21.4%, 5.1% and 2.3%).

#### Supplementary Results 12. Independent implementation, repeated recordings, outcome exposure, published cohorts and patient positive controls

Independent implementation. On the same 200 clinical EEG development windows, the independent du Bois 2026 implementation called 192 (96.0%; 95% CI, 92.3%-98.3%) and this study's 53 (26.5%; 95% CI, 20.5%-33.2%), agreeing on 29.5% of windows (kappa, 0.02); the independent rate lay above the range of readings consistent with the publication (22.2%-90.0%). For Guger 2018, on the 178 windows with a row from both versions, the independent implementation called 31 (17.4%; 95% CI, 12.2%-23.8%) and this study's 15 (8.4%; 95% CI, 4.8%-13.5%), agreeing on 83.1% (kappa, 0.26); the independent rate lay inside the range (13.8%-53.5%). Neither met the agreement bar of kappa 0.6, and both independent versions called more windows than this study's (Table S24). The two versions of each text therefore disagreed: the check shows that the texts do not fix one executable analysis, not which version is closer to the original. For du Bois 2026, Supplementary Results 13 measures where the two part: the independent version follows more of the choices the publication states, the fold rule, which the publication does not state, accounts for most of the difference in rates, and agreement on which windows are called stays low after every change. The independent versions' recorded choices are in Table S24B in Supplementary Data S2.

Repeated recordings. Hypothetical. Treating each further recording as another patient's first window, which does not measure within-patient variation, the Claassen 2019 rule with its correction across a patient's recordings would call 6.9% of clinical EEG patients with 1 recording, 8.6% with 2 and 10.6% with 4, falling to 3.8% and 4.6% with 7 and 8 recordings as the correction becomes stricter; identical recordings would give 6.9% at any number. In post-arrest coma, the rates were 8.2% with 1 recording and 13.6% with 5. Edlow 2017 would call at most 1.8% of clinical EEG patients and 2.1% in coma with 5 sessions (Table S25 in Supplementary Data S2). Uniformity of the Edlow permutation P values on command-free first windows could not be tested: 99.6% were censored at .05 by sequential stopping.

Outcome exposure. Among comatose patients with a poor outcome, du Bois 2026 called 73 of 287 (25.4%; 95% CI, 20.5%-30.9%), Guger 2018 44 of 300 (14.7%; 95% CI, 10.9%-19.2%), Goldfine 2011 40 of 273 (14.7%; 95% CI, 10.7%-19.4%) and Claassen 2019 29 of 304 (9.5%; 95% CI, 6.5%-13.4%). Among patients assessed by all 3 analyses above 5%, 127 of 270 with a poor outcome (47.0%; 95% CI, 41.0%-53.2%) and 79 of 171 with a good outcome (46.2%; 95% CI, 38.6%-54.0%) were called positive by at least 1 (Table S26). This is the share of patients with at least one positive classification under three sham tests, one of them a training-entry criterion; it is not a diagnostic error rate, and a poor outcome does not establish absence of consciousness at the time of recording.

Published cohorts. These comparisons are exploratory. The published cohorts differ from these recordings in patients, equipment, timing and processing, so they are not estimates of diagnostic accuracy, and a comparison that is not significant does not show that published patients responded at the no-command rate. Against the two-session no-command rate of the du Bois 2026 criterion on clinical EEG (30 of 95; 31.6%), the published advancement rates after Phase I were higher in minimally conscious (13 of 17; Holm-adjusted P = .002) and locked-in patients (10 of 11; Holm-adjusted P < .001) and higher but not significantly in unresponsive patients (8 of 14; P = .06; Holm-adjusted P = .06). The Claassen 2019 activation rate (16 of 104; 15.4%) exceeded the two-recording rate of the Claassen 2019 rule in post-arrest coma (9.3%, independent recordings) before adjustment (P = .03) but not after (Holm-adjusted P = .06); across the 95% CI of that rate, P ranged from .001 to .34, and against the 1-recording rate (8.2%) it was .01. Against the secondary no-command rates (du Bois 2026, 18 of 38 prolonged-DoC patients over 2 sessions, 47.4%; Claassen 2019, two-recording rate on clinical EEG, 8.6%), P values were .38 (unresponsive), .04 (minimally conscious), .01 (locked-in) and .02 (Claassen 2019) (Table S27).

Stroke positive control. In 47 patients with acute stroke, Guger 2017 called 9 (19.1%; 95% CI, 9.1%-33.3%) with true labels on sensorimotor channels and 3.2% (95% CI, 2.1%-4.4%) of label-shuffled sessions; 21 patients (44.7%) had at least 1 false call in 20 shuffles, and calls with true labels exceeded false calls by 16.0 percentage points (95% CI, 5.1-27.3). On all channels, the rates were 7 of 47 (14.9%) and 2.4% (Table S28).

P300 positive control in amyotrophic lateral sclerosis. Nine of 48 patients passed the check (34 required), with a grand-average target-minus-non-target difference of +0.14 μV. The target response peaked at 150 to 300 ms (+1.6 μV at Pz) and was followed by a negative deflection from 450 to 700 ms, which cancelled within the fixed window. The window was not changed, and the control was not used.

#### Supplementary Results 13. Genuine tasks, sham labels, the du Bois 2026 implementations and calibration within each design

Tactile P300. With the attended finger as the label, the primary reading called 36 of 38 sessions (94.7%; 95% CI, 82.3%-99.4%) and all 19 participants; with an unattended finger as the label, it called 10 of 76 runs (13.2%), near the threshold's chance rate of about 12%, and under the 2-run rule 9 of 38 sessions (23.7%; 95% CI, 11.4%-40.2%; above 5%, $P < .001$) and 8 of 19 participants (exact McNemar, 27 vs 0 discordant sessions, $P < .001$). Real touch to an unattended finger therefore did not raise calls above the threshold's chance rate. The default reading called 35 and 13 of 38 sessions and the independent implementation 37 and 13. The randomization test called 33 of 38 with true labels (86.8%; 95% CI, 71.9%-95.6%) and 2 of 38 with sham labels (5.3%; 95% CI, 0.6%-17.7%; above 5%, $P = .57$). Sham calls were not concentrated where the sham label stayed on the attended hand (1 of 7 sessions, against 8 of 31 where it moved to the other hand). The positive control did not pass its preset bar, because the upper 95% limit of 36 of 38 (99.4%) falls short of the published 3 of 3; detection was nevertheless near complete and well above sham calls (Table S32).

Stroke motor imagery. Of 64 sessions, 3 byte-identical ones were excluded, leaving 61 sessions of 23 patients. The Goldfine 2011 primary reading called 98.4% of sessions with true labels (95% CI, 95.0%-100%), 2.5% with rest-only sham labels (95% CI, 1.2%-4.4%) and 12.0% with half of each run's labels exchanged (95% CI, 7.9%-16.8%). With the command-level jackknife it called 93.4% (95% CI, 82.5%-100%), 95% of the primary reading's detection, with 0.0% sham calls and 0.6% exchanged-label calls (95% CI, 0.1%-1.2%); every Goldfine 2011 reading separated true from null labels and was calibrated against the rest-only sham. No true-label call rested on posterior sites only. Our du Bois 2026 implementation with random folds met the advancement criterion in 86.9% of sessions with true labels (95% CI, 73.8%-96.8%) and 46.7% with labels redrawn at random (95% CI, 43.2%-50.2%); the second implementation, in 93.4% (95% CI, 86.7%-98.4%) and 68.0% (95% CI, 65.4%-70.4%). On real patient EEG with uninformative labels, the advancement criterion is thus met in about half to two thirds of sessions, in a design where random folds are legitimate (Table S33).

The second du Bois 2026 implementation. On the 200 development windows, the second implementation as written called 192 (96.0%; 95% CI, 92.3%-98.3%), reproducing its rate in Table S24, and our primary reading 53 (26.5%). Applied alone, block-wise folds lowered the second implementation's rate to 85 (42.5%; 95% CI, 35.6%-49.7%; exact McNemar, 109 vs 2 discordant windows), closing 77% of the rate gap; our peak rule alone, to 164 (82.0%), closing 20%; a 100-ms step, to 184 (92.0%), 6%; and each other departure, to 188 to 191 (94.0%-95.5%), at most 3%. With every listed change applied cumulatively, the rate was 81 (40.5%); the training scheme and the spatial-filter covariance, which were not varied, still differ from our primary reading, and agreement on which windows are called stayed at kappa 0.10 or less for every variant, meeting the reading fixed beforehand that the remaining choices matter. Our implementation with random folds called 174 (87.0%). In the factorial, the 84-configuration search raised the rate under either fold rule and the refined peak only with random folds; under block-wise folds our peak rule called more windows than the refinement. The most conservative combination tested (block-wise folds, the published peak refinement, a configuration fixed in advance) still called 28 (14.0%; 95% CI, 9.5%-19.6%). Run literally, the 100-permutation evaluation passed all 200 windows with random folds (95% CI, 98.2%-100%) and 112 (56.0%) with block-wise folds; with the advancement criterion added, 192 (96.0%) and 61 (30.5%). With the search repeated inside each permutation, it passed 156 (78.0%; 95% CI, 71.6%-83.5%) with random folds and 12 (6.0%; 95% CI, 3.1%-10.2%; above 5%, P = .30) with block-wise folds, the same with the advancement criterion added; every window passed with block-wise folds also passed with random folds. On the first 60 windows, fixed before the run, the rates were 43 of 60 (71.7%) and 1 of 60 (1.7%). With block-wise folds the P values were far from uniform: the observed accuracy usually fell below the permuted ones (median P, 1.0; 111 of 200 at P = 1.0), as when held-out blocks differ from training blocks, so the 6.0% is not evidence of calibration, and detection by this version was not measured. On synthetic recordings without drift, the repeated-search evaluation passed 2 of 30 (random folds) and 0 of 30 (block-wise); with strong drift, 29 of 30 and 8 of 30. Repeating the search therefore removes the selection effect; on synthetic EEG, random folds failed only with drift, consistent with a free permutation of single trials breaking the alignment between the block design and slow drift that the true labels keep (Table S30).

The Goldfine 2011 variance. The primary reading reproduced the stored call in all 342 development windows and called 79 (23.1%; 95% CI, 18.7%-27.9%); decimating the frequency grid alone, 59 (17.3%); and the command-level jackknife, with a normal reference, a t reference or a t reference on the decimated grid, 2 (0.6%; 95% CI, 0.1%-2.1%), calibrated by the 10% bar. The command-level variance of the standardized difference was a median 3.92 times its value for independent estimates (IQR, 2.88-6.18), at least 2 in 94% of windows, and larger in windows the primary reading called (median 5.19) than in the others (3.63), meeting the bar for dependence as the mechanism. On synthetic recordings, the primary reading called 0, 2 and 29 of 100 as slow amplitude drift rose (variance inflation 1.05, 1.54 and 3.68) and the command-level jackknife none at any drift (Table S31).

Calibration within each design. Achieved held-out false calls lay at or slightly above the per-assessment target for Goldfine 2011 and du Bois 2026 (Table S34, in parentheses; targets 5.0%, 2.5%, 1.0% and 0.5% for k = 1, 2, 5 and 10; 10 of 20 cells above it, the largest relative excesses 1.5% at k = 5 for the command-level Goldfine 2011 stroke sessions and 0.7% at k = 10 for the du Bois 2026 stroke sessions). For Guger 2018 they were further above it: 6.4%, 1.8% and 1.5% for the auditory counting null at k = 1, 5 and 10, and 10.5% for the tactile runs at k = 1 (95% CI, 2.6%-18.4%), and from k = 2 the tactile threshold reached the largest of the 38 sham runs per half, so its held-out rate (2.6%) could not fall to the smaller targets. Participant-level intervals were close to the exact intervals for the clinical EEG and stroke designs and wider for the tactile runs from k = 2 (upper bounds 15.8% to 17.1%, against 9.2%). Detection in these cells is optimistic, and the thresholds should be read as targeting, not achieving, the stated patient-level rate. With 1 assessment, single-assessment detection was 100% (healthy volunteers) and 96.7% (stroke) for the Goldfine 2011 statistic, 21.6% and 39.3% for the du Bois 2026 paired t, and 61.5% (auditory counting) and 61.8% (tactile runs) for the Guger 2018 statistic. With thresholds targeting a 5% chance of any false call over 5 assessments, these were 100% and 95.1%, 2.0% and 16.4%, and 7.7% and 30.3%; with 10, 96.1% and 95.1%, 2.0% and 9.8%, and 0% and 30.3%. A genuine responder assessed repeatedly has more than one chance of a call, so single-assessment detection understates detection over the k assessments. The published rules detected more only by calling more: the du Bois 2026 rule detected 96.1% of the volunteers' original-assembly sessions (41 of 51 one-source sessions, Supplementary Results 6) at its 22.2% false-call rate, and the Guger 2018 rule 69.2% at 13.8%. The randomization-based versions, on randomized-order sessions without slow drift, detected 96.1%, 86.3% and 30.8% of volunteers at 1 assessment and 94.1%, 82.4% and 23.1% at 5; their null rates at 5 assessments were 1.6%, 0.6% and 0.5% per assessment. The command-level Goldfine 2011 statistic detected 98.4% of stroke sessions at k = 1 and 95.1% at k = 5 and 10.

Frozen test of the command-level Goldfine 2011 variance. Every usable window reproduced its stored published call (722 of 722 replication windows, 494 of 494 post-arrest windows; 8 and 49 windows without usable trials and 1 without a fitting overlay left out, as in the primary analysis). The published reading called 165 of 722 replication patients (22.9%) and 57 of 494 post-arrest patients (11.5%); the command-level reading called 6 (0.8%; 95% CI, 0.3%-1.8%) and 12 (2.4%; 95% CI, 1.3%-4.2%), below 5% by the frozen test (Holm-adjusted $P < .001$ and $P = .003$). No window was called by the command-level reading alone. With the t reference, 4 and 9; with the t reference on the decimated grid, 3 and 8. By continuity grade after cardiac arrest, the command-level reading called 6 of 174 patients with burst suppression (3.4%), 5 of 110 with a discontinuous (4.5%), 1 of 135 with a continuous (0.7%) and none of 75 with a suppressed background; in the development windows, none of 112 awake, 150 sleep and 17 ICU-typical windows and 2 of 63 mixed (Table S35).

Time-matched specificity. Contrasting left- with right-hand imagery without rest, the published reading called 5 of 47 patients (10.6%; 95% CI, 3.5%-23.1%), no more than expected from its 7.7% rate with pair-swapped labels (95% CI, 4.3%-11.5%; $P = .29$ for 5 or more), and the command-level reading 2 (4.3%; 95% CI, 0.5%-14.5%) against 0.5% (95% CI, 0.0%-1.2%; $P = .03$). All 5 published calls and 1 of the 2 command-level calls had contralateral sensorimotor bins and none had posterior bins only, which meets the fixed rule for both readings; but 34 of the 72 null calls of the published reading were also contralateral and none was posterior only, so the rule does not separate true from null calls here. With rest excluded, either variance detects few acute-stroke patients; whether the imagery-against-rest detection in the

Wang recordings reflects the content of the imagery was not tested within that dataset (Table S37).

Cluster-aware inference. With participants as clusters, the tactile primary reading called 94.7% of sessions with the attended finger (95% CI, 86.8%-100%) and 23.7% with an unattended finger (95% CI, 10.5%-36.8%), a difference of 71.1 points (95% CI, 55.3-84.2); 17 participants had more true than sham sessions called, none the reverse and 2 tied ($P < .001$). The change in stroke detection from the published to the command-level Goldfine 2011 jackknife was 4.9 points (95% CI, 0.0-12.3), from 3 sessions called by the published reading only and none by the command-level reading only (exact McNemar $P = .25$); the lower limit of 0 reflects the absence of discordance in the other direction (Table S36).

Direct patient-level calibration. In the clinical null of Guger 2018, the independence-based thresholds gave measured held-out patient-level false-call rates of 4.8% (95% CI, 3.5%-6.6%; $k = 2$, 806 patients), 6.2% (4.5%-8.3%; $k = 5$, 661) and 6.4% (3.8%-10.0%; $k = 10$, 266), slightly above 5% at $k = 5$ and 10; the two halves' thresholds differed widely at these levels, so part of the excess is sampling error in a threshold at a small level. Thresholds set on each patient's maximum over k runs gave 5.1% (3.7%-6.8%), 4.8% (3.3%-6.8%) and 6.4% (3.8%-10.0%), with volunteer detection of 38.5%, 7.7% and 7.7%, close to or above the independence-based detection (30.8%, 7.7% and 0%). For du Bois 2026 ($k = 2$, 95 patients), 7.4% (3.0%-14.6%) with the maximum-based threshold and 3.2% (0.7%-9.0%) with the independence-based one. The low Guger 2018 detection at a 5% patient-level rate therefore does not depend on the independence assumption (Table S38).

#### Supplementary Results 14. Full-length schedules and Claassen 2019 on six sites

On the 89 clinical development recordings that hold 20 blocks in a clean stretch, Chennu 2013's two eligible 20-block readings called 0 and 2 patients, and the reading with fewer calls was chosen (0 of 89; 0.0%; 95% CI, 0.0%-4.1%; cross-fitted held-out rate 2 of 89, 2.2%; 95% CI, 0.3%-7.9%); it called none of 60 white-noise recordings. In the replication set it called 0 of 231 (0.0%; 95% CI, 0.0%-1.6%), Cruse 2011 7 of 365 (1.9%; 95% CI, 0.8%-3.9%) and Cruse 2012 17 of 495 (3.4%; 95% CI, 2.0%-5.4%); 33 and 45 of their windows had no usable trials. After cardiac arrest, in the 231 patients with a long clean window, Cruse 2011 called 5 of 205 (2.4%; 95% CI, 0.8%-5.6%), Cruse 2012 6 of 223 (2.7%; 95% CI, 1.0%-5.8%) and Chennu 2013 0 of 198 (0.0%; 95% CI, 0.0%-1.8%) (Table S39A). None of these rates is above 5%. The Cruse 2011 rates in new windows were above the 0.5% its 99% binomial interval implies (one-sided exact P = .003 in the replication set and .004 after arrest), although its development rate was not (1 of 147, 0.7%). The covert-processing analysis called 58 of 693 replication patients (8.4%; 95% CI, 6.4%-10.7%), above 5% (one-sided P < .001), as in development (41 of 565, 7.3%).

Claassen 2019 on the 6 prolonged-DoC sites called 1 of 38 patients by its published test (2.6%; 95% CI, 0.1%-13.8%) and none by its randomization-based test (0.0%; 95% CI, 0.0%-9.3%) (Table S39B). On the 481 clinical EEG recordings of its development analysis, the 6-site pipeline called 40 by the published test (8.3%; 95% CI, 6.0%-11.2%) against 33 with 19 sites (6.9%; 95% CI, 4.8%-9.5%; exact McNemar P = .42), and 26 against 21 by the randomization-based test (P = .51). The calls fell largely on different recordings: by the published test, 9 were called with both montages, 24 with 19 sites only and 31 with 6 sites only. The reduced montage alone therefore did not lower the rate of false calls; it changed which recordings were called.

#### Supplementary Tables

Table S1. Datasets, licences and splits. Access terms are as stated by the data host. VS/UWS indicates vegetative state or unresponsive wakefulness syndrome; MCS, minimally conscious state; DoC, disorders of consciousness.

| Dataset | Content and use | Access and licence | Analysis sample | Excluded from every reported number |
| --- | --- | --- | --- | --- |
| SpikeNet 2.0 (Brain Data Science Platform) | De-identified routine, epilepsy-monitoring and long-term clinical EEG from three institutions; 19-channel 10-20 montage, 128 Hz as staged. Primary no-command and no-stimulus EEG | Restricted access under the platform data use agreement; collected under a protocol approved by the Mass General Brigham Institutional Review Board with a waiver of consent | 974 recordings, one per patient (Table S6) | 20 development recordings; 6 recordings touched before the freeze (excluded everywhere) |

| Dataset | Content and use | Access and licence | Analysis sample | Excluded from every reported number |
|---|---|---|---|---|
| EEG foundation model background labels (Brain Data Science Platform) | Model-assigned background for each analysis window: normal awake, sleep, transitioning, mixed, diffuse slowing, burst suppression, periodic patterns. A model-assigned proxy, not a clinical label | Used under the platform terms | All 974 recordings | Not applicable |
| Overnight polysomnography in chronic disorders of consciousness (Mendeley Data, parts 1 and 2) | Six EEG channels (F3, F4, C3, C4, O1, O2); diagnosis (VS/UWS or MCS) by the Coma Recovery Scale-Revised. Prolonged DoC null set; estimation only | CC BY 4.0 | 38 patients (28 VS/UWS, 10 MCS); clinical table linked for 30 | 1 byte-identical duplicate removed; 3 development patients |
| I-CARE (PhysioNet, version 2.1) | Post-arrest coma EEG from five hospitals, one 30-minute stretch nearest 24 hours after return of circulation; outcome (Cerebral Performance Category at 3-6 months). Post-arrest null set and outcome model; estimation only | CC BY-NC-SA 4.0 | 544 patients with a usable window; 490 with complete data in the false-call rates and the outcome model | 3 development patients; 60 without a usable window (37 with no segment starting 12-36 hours after return of circulation, 19 with no clean 30-minute stretch, 4 with no segment long enough); 54 without complete data (37 out-of-hospital status, 6 rhythm, 1 age, 10 no mechanism measure) |
| OpenBMI (GigaDB) | Healthy-participant motor imagery. Detection of true responders and label-shuffled controls | CC0 1.0 | 51 participants (Claassen 2019: seeded sample of 20) | 3 development participants |

| Dataset | Content and use | Access and licence | Analysis sample | Excluded from every reported number |
|---|---|---|---|---|
| EEG Motor Movement/Imagery Database (PhysioNet) | Healthy-participant motor imagery and execution runs. Secondary detection arm; source trials are reused | Open Data Commons Attribution License v1.0 | Up to 102 participants (20 for the slowest analyses) | 3 development participants |
| Four-class auditory BCI, Kojima (Harvard Dataverse) | Healthy-participant attended auditory counting (detection) and the unattended stream of the same recordings (sensory responses) | CC0 1.0 | 13 participants | 2 development participants |
| ERP CORE mismatch negativity | Healthy-participant passive oddball. Sensory responses read as volition | As stated by the host: CC BY 4.0 on the project record; CC BY-SA 4.0 in the licence file; CC0 in the dataset description. Used for analysis with attribution | 38 participants | 2 development participants |
| Auditory oddball, OpenNeuro ds003061 | Healthy-participant passive oddball. Sensory responses read as volition | CC0 | 11 participants | 2 development participants |
| Acute-stroke hand motor imagery, Liu 2024 (figshare) [38] | Patients 1 to 30 days after stroke, 40 strictly alternating left and right hand imagery trials. Instructed positive control (Table S28) and time-matched specificity of Goldfine 2011 (Table S37) | CC BY 4.0 | 47 patients | 3 recordings duplicating other patients (sub-05, 15 and 49) |

| Dataset | Content and use | Access and licence | Analysis sample | Excluded from every reported number |
|---|---|---|---|---|
| Braille tactile P300 brain-computer interface, Grigoryan 2020 (G-Node) [36] | Blind and sighted adults counting touches on 1 of 8 fingertips, 2 sessions each. Tactile P300 positive control with sham labels (Table S32) | CC0 1.0 | 19 participants, 38 sessions | 1 participant used only to build the loader |
| Sixth-finger and affected-hand motor imagery after stroke, Wang 2026 (figshare) [37] | Patients 3 to 14 months after stroke at 3 time points, 4 runs each; imagery against rest (Goldfine 2011) and between imagery types (du Bois 2026) with sham labels (Table S33) | CC BY 4.0 | 23 patients, 61 sessions | 1 patient used only to build the loader; 3 byte-identical sessions |

Table S2. Published analyses and their reimplementation. Analysis windows are the schedule lengths each analysis needs. k is the number of assessments per patient used for the per-patient rule. Healthy-participant detection is the published rate used as the bar for the positive control (Supplementary Methods 9).

| **Analysis [publication]** | **Paradigm and analysis window** | **Statistic** | **Stated call rule** | **Published assessments per patient (k)** | **Published healthy-participant detection** | **Reimplementation** |
|---|---|---|---|---|---|---|
| Claassen 2019 [1] | Spoken commands to keep opening and closing, and to stop opening and closing, each hand, in alternating blocks (48 commands over 1,505 s) | Linear support vector machine on spectral band-power features of 19 channels in a current-source-density montage; cross-validated AUC with a one-tailed permutation test (500 permutations) | AUC above 0.5 and permutation P < .05, Benjamini-Hochberg control across a patient's recordings | 2 (1-8 recordings per patient; mean 2.3) | 10/10 | Authors' public code reproduced |

| Analysis [publication] | Paradigm and analysis window | Statistic | Stated call rule | Published assessments per patient (k) | Published healthy-participant detection | Reimplementation |
|---|---|---|---|---|---|---|
| Goldfine 2011 [9,25] | Runs of 8 motor-imagery commands with a 9-s response period cut into 3-s epochs, 1,065 s per assessment | Multitaper spectral two-group test (4-24 Hz) comparing task and rest epochs per channel and frequency; jackknife variance; false discovery rate control | Positive when a contiguous difference over more than 2 Hz in one run is supported in another run and at least one difference survives false discovery rate control across channels and frequencies; positive on at least 1 paradigm in at least 1 assessment | 1 (28 patients, 34 assessments) | 15/15 (4/4 in the first report) | Core statistic from the public toolbox; manual epoch pruning automated |
| Cruse 2011 [10] | Tone-cued hand and toe movement imagery, 4 to 8 blocks per movement type, 3,244 s per assessment | Band-power classification in four bands (7-30 Hz), cross-validation across blocks; binomial confidence interval of the accuracy against chance and against a baseline period | 99% and 99.9% interval excludes chance and the action-period accuracy exceeds the baseline interval; no correction across patients | 1 | 9/12 | Reimplemented from the published description |

| Analysis [publication] | Paradigm and analysis window | Statistic | Stated call rule | Published assessments per patient (k) | Published healthy-participant detection | Reimplementation |
|---|---|---|---|---|---|---|
| Cruse 2012 [26] | Five blocks of 180 trials of hand-squeeze commands, 1,871 s per assessment | Naive Bayes single-trial classification of band power over time; 10-fold cross-validation; maximum-over-time label permutation (1,000) | Either comparison (left hand or right hand against rest) significant at P < .025, one-tailed | 1 | 5/6 | Reimplemented from the published description |
| Edlow 2017 [11] | Motor imagery: 24-s blocks of hand squeezing and rest, 12 blocks of each, 586 s | Per-second band-power features, support vector machine, 20-fold cross-validation repeated 10 times; label permutation (500) | Permutation P < .05, one-sided | 1 | 12/16 | Reimplemented from the published description |
| Guger 2017 [27-29] | Spoken left and right hand imagery commands, 60 or 120 trials, 1,150 s | Common spatial patterns with linear discriminant classification, 10 x 10-fold cross-validation; accuracy course in 0.5-s steps | Accuracy above a binomial chance bound (63.19% for 60 trials); repeat the run if below | 1 (2 as a sensitivity analysis) | 3/3 | Reimplemented from published descriptions |

| Analysis [publication] | Paradigm and analysis window | Statistic | Stated call rule | Published assessments per patient (k) | Published healthy-participant detection | Reimplementation |
|---|---|---|---|---|---|---|
| du Bois 2026 [30] | Phase I: runs of 6 blocks of 15 trials alternating two imagined movements, 1,065 s | Filter-bank common spatial patterns with mutual-information selection; configuration search; peak decoding accuracy in the task interval against the baseline peak; 6-fold cross-validation | Peak accuracy above 70% or task accuracy above baseline by a one-tailed paired t test across folds ($P < .05$); a pass advances the participant to training | 2 (sessions 1 and 2) | 2/2 | Reimplemented from the published description |
| Guger 2018 [31,32] | Vibrotactile stimulation of both wrists and distractor sites in 60 groups of 8 stimuli, instructed target wrist, 157 s per run | Linear discriminant classification of evoked responses, 8-way accuracy over random split-halves; accuracy curve summarized | Accuracy above 23% in at least 1 run | 2 runs (up to 7 reported per patient) | 3/3 | Reimplemented from published descriptions |
| Chennu 2013 [34] | 20 blocks of spoken words with an explicit target to count, 2,530 s | Global field power cluster permutation against baseline, temporal cluster test, explicit-versus-implicit contrast | Not stated as a rule; the reported positive patient met all three criteria | 1 | 8/8 | Authors' public code, core statistic; manual rejection automated |

| Analysis [publication] | Paradigm and analysis window | Statistic | Stated call rule | Published assessments per patient (k) | Published healthy-participant detection | Reimplementation |
|---|---|---|---|---|---|---|
| Gibson 2016 [33] | Vibrotactile stimulation with an instructed target wrist, 1,130 s | Cluster-mass permutation for the P3a and P3b contrasts; steady-state F test at 5 and 10 Hz | P3b cluster test (target against non-target wrist), tails and cluster alpha not stated | 1 | 10/15 | Reimplemented from the published description |
| Sokoliuk 2021 [35] | Passive listening to isochronous speech (words, phrases, sentences), 1,346 s | Inter-trial phase coherence at the word, phrase and sentence rates | No per-recording call rule; a prognostic cut-off of 0.116 on phrase and sentence coherence | 1 | Not stated | Core statistic from the authors' public code |

Table S4. Departures from the publications shared by every reading of an analysis. The expected direction is the most likely effect on the false-call rate and was not measured here; for du Bois 2026, Table S30 measures each departure, whose effect differed with the fold rule.

| Analysis | Element | Publication | This study (applied to every reading) | Expected direction for false calls |
|---|---|---|---|---|
| Goldfine 2011 | Channels | 37 electrodes in an augmented 10-20 system | 19 standard sites with a Hjorth Laplacian | Probably lower false-call rate for the contiguity criterion; unknown for the any-bin criterion |
| Goldfine 2011 | Contiguity span | At least 2 Hz (Curley 2018) or more than 2 Hz (Goldfine 2011) | More than 2 Hz | Probably lower |
| Goldfine 2011 | Filtering order | 1-50 Hz band-pass after concatenating cleaned epochs | Zero-phase filter on the continuous signal before the Laplacian | Unknown |

| Analysis | Element | Publication | This study (applied to every reading) | Expected direction for false calls |
|---|---|---|---|---|
| Goldfine 2011 | Artifact handling | Independent component analysis and line-noise regression (Goldfine 2011); visual pruning (Curley 2018) | Neither; epochs pruned by the reading's automated rule | Unknown |
| Guger 2017 | Channels | 16 sensorimotor sites | 11 nearest 10-20 sites | Probably lower for common spatial patterns fitted once on all trials |
| du Bois 2026 | Channel sets | 16-channel full set and 9-channel motor set | 17 standard sites (all but Fp1 and Fp2) and a 9-channel motor set | Unknown |
| du Bois 2026 | Sampling rate and acquisition filter | 250 Hz down-sampled to 125 Hz; 0.5-100 Hz eighth-order filter | Native rate; recordings used as stored | Negligible expected |
| du Bois 2026 | Band edges and filters | Six bands from 0.5 to 40 Hz; finite-impulse-response filters | Six bands from 1 to 40 Hz; zero-phase Butterworth filters | Unknown |
| du Bois 2026 | Spatial filters and feature selection | Three filter pairs per band; mutual information with a fixed quantization | Two filters from each end per band; Gaussian mutual information | Probably lower (fewer candidate features) |
| du Bois 2026 | Time step and peak refinement | 40-ms step; raw-curve peak within 150 ms of the smoothed peak | 100-ms step; smoothed maximum | Probably lower for the 70% criterion |
| Cruse 2011 | Channels and reference | 25 electrodes over the motor area, each referenced to its four nearest neighbours | 9 standard sites, each referenced to its four nearest standard sites | Unknown |
| Cruse 2011 | Interpolation and baseline correction | Bad channels interpolated; baseline corrected before the tone | No interpolation; baseline correction omitted (no effect on band power expected) | Unknown / none |

| Analy sis | Element | Publication | This study (applied to every reading) | Expected direction for false calls |
|---|---|---|---|---|
| Edlow 2017 | Seconds kept and artifact rejection | 388 of 576 s; independent component analysis and consensus review | 384 s; no component analysis; columns above 200 μV dropped | Negligible / unknown |
| Cruse 2012 | Montage | FC3-CP3 and FC4-CP4 bipolar | F3-P3 and F4-P4 bipolar | Unknown |
| Guger 2018 | Groups per run | 30 groups of 8 (internally inconsistent in the publication) | 60 groups of 8, 480 stimuli | Unknown |
| Guger 2018 | Channels and reference | 8 channels; right earlobe reference | 7 standard sites; recording's own reference | Unknown |
| Guger 2018 | Trial balancing after rejection | Remove trials until the target ratio of 1 to 7 is restored | Drop a whole group of 8 when any epoch is rejected | Unknown |
| Chenn u 2013 | Channels and reference | 128-channel net, 91 retained, average reference | 19 standard sites, average reference | Probably lower for the baseline-contrast rules |
| Chenn u 2013 | Artifact handling and sampling | Independent component analysis; interpolation; 250 Hz | None; native rate | Unknown |
| Gibson 2016 | Channels | 12 sites | 13 nearest standard sites | Unknown |
| Gibson 2016 | Reference and trial rejection | Right earlobe reference; kurtosis per stimulus type | Recording's own reference; kurtosis z-scored over all kept trials | Unknown; small on a null |
| Sokoli uk 2021 | Preprocessing | Independent component analysis; average reference over all channels; interpolation | None; average over 19 scalp channels; none | Unknown; expected small |

Table S5. Sensory responses read as volition. Rates are per run, with participant-level bootstrap intervals; above 5% is after Holm adjustment across the analyses of the family. Each analysis used its planned lowest-rate reading. The construction check draws sham instructions independently of the EEG, so it is exact by construction and is not a test of false-call control. Where no call occurred, the bootstrap interval is degenerate and the exact interval for 0 of the runs or patients analyzed is shown.

| Dataset | Analysis | Reading used (planned lowest-rate reading) | Runs called, No./No. analyzed | Participants, No. | Runs called, % (95% patient-level bootstrap CI) | Above 5% after Holm adjustment | Construction check: sham instructions, participants called, % |
|---|---|---|---|---|---|---|---|
| ERP CORE mismatch negativity | Chennu 2013 | P3b cluster against baseline, temporal cluster test and explicit-versus-implicit contrast; minimum cluster 10 samples; no epoch rejection; 20 blocks | 1/38 | 38 | 2.6 (0.0-7.9) | No | 1/38 (2.6) |
| ERP CORE mismatch negativity | Gibson 2016 | P3a contrast (wrist versus random subset of back-position stimuli); one-tailed positive, cluster alpha .05; neighbours: Delaunay triangulation | 9/38 | 38 | 23.7 (10.5-36.8) | Yes | 3/38 (7.9) |

| Dataset | Analysis | Reading used (planned lowest-rate reading) | Runs called, No./No. analyzed | Participants, No. | Runs called, % (95% patient-level bootstrap CI) | Above 5% after Holm adjustment | Construction check: sham instructions, participants called, % |
|---|---|---|---|---|---|---|---|
| ERP CORE mismatch negativity | Guger 2018 | 2-stimulus paradigm; Annen 2018 stimulus timing; 7-sample features; median accuracy summary; binomial bar for 10 trials | 13/38 | 38 | 34.2 (18.4-50.0) | Yes | 1/38 (2.6) |
| Auditory oddball (ds003061) | Chennu 2013 | P3b cluster against baseline, temporal cluster test and explicit-versus-implicit contrast; minimum cluster 10 samples; no epoch rejection; 20 blocks | 3/32 | 11 | 9.4 (0.0-18.8) | No | 0/11 (0.0) |
| Auditory oddball (ds003061) | Gibson 2016 | Steady-state F test at Pz on 5 Hz power alone; noise bins excluding harmonics | 0/10 | 5 | 0.0 (0.0-30.8) | No | 0/11 (0.0) |

| Dataset | Analysis | Reading used (planned lowest-rate reading) | Runs called, No./No. analyzed | Participants, No. | Runs called, % (95% patient-level bootstrap CI) | Above 5% after Holm adjustment | Construction check: sham instructions, participants called, % |
|---|---|---|---|---|---|---|---|
| Auditory oddball (ds003061) | Guger 2018 | 2-stimulus paradigm; Murovec 2020 stimulus timing; 8-segment features; median accuracy summary; binomial bar for 10 trials | 5/30 | 11 | 16.7 (3.2-33.3) | No | 0/11 (0.0) |
| Kojima unattended stream | Chennu 2013 | P3b cluster against baseline, temporal cluster test and explicit-versus-implicit contrast; minimum cluster 10 samples; no epoch rejection; 20 blocks | 1/78 | 13 | 1.3 (0.0-3.8) | No | 0/13 (0.0) |
| Kojima unattended stream | Gibson 2016 | Steady-state F test at Pz on 5 Hz power alone; noise bins including harmonics | 2/78 | 13 | 2.6 (0.0-6.4) | No | 0/13 (0.0) |

| Dataset | Analysis | Reading used (planned lowest-rate reading) | Runs called, No./No. analyzed | Participants, No. | Runs called, % (95% patient-level bootstrap CI) | Above 5% after Holm adjustment | Construction check: sham instructions, participants called, % |
|---|---|---|---|---|---|---|---|
| Kojima unattended stream | Guger 2018 | 3-stimulus paradigm; Murovec 2020 stimulus timing; 8-segment features; median accuracy summary; binomial bar for 10 trials | 14/78 | 13 | 17.9 (10.3-25.6) | Yes | 2/13 (15.4) |

Table S6A. Assembly of the clinical EEG analysis set.

| Step | Recordings, No. |
|---|---|
| Design sample listed | 500 |
| Development recordings removed (frozen before any analysis) | -20 |
| Recordings touched before the freeze, removed from every analysis | -6 |
| Design recordings in the analysis set | 474 |
| Added recordings (patient-disjoint from the design sample; never pulled before the freeze) | +500 |
| Clinical EEG analysis set (one recording per patient) | 974 |
| Touched recordings that reached any merged result | 0 |
| Claassen 2019 analysis subset (230 design, 251 added) | 481 |
| Patients with a first-window row in the mechanism analyses | 900 |

Table S6B. First-window EEG background and recordings analyzed, by analysis, under the originally planned reading. The model-assigned background of each analysis's first window; percentages are of the recordings analyzed by that analysis. Denominators under the primary reading, which differ for Cruse 2011 (147), Cruse

2012 (210), Guger 2018 (850) and Sokoliuk 2021 (565), are in Table S7A.

Denominators differ because each schedule needs a clean window of its own length.

| Analysis | Recordings analyzed, No. | Normal awake, No. (%) | Sleep, No. (%) | Transitioning, No. (%) | Mixed, No. (%) | Periodic patterns; diffuse slowing; burst suppression, No. (%) |
|---|---|---|---|---|---|---|
| Claassen 2019 | 481 | 136 (28.3%) | 211 (43.9%) | 84 (17.5%) | 36 (7.5%) | 9 (1.9%); 2 (0.4%); 3 (0.6%) |
| Goldfine 2011 | 703 | 192 (27.3%) | 315 (44.8%) | 115 (16.4%) | 56 (8.0%) | 15 (2.1%); 4 (0.6%); 6 (0.9%) |
| Cruse 2011 | 26 | 8 (30.8%) | 12 (46.2%) | 5 (19.2%) | 1 (3.8%) | 0 (0.0%); 0 (0.0%); 0 (0.0%) |
| Cruse 2012 | 439 | 138 (31.4%) | 184 (41.9%) | 77 (17.5%) | 28 (6.4%) | 7 (1.6%); 2 (0.5%); 3 (0.7%) |
| Edlow 2017 | 796 | 236 (29.6%) | 342 (43.0%) | 121 (15.2%) | 66 (8.3%) | 21 (2.6%); 3 (0.4%); 7 (0.9%) |
| Guger 2017 | 550 | 165 (30.0%) | 231 (42.0%) | 96 (17.5%) | 46 (8.4%) | 5 (0.9%); 1 (0.2%); 6 (1.1%) |
| du Bois 2026 | 707 | 191 (27.0%) | 329 (46.5%) | 107 (15.1%) | 56 (7.9%) | 15 (2.1%); 3 (0.4%); 6 (0.8%) |
| Guger 2018 | 838 | 283 (33.8%) | 372 (44.4%) | 107 (12.8%) | 48 (5.7%) | 16 (1.9%); 4 (0.5%); 6 (0.7%) |
| Chennu 2013 | 467 | 132 (28.3%) | 203 (43.5%) | 91 (19.5%) | 28 (6.0%) | 8 (1.7%); 2 (0.4%); 3 (0.6%) |
| Gibson 2016 | 683 | 191 (28.0%) | 306 (44.8%) | 108 (15.8%) | 56 (8.2%) | 13 (1.9%); 3 (0.4%); 6 (0.9%) |
| Sokoliuk 2021 (covert processing) | 595 | 163 (27.4%) | 261 (43.9%) | 105 (17.6%) | 49 (8.2%) | 11 (1.8%); 2 (0.3%); 4 (0.7%) |

Table S6C. Prolonged-DoC cohort. Clinical data are from the table released with the recordings and are available for 30 of the 38 patients. CRS-R indicates Coma Recovery Scale-Revised; VS/UWS, vegetative state or unresponsive wakefulness syndrome; MCS, minimally conscious state.

| **Characteristic** | **Value** |
|---|---|
| Files released / duplicate removed / development / analyzed | 42 / 1 / 3 / 38 (28 VS/UWS, 10 MCS) |
| Recording duration, median (IQR), hours | 6.0 (6.0-6.0); range 4.0-7.0 |
| Patients with linked clinical data, No. | 30 of 38 |
| Age, median (IQR) [range], years | 42 (37-57) [20-70] |
| Women, No. (%) | 14 (47) |
| Etiology, No. | Traumatic 14; anoxic 10; vascular 5; inflammatory 1 |
| Months since injury, median (IQR) [range] | 4 (3-5) [2-22] |
| CRS-R total, median (IQR) [range] | 6.5 (5-8) [2-17] |
| Outcome in the clinical table, No. | Persistent DoC 16; recovered consciousness 10; died 4 |

Table S7A. False-call rates under the primary reading, by analysis and dataset. Every positive call is false. The call rule column states whether the publication fully states the call criterion and threshold. Holm adjustment is within each clinical EEG null set (no-command: 7 motor analyses; no-stimulus: 2 ERP analyses). A single family of 9 gives Claassen 2019 Holm-adjusted P = .26 (primary), .30 (generous) and .22 (strict); no conclusion changes. The prolonged-DoC rows are estimates and carry no test. The originally planned reading is the planned lowest-rate reading on the original rows. Motor imagery: 8/550 here (first window) and 8/559 in Table S23 and in Table S9A in Supplementary Data S2, which use every patient with at least one usable window. Where no call occurred, the bootstrap interval is degenerate and the exact interval for 0 of the runs or patients analyzed is shown.

| Analysis | Dataset | Call rule | Primary reading, No. called/No. analyzed | Primary reading, % (95% CI) | Cross-fitted, % (95% CI) | Holm-adjusted P (cross-fitted) | Originally planned reading, No. called/No. analyzed; % (95% CI) |
|---|---|---|---|---|---|---|---|
| Claassen 2019 | Clinical EEG | Stated | 33/481 | 6.9 (4.8-9.5) | 6.9 (4.8-9.2) | 0.217 | 33/481; 6.9 (4.8-9.5) |
| Goldfine 2011 | Clinical EEG | Not stated | 164/703 | 23.3 (20.2-26.6) | 23.3 (20.2-26.5) | <0.001 | 268/703; 38.1 (34.5-41.8) |
| Cruse 2011 | Clinical EEG | Not fully stated | 1/147 | 0.7 (0.0-3.7) | 0.6 (0.0-1.9) | 1.000 | 0/26; 0.0 (0.0-13.2) |
| Cruse 2012 | Clinical EEG | Stated | 5/210 | 2.4 (0.8-5.5) | 2.8 (0.9-5.2) | 1.000 | 11/439; 2.5 (1.3-4.4) |
| Edlow 2017 | Clinical EEG | Stated | 3/796 | 0.4 (0.1-1.1) | 0.5 (0.1-1.1) | 1.000 | 3/796; 0.4 (0.1-1.1) |
| Guger 2017 | Clinical EEG | Stated | 8/550 | 1.5 (0.6-2.8) | 1.5 (0.5-2.6) | 1.000 | 8/550; 1.5 (0.6-2.8) |
| du Bois 2026 | Clinical EEG | Stated | 157/707 | 22.2 (19.2-25.5) | 22.3 (19.3-25.5) | <0.001 | 36/707; 5.1 (3.6-7.0) |
| Guger 2018 | Clinical EEG | Stated | 117/850 | 13.8 (11.5-16.3) | 15.3 (11.6-17.8) | <0.001 | 40/838; 4.8 (3.4-6.4) |
| Chennu 2013 | Clinical EEG | Not stated | - | No consistent reading fit the recordings | - | - | 1/467; 0.2 (0.0-1.2) |
| Gibson 2016 | Clinical EEG | Not stated | 25/683 | 3.7 (2.4-5.4) | 4.1 (2.4-5.7) | 0.881 | 25/683; 3.7 (2.4-5.4) |

| Analysis | Dataset | Call rule | Primary reading, No. called/No. analyzed | Primary reading, % (95% CI) | Cross-fitted, % (95% CI) | Holm-adjusted P (cross-fitted) | Originally planned reading, No. called/No. analyzed; % (95% CI) |
|---|---|---|---|---|---|---|---|
| Sokoliuk 2021 (covert processing) | Clinical EEG | Not stated | 41/565 | 7.3 (5.3-9.7) | 8.1 (5.1-11.2) | - | 43/595; 7.2 (5.3-9.6) |
| Goldfine 2011 | Prolonged DoC | Not stated | 3/38 | 7.9 (1.7-21.4) | 18.4 (0.0-34.2) | - | 6/38; 15.8 (6.0-31.3) |
| Cruse 2011 | Prolonged DoC | Not fully stated | 0/38 | 0.0 (0.0-9.3) | 2.6 (0.0-8.8) | - | 0/38; 0.0 (0.0-9.3) |
| Cruse 2012 | Prolonged DoC | Stated | 0/37 | 0.0 (0.0-9.5) | 0.0 (0.0-9.5) | - | 0/37; 0.0 (0.0-9.5) |
| Edlow 2017 | Prolonged DoC | Stated | 0/38 | 0.0 (0.0-9.3) | 0.0 (0.0-9.3) | - | 0/38; 0.0 (0.0-9.3) |
| Guger 2017 | Prolonged DoC | Stated | 0/36 | 0.0 (0.0-9.7) | 0.0 (0.0-9.7) | - | 0/32; 0.0 (0.0-10.9) |
| du Bois 2026 | Prolonged DoC | Stated | 9/38 | 23.7 (11.4-40.2) | 23.7 (10.8-37.5) | - | 4/38; 10.5 (2.9-24.8) |
| Guger 2018 | Prolonged DoC | Stated | 4/36 | 11.1 (3.1-26.1) | 25.0 (2.3-39.6) | - | 1/35; 2.9 (0.1-14.9) |
| Chennu 2013 | Prolonged DoC | Not stated | 0/38 | 0.0 (0.0-9.3) | 0.0 (0.0-9.3) | - | 0/38; 0.0 (0.0-9.3) |
| Gibson 2016 | Prolonged DoC | Not stated | 2/38 | 5.3 (0.6-17.7) | 7.9 (0.0-17.0) | - | 2/38; 5.3 (0.6-17.7) |

| Analysis | Dataset | Call rule | Primary reading, No. called/No. analyzed | Primary reading, % (95% CI) | Cross-fitted, % (95% CI) | Holm-adjusted P (cross-fitted) | Originally planned reading, No. called/No. analyzed; % (95% CI) |
|---|---|---|---|---|---|---|---|
| Sokoliuk 2021 (covert processing) | Prolonged DoC | Not stated | 1/37 | 2.7 (0.1-14.2) | 2.6 (0.0-8.3) | - | 1/37; 2.7 (0.1-14.2) |

Table S8A. Shortened-to-fit readings. These readings keep the design of the publication but shorten its length to fit a 30-minute window; they are never eligible for the primary reading or the Holm family. They are reported for the clinical EEG, where the native length did not fit.

| Analysis | Dataset | Shortened reading selected | No. called/No. analyzed | % (95% CI) | Cross-fitted, % (95% CI) | Range over shortened readings, % (selected, not an estimate) |
|---|---|---|---|---|---|---|
| Cruse 2011 | Clinical EEG | Cruse 2011 rule; 3 blocks per hand movement type; squared signal with a 250-ms boxcar; baseline interval 99.9% (shortened to fit) | 7/347 | 2.0 (0.8-4.1) | 2.9 (0.9-5.2) | 2.0-8.8 (6 readings) |
| Cruse 2012 | Clinical EEG | Contiguous cross-validation groups; per-class variance; 4 blocks (shortened to fit) | 12/439 | 2.7 (1.4-4.7) | 2.7 (1.4-5.2) | 2.7-3.6 (2 readings) |
| Chennu 2013 | Clinical EEG | P3b cluster against baseline, temporal cluster test and explicit-versus-implicit contrast; minimum cluster 40 ms; variance-based epoch rejection (z above 3); 12 blocks (shortened to fit) | 2/467 | 0.4 (0.1-1.5) | 0.4 (0.0-1.1) | 0.4-0.4 (2 readings) |

Table S8B. Covert-processing analysis (Sokoliuk 2021). The surrogate-test rate is a false covert-processing call. The prognostic cut-off is not a per-recording call rule, so its row is the share exceeding it. This analysis is never pooled with the command-following analyses.

| Dataset | Primary surrogate-test reading, No. called/No. analyzed; % (95% CI) | Cross-fitted, % (95% CI) | Prognostic cut-off of 0.116 exceeded (a priori default), No.; % (95% CI) | Range over cut-off readings, % | Range over surrogate-test readings, % |
|---|---|---|---|---|---|
| Clinical EEG | 41/565; 7.3 (5.3-9.7) | 8.1 (5.1-11.2) | 136/595; 22.9 (19.5-26.4) | 22.9-53.6 (4 readings) | 7.2-17.1 (24 readings) |
| Prolonged DoC | 1/37; 2.7 (0.1-14.2) | 2.6 (0.0-8.3) | 9/38; 23.7 (11.4-40.2) | 23.7-44.7 (4 readings) | 2.7-26.3 (24 readings) |

Table S9B. Claassen 2019 two-recording bracket. The true rate for a patient assessed twice is expected to lie between the two when calls are positively associated within a patient; because the rule corrects across recordings, the bracket is approximate. Its independent-days end uses other patients' windows as second recordings, so it does not measure within-patient variation between days.

| Claassen 2019 two-recording rule | Patients, No. | Positive on at least 1 of 2, % | 95% CI |
|---|---|---|---|
| Two independent recordings (upper bound) | 481 | 8.6 | 5.4-12.2 (patient-level bootstrap) |
| Two windows of one session (lower bound) | 14 | 7.1 | 0.2-33.9 (exact) |

Table S9C. Accumulation of Guger 2018 over repeated runs: patients with at least 1 positive run among the first k runs, among patients with at least k usable runs. The published range extends to 7 runs per patient; larger k is descriptive. Fixed cohort:

the 639 clinical EEG patients with at least 7 usable runs, read at every k, so the population does not change with k.

| Analysis | Dataset | Runs counted (k) | Patients with at least k runs, No. | Positive on at least 1 run, % (95% CI) | Status |
|---|---|---|---|---|---|
| Guger 2018 | Clinical EEG | 1 | 883 | 13.8 (11.6-16.3) | Within published range |
| Guger 2018 | Clinical EEG | 3 | 790 | 36.1 (32.7-39.5) | Within published range |
| Guger 2018 | Clinical EEG | 5 | 698 | 52.6 (48.8-56.3) | Within published range |
| Guger 2018 | Clinical EEG | 7 | 639 | 62.9 (59.0-66.7) | Within published range |
| Guger 2018 | Clinical EEG | 10 | 266 | 75.2 (69.5-80.3) | Beyond published range (descriptive) |
| Guger 2018 | Clinical EEG, fixed cohort | 1 | 639 | 14.1 (11.5-17.0) | Within published range |
| Guger 2018 | Clinical EEG, fixed cohort | 2 | 639 | 24.4 (21.1-27.9) | Within published range |
| Guger 2018 | Clinical EEG, fixed cohort | 3 | 639 | 35.8 (32.1-39.7) | Within published range |
| Guger 2018 | Clinical EEG, fixed cohort | 5 | 639 | 51.8 (47.8-55.7) | Within published range |
| Guger 2018 | Clinical EEG, fixed cohort | 7 | 639 | 62.9 (59.0-66.7) | Within published range |
| Guger 2018 | Prolonged DoC | 1 | 38 | 10.5 (2.9-24.8) | Within published range |
| Guger 2018 | Prolonged DoC | 3 | 38 | 31.6 (17.5-48.7) | Within published range |
| Guger 2018 | Prolonged DoC | 5 | 38 | 47.4 (31.0-64.2) | Within published range |
| Guger 2018 | Prolonged DoC | 7 | 38 | 60.5 (43.4-76.0) | Within published range |
| Guger 2018 | Prolonged DoC | 10 | 38 | 71.1 (54.1-84.6) | Beyond published range (descriptive) |
| Guger 2018 | Prolonged DoC | 20 | 38 | 84.2 (68.7-94.0) | Beyond published range (descriptive) |
| Guger 2018 | Prolonged DoC | 40 | 37 | 100.0 (90.5-100.0) | Beyond published range (descriptive) |

| Analy sis | Dataset | Runs counte d (k) | Patients with at least k runs, No. | Positive on at least 1 run, % (95% CI) | Status |
|---|---|---|---|---|---|
| Guger 2018 | Prolonged DoC | 80 | 37 | 100.0 (90.5-100.0) | Beyond published range (descriptive) |
| Guger 2018 | Prolonged DoC | 120 | 26 | 100.0 (86.8-100.0) | Beyond published range (descriptive) |

Table S9D. Per-patient rules at k = 2 under the generous and strict readings.

| Sensitivity reading | Analysis | Dataset | k | Reading | No. positive/No. analyzed | % (95% CI) |
|---|---|---|---|---|---|---|
| Generous | du Bois 2026 | Clinical EEG | 2 | Assessment runs; block-wise cross-validation; call: label permutation; per-fold maxima | 9/95 | 9.5 (4.4-17.2) |
| Generous | Guger 2018 | Clinical EEG | 2 | 3-stimulus paradigm; Murovec 2020 stimulus timing; 8-segment features; median accuracy summary; threshold of 23% | 206/841 | 24.5 (21.6-27.5) |
| Generous | du Bois 2026 | Prolonged DoC | 2 | Assessment runs; block-wise cross-validation; call: label permutation; per-fold maxima | 6/38 | 15.8 (6.0-31.3) |
| Generous | Guger 2018 | Prolonged DoC | 2 | 3-stimulus paradigm; Annen 2018 stimulus timing; 8-segment features; median accuracy summary; threshold of 23% | 8/38 | 21.1 (9.6-37.3) |
| Strict | du Bois 2026 | Clinical EEG | 2 | Assessment runs; block-wise cross-validation; call: peak accuracy above 70% or paired t test; fold-mean peak times (corrected) | 30/95 | 31.6 (22.4-41.9) |
| Strict | Guger 2018 | Clinical EEG | 2 | 3-stimulus paradigm; Murovec 2020 stimulus timing; 8-segment features; median accuracy summary; threshold of 23% | 206/841 | 24.5 (21.6-27.5) |
| Strict | du Bois 2026 | Prolonged DoC | 2 | Assessment runs; block-wise cross-validation; call: peak accuracy above 70% or paired t test; fold-mean peak times (corrected) | 18/38 | 47.4 (31.0-64.2) |

| Sensitivity reading | Analysis | Dataset | k | Reading | No. positive/No. analyzed | % (95% CI) |
|---|---|---|---|---|---|---|
| Strict | Guger 2018 | Prolonged DoC | 2 | 3-stimulus paradigm; Murovec 2020 stimulus timing; 8-segment features; median accuracy summary; threshold of 23% | 9/38 | 23.7 (11.4-40.2) |

Table S11D. Goldfine 2011 planned lowest-rate reading on EEG-like noise by amplitude fluctuation and correlation time of the envelope (calls/60 recordings).

| Amplitude fluctuation (log scale) | 1 s | 5 s | 10 s | 30 s | 60 s |
|---|---|---|---|---|---|
| SD 0.25 | 5/60 | 9/60 | 13/60 | 2/60 | 1/60 |
| SD 0.5 | 16/60 | 20/60 | 27/60 | 12/60 | 7/60 |
| SD 0.75 | 25/60 | 23/60 | 30/60 | 11/60 | 7/60 |

Table S11E. Atlas implementation bar: the planned lowest-rate reading should lie within the binomial 95% range of 5% on white noise. Guger 2018 and du Bois 2026 analyses were declared exceptions in advance.

| **Analysis (planned lowest-rate reading)** | **White-noise recordings called, No./No. analyzed** | **Binomial 95% range of 5%, No. called** | **Within range** | **Pre-declared exception** |
|---|---:|---:|---|---|
| Claassen 2019 | 1/30 | 0-4 | Yes | No |
| Goldfine 2011 | 0/60 | 0-7 | Yes | No |
| Cruse 2011 | 1/60 | 0-7 | Yes | No |
| Cruse 2012 | 7/60 | 0-7 | Yes | No |
| Edlow 2017 | 0/60 | 0-7 | Yes | No |
| Guger 2017 | 0/60 | 0-7 | Yes | No |
| du Bois 2026 | 0/30 | 0-4 | Yes | Yes |
| Guger 2018 | 2/60 | 0-7 | Yes | Yes |
| Chennu 2013 | 0/60 | 0-7 | Yes | No |
| Gibson 2016 | 1/60 | 0-7 | Yes | No |
| Sokoliuk 2021 (covert processing) | 3/60 | 0-7 | Yes | No |

Table S11G. Diagnostic of the Cruse 2012 white-noise excess, on synthetic noise only. Two cross-validation readings of Cruse 2012 are shown (groups drawn at random within each class; contiguous groups). P tests whether the rate exceeds 5% (one-sided exact binomial). The rate on the clinical recordings is shown for comparison and differed from the 500-draw white-noise rate (Fisher P = .006).

| **Condition** | **Cross-validation groups** | **Calls/draws** | **Rate, % (95% CI)** | **P (rate above 5%)** |
|---|---|---:|---:|---:|
| White noise, 500 draws | random groups | 38/500 | 7.6 (5.4-10.3) | 0.008 |
| White noise, 500 draws | contiguous groups | 35/500 | 7.0 (4.9-9.6) | 0.030 |
| Pink (1/f) noise, same 500 draws | random groups | 38/500 | 7.6 (5.4-10.3) | 0.008 |

| Condition | Cross-validation groups | Calls/draws | Rate, % (95% CI) | P (rate above 5%) |
|---|---|---|---|---|
| Pink (1/f) noise, same 500 draws | contiguous groups | 37/500 | 7.4 (5.3-10.1) | 0.012 |
| White noise, first 353 draws: the analysis's own null | random groups | 25/353 | 7.1 (4.6-10.3) | 0.053 |
| White noise, first 353 draws: the analysis's own null | contiguous groups | 23/353 | 6.5 (4.2-9.6) | 0.120 |
| White noise, first 353 draws: null permutes labels within each cross-validation fold | random groups | 12/353 | 3.4 (1.8-5.9) | 0.941 |
| White noise, first 353 draws: null permutes labels within each cross-validation fold | contiguous groups | 15/353 | 4.2 (2.4-6.9) | 0.775 |
| White noise, first 353 draws: groups re-stratified for each permutation | random groups | 17/353 | 4.8 (2.8-7.6) | 0.597 |
| White noise: trials cut to the clinical trial counts (347 evaluable draws) | random groups | 21/347 | 6.1 (3.8-9.1) | 0.214 |
| Same, with groups re-stratified for each permutation | random groups | 11/347 | 3.2 (1.6-5.6) | 0.962 |
| Clinical EEG (primary reading) | random groups | 5/210 | 2.4 (0.8-5.5) | - |

Table S11H. Calibration of the permutation P values of the Cruse 2012 random-group reading on 200 white-noise draws (1,000 permutations per comparison). D is the Kolmogorov-Smirnov distance from a uniform distribution. Under a calibrated null each share would equal its threshold. Observed accuracy exceeded the mean of its permutation null by 0.0097 (SE, 0.0010). The mean absolute deviation of a test fold's class share from the overall share was 0.000 for observed labels and 0.107 for permuted labels (random groups), and 0.071 and 0.107 (contiguous groups).

| P value | P values, No. | D | Below .05, % | Below .01, % | Below .001, % |
|---|---|---|---|---|---|
| Right tail | 200 | 0.186 | 9.00 | 0.50 | 0.50 |
| Left tail | 200 | 0.211 | 7.00 | 1.00 | 0.00 |
| Both tails pooled | 400 | 0.185 | 8.00 | 0.75 | 0.25 |
| Analysis's own P (smaller of the two) | 200 | 0.436 | 15.50 | 1.50 | 0.50 |

Table S13D. Label-shuffled and true-responder counts of the published default readings, original assembly against one source trial per slot.

| Analysis | Shuffled calls, original assembly | Shuffled calls, one source per slot | True responders, original assembly | True responders, one source per slot |
|---|---|---|---|---|
| Claassen 2019 | 5/20 (25.0) | 2/20 (10.0) | 20/20 | 20/20 |
| Edlow 2017 | 14/51 (27.5) | 3/51 (5.9) | 50/51 | 50/51 |
| Guger 2017 | 6/15 (40.0) | 2/15 (13.3) | 7/14 | 3/13 |
| du Bois 2026 | 26/51 (51.0) | 18/51 (35.3) | 47/51 | 45/51 |
| Goldfine 2011 | 51/51 (100.0) | 51/51 (100.0) | 51/51 | 51/51 |

Table S13E. Detection range across all readings of an analysis on the original assembly, primary arm.

| Analysis | Readings, No. | Detection range over readings, % | Readings with upper bound below the published rate, No. |
|---|---|---|---|
| Claassen 2019 | 1 | 100.0-100.0 | 0 |
| Goldfine 2011 | 16 | 100.0-100.0 | 0 |
| Guger 2017 | 16 | 13.0-100.0 | 15 |
| du Bois 2026 | 12 | 51.0-100.0 | 7 |
| Edlow 2017 | 8 | 98.0-100.0 | 0 |
| Cruse 2012 | 4 | 91.5-95.7 | 0 |
| Guger 2018 | 120 | 53.8-100.0 | 110 |
| Chennu 2013 | 32 | 7.7-100.0 | 28 |
| Gibson 2016 | 16 | 0.0-92.3 | 12 |

Table S14A. False-call control of the randomization-based versions. The bar is a pooled point estimate of 5% or less with no Holm-significant excess in any dataset, background stratum or recording unit. A point estimate above 5% with no significant excess is labelled not met on the point estimate only.

| **Analysis** | **Randomized overlays, No.** | **Pooled false-call rate, %** | **Clinical EEG, %** | **Prolonged DoC, %** | **Holm-significant excesses, No.** | **Smallest Holm-adjusted P** | **False-call bar** |
|---|---|---|---|---|---|---|---|
| Claassen 2019 | 481 | 4.4 | 4.4 | - | 0 | 0.767 | Met |
| Goldfine 2011 | 1,860 | 4.7 | 4.6 | 5.0 | 0 | 1.000 | Met |
| Cruse 2011 | 1,681 | 4.3 | 4.2 | 4.5 | 0 | 0.958 | Met |
| Cruse 2012 | 867 | 3.9 | 3.6 | 5.0 | 0 | 1.000 | Met |
| Edlow 2017 | 1,024 | 5.3 | 5.2 | 5.7 | 0 | 0.708 | Not met (point estimate above 5%) |
| Guger 2017 | 8,252 | 4.8 | 5.1 | 4.4 | 0 | 0.470 | Met |
| du Bois 2026 | 2,522 | 5.1 | 5.5 | 4.3 | 0 | 0.297 | Not met (point estimate above 5%) |
| Guger 2018 | 42,896 | 4.9 | 4.9 | 5.0 | 0 | 0.686 | Met |
| Chennu 2013 | 695 | 5.5 | 6.0 | 4.4 | 0 | 0.250 | Not met (point estimate above 5%) |
| Gibson 2016 | 909 | 5.3 | 5.3 | 5.3 | 0 | 0.793 | Not met (point estimate above 5%) |
| Sokoliuk 2021 (covert processing) | 823 | 4.6 | 3.9 | 6.6 | 0 | 0.342 | Met |

Table S14C. Uncorrected against size-corrected comparators for the two analyses with the largest false-call excess. The uncorrected comparison is inflated.

| Analysis | Uncorrected comparator, % detected (range over 4 points) | Size-corrected comparator, % detected (range over 4 points) |
|---|---|---|
| Goldfine 2011 | 91.6-98.5 | 42.0-82.5 |
| du Bois 2026 | 100.0-100.0 | 81.1-99.5 |

Table S15B. Sensitivity analyses of the planned alignment test (M1). One-sided P for an odds ratio above 1. The point estimate is positive in every analysis; these are sensitivities, not tests.

| Model | OR per SD of X (95% CI) | One-sided P | Rows / patients | Readings |
|---|---|---|---|---|
| M1 (primary) | 1.06 (0.98-1.15) | 0.064 | 157,578 / 900 | 233 |
| X from the whole recording | 1.02 (0.94-1.11) | 0.321 | 157,597 / 900 | 233 |
| X from the module's own bands | 1.05 (0.96-1.14) | 0.114 | 157,026 / 900 | 233 |
| X from the first principal component across channels | 1.08 (1.01-1.16) | 0.015 | 156,358 / 900 | 233 |
| Post-freeze recordings only | 1.04 (0.94-1.14) | 0.238 | 82,148 / 469 | 233 |
| Without the Claassen 2019 rows | 1.10 (1.03-1.17) | 0.006 | 157,100 / 900 | 232 |
| Activation-clean windows | 1.08 (1.00-1.16) | 0.027 | 153,736 / 896 | 233 |
| Flat failed channel entered as X = 0 | 1.06 (0.98-1.15) | 0.081 | 157,768 / 900 | 233 |
| Artifact mask | 1.10 (1.01-1.20) | 0.018 | 157,088 / 900 | 233 |
| Random intercept (default reading, 1 row per patient and analysis) | 1.17 (1.08-1.28) | - | 5,415 / 900 | 9 analyses |

Table S15C. Models of the background test (S2) and the artifact test (S3). Odds ratios of a false call, with and without X in the model; Holm-adjusted P for S2 is .008 and for S3 .657.

| Model (odds ratio of a false call) | OR (95% CI) | Two-sided P |
|---|---|---|
| S2: organized backgrounds, before X | 1.50 (1.22-1.83) | 0.002 |
| S2: organized backgrounds, rows with X | 1.67 (1.36-2.00) | 0.001 |
| S2: organized backgrounds, after X | 1.67 (1.35-2.01) | 0.001 |
| S2: X after X is in the model | 1.03 (0.94-1.13) | 0.481 |
| S2: transitioning windows, before X | 1.05 (0.85-1.30) | 0.668 |
| S3: artifact share, per 10 percentage points, before X | 1.01 (0.99-1.03) | 0.219 |

Table S15D. Exploratory estimates in the prolonged-DoC recordings. Level-of-consciousness contrast (S4): patient-median X by diagnosis, with resting markers on the same windows; the three analyses are overall, wake-like windows only, and adjusted for state. Arousal and recovery estimates (A3): 30 patients (10 recovered, 4 died); the study was not powered for prognosis and no inference about prognosis is drawn.

| S4 analysis | Patient-median X, MCS (10) against VS/UWS (28) | AUROC, resting markers | AUROC, markers plus X | Change in AUROC (95% CI) | AUROC, X alone |
|---|---|---|---|---|---|
| Overall | Hodges-Lehmann shift, MCS minus VS/UWS: -0.008 (-0.244 to 0.199) | 0.639 | 0.632 | -0.007 (-0.121 to 0.182) | 0.496 |
| Wake-like windows | Hodges-Lehmann shift: -0.054 (-0.254 to 0.160) | 0.629 | 0.604 | -0.025 (-0.125 to 0.125) | 0.436 |
| Adjusted for state | Coefficient of MCS: 0.016 (-0.200 to 0.251) | 0.571 | 0.571 | 0.000 (-0.121 to 0.189) | 0.496 |

| A3 exploratory estimate (not powered for prognosis; no inference about prognosis is drawn) | Estimate (95% CI) | Patients, No. |
|---|---|---|
| Spearman correlation with CRS-R arousal subscale | 0.062 (-0.266 to 0.379) | 30 |
| Same, adjusted for MCS | 0.098 (-0.246 to 0.429) | 30 |
| AUROC for recovery | 0.470 (0.254 to 0.684) | 30 |
| Same, adjusted for MCS | 0.470 (0.212 to 0.737) | 30 |
| AUROC for recovery, deaths excluded | 0.494 (0.255 to 0.733) | 26 |
| Same, adjusted for MCS | 0.487 (0.186 to 0.781) | 26 |

Table S15E. Label-aligned heart-rate changes and false calls (exploratory). Exposed designs are those not typed immune. A circular-shift placebo moves the label schedule within the same recording. Contrasts are ratios of odds ratios with shared patient draws.

| Set (odds ratio per SD of label-aligned heart-rate change) | Rows / recordings | Aligned OR (95% CI) | Placebo OR (95% CI) |
|---|---|---|---|
| Exposed designs, aligned only | 20,360 / 607 | 1.11 (1.02-1.21) | - |
| Exposed, aligned and placebo | 13,395 / 557 | 1.19 (1.08-1.32) | 1.06 (0.96-1.17) |
| Immune designs, aligned only | 9,394 / 569 | 0.96 (0.88-1.04) | - |
| Immune, aligned and placebo | 7,380 / 501 | 0.97 (0.87-1.07) | 1.02 (0.92-1.12) |
| Randomization-based versions, aligned only | 20,356 / 607 | 1.08 (0.91-1.27) | - |
| Randomization-based, aligned and placebo | 13,945 / 553 | 1.03 (0.81-1.30) | 1.03 (0.78-1.31) |
| Exposed, stable background state | 15,354 / 538 | 1.08 (0.98-1.19) | - |
| Exposed, stable state, aligned and placebo | 10,051 / 486 | 1.15 (1.03-1.31) | 1.06 (0.95-1.18) |
| Exposed, with 45-58 Hz muscle activity | 20,360 / 607 | 1.12 (1.03-1.21) | - |
| Exposed, placebo and muscle activity | 13,395 / 557 | 1.19 (1.08-1.34) | 1.06 (0.97-1.17) |
| Exposed, activation-clean windows | 19,384 / 598 | 1.12 (1.03-1.23) | - |

| Contrast (ratio of odds ratios, shared patient draws) | Ratio (95% CI) | P |
|---|---|---|
| Aligned minus placebo, exposed | 1.13 (0.97-1.31) | 0.101 |
| Exposed minus randomized, same rows | 1.03 (0.86-1.25) | 0.723 |
| Exposed minus immune | 1.16 (1.03-1.31) | 0.019 |

Table S16A. False-call rates in post-arrest coma (490 patients, first overlay; every positive call is false). Primary: the stated-rule reading selected on these rows; originally planned: the lowest-rate reading selected on these rows; default: the a priori default reading. Exact 95% CIs; cross-fitted rates with patient-bootstrap CIs. Randomization-based: the randomized version of the analysis pooled over every overlay, as on clinical EEG. Primary readings: estimation only (no Holm adjustment); randomization-based rows were tested against the false-call bar as on clinical EEG (Supplementary Results 10).

| **Analysis** | **Primary reading, No./No.; % (95% CI)** | **Cross-fitted, % (95% CI)** | **Originally planned reading, No./No.; % (95% CI)** | **A priori default, No./No.; % (95% CI)** | **Randomization-based, No./No.; %** |
|---|---|---|---|---|---|
| Claassen 2019 | 40/489; 8.2 (5.9-11.0) | 8.3 (5.9-10.9) | 40/489; 8.2 (5.9-11.0) | 40/489; 8.2 (5.9-11.0) | 25/489; 5.1 |
| Goldfine 2011 | 54/446; 12.1 (9.2-15.5) | 12.0 (9.0-20.8) | 79/428; 18.5 (14.9-22.5) | 366/446; 82.1 (78.2-85.5) | 61/886; 6.9 |
| Cruse 2011 | No consistent reading fits the 30-min window | - | 9/463; 1.9 (0.9-3.7) | No rows | 30/924; 3.2 |
| Cruse 2012 | No consistent reading fits the 30-min window | - | 21/474; 4.4 (2.8-6.7) | No rows | 18/472; 3.8 |
| Edlow 2017 | 2/484; 0.4 (0.1-1.5) | 0.4 (0.0-2.8) | 2/484; 0.4 (0.1-1.5) | 50/484; 10.3 (7.8-13.4) | 24/484; 5.0 |
| Guger 2017 | 2/418; 0.5 (0.1-1.7) | 0.5 (0.0-1.2) | 1/403; 0.2 (0.0-1.4) | 64/403; 15.9 (12.5-19.8) | 120/2805; 4.3 |
| du Bois 2026 | 129/464; 27.8 (23.8-32.1) | 27.9 (24.0-32.0) | 8/464; 1.7 (0.7-3.4) | 306/464; 65.9 (61.4-70.3) | 43/927; 4.6 |
| Guger 2018 | 64/481; 13.3 (10.4-16.7) | 13.8 (10.2-17.6) | 20/481; 4.2 (2.6-6.3) | 85/481; 17.7 (14.4-21.4) | 997/19082; 5.2 |

| Analysis | Primary reading, No./No.; % (95% CI) | Cross-fitted, % (95% CI) | Originally planned reading, No./No.; % (95% CI) | A priori default, No./No.; % (95% CI) | Randomization-based, No./No.; % |
|---|---|---|---|---|---|
| Chennu 2013 | No consistent reading fits the 30-min window | - | 0/489; 0.0 (0.0-0.8) | 0/489; 0.0 (0.0-0.8) | 29/489; 5.9 |
| Gibson 2016 | 22/483; 4.6 (2.9-6.8) | 4.6 (2.9-7.5) | 22/483; 4.6 (2.9-6.8) | 22/483; 4.6 (2.9-6.8) | 25/483; 5.2 |
| Sokoliuk 2021 (covert processing) | 32/439; 7.3 (5.0-10.1) | 7.3 (5.0-11.1) | 35/439; 8.0 (5.6-10.9) | 107/489; 21.9 (18.3-25.8) | 25/489; 5.1 |

Table S16C. The planned post-arrest test: good outcome (Cerebral Performance Category 1-2) and the mechanism measure X on the fixed 12-block schedule, adjusted as in Supplementary Methods 14. Odds ratio (OR) per SD of X with a patient-bootstrap 95% CI (2,000 resamples).

| Model | No. of patients | OR per SD of X (95% CI) | P |
|---|---|---|---|
| Primary (continuity grade) | 490 | 0.79 (0.59-1.03) | .10 |
| Without the 40 patients of an earlier null set | 452 | 0.75 (0.54-0.98) | .04 |
| Foundation-model background labels | 490 | 0.91 (0.69-1.18) | .47 |
| Without patients with a flat channel | 355 | 0.92 (0.64-1.29) | .64 |
| Leaving out hospital A | 286 | 1.01 (0.67-1.56) | .97 |
| Leaving out hospital B | 385 | 0.89 (0.61-1.25) | .50 |
| Leaving out hospital D | 438 | 0.77 (0.55-1.04) | .08 |
| Leaving out hospital E | 424 | 0.76 (0.55-1.02) | .07 |
| Leaving out hospital F | 427 | 0.68 (0.49-0.90) | .009 |

Change in leave-one-hospital-out AUROC when X was added to the background-only model: 0.722 to 0.731 (change, 0.009; 95% CI, -0.002 to 0.020). Patients outside the model (60 without a window and 54 without complete data) had good outcome in 40 of 114 (35.1%) against 185 of 490 (37.8%) in the model (Fisher exact P = .67).

Calibration of the clinical EEG mechanism model in coma: intercept -0.31 (95% CI, -0.38 to -0.24), slope 0.81 (0.77-0.85).

Table S16D. False call and good outcome, with the adjustment of Table S16C without X (OR of good outcome for a false call; patient-bootstrap 95% CI). A priori default: each analysis's a priori default reading. Primary: the primary reading of Table S16A. Estimates only, never ranked, no multiplicity adjustment.

| Analysis | A priori default: false calls/patients | A priori default: OR (95% CI) | Primary: false calls/patients | Primary: OR (95% CI) |
|---|---|---|---|---|
| Claassen 2019 | 40/489 | 0.41 (0.12-1.20) | 40/489 | 0.41 (0.12-1.18) |
| Goldfine 2011 | 366/446 | 0.78 (0.32-1.64) | 54/446 | 0.69 (0.22-1.96) |
| Cruse 2011 | - | Default reading has no post-arrest rows | - | No consistent reading |
| Cruse 2012 | - | Default reading has no post-arrest rows | - | No consistent reading |
| Edlow 2017 | 50/484 | 0.72 (0.25-1.90) | 2/484 | Not estimable |
| Guger 2017 | 64/403 | 1.29 (0.63-2.78) | 2/418 | Not estimable |
| du Bois 2026 | 306/464 | 0.53 (0.28-0.96) | 129/464 | 1.81 (1.04-3.47) |
| Guger 2018 | 85/481 | 0.78 (0.38-1.48) | 64/481 | 0.74 (0.32-1.52) |
| Chennu 2013 | 0/489 | Not estimable | - | No consistent reading |
| Gibson 2016 | 22/483 | 2.06 (0.68-9.01) | 22/483 | 2.06 (0.68-9.62) |
| Sokoliuk 2021 (covert processing) | 107/489 | 0.80 (0.39-1.55) | 32/439 | 0.51 (0.10-2.10) |

Table S16E. False calls by design type (planned estimate), pooled over every reading of each analysis, including liberal departures, with the clinical EEG row weights (patient-bootstrap 95% CI). The pooled rates are not rates of any published method; the immune designs are the du Bois 2026 training readings and the motor imagery analysis, so that row is dominated by the du Bois 2026 training readings.

| **Design type** | **Rows** | **Patients** | **% (95% CI)** |
|---|---:|---:|---:|
| Inflated | 10,350 | 489 | 19.9 (19.1-20.7) |
| Immune | 9,294 | 478 | 54.9 (53.8-56.0) |
| Deflated | 80,305 | 490 | 17.9 (17.2-18.7) |

Table S16F. Slow EEG non-stationarity in post-arrest coma (secondary; Supplementary Results 10). Primary population: continuous or discontinuous background; full sample: every background. Heart-rate coupling: standardized beta of log non-stationarity on the heart-rate fluctuation, adjusted for hours since return of circulation and hospital (HC3 robust 95% CI). Outcome: OR of good outcome per SD of log non-stationarity, with the adjustment of Table S16C (patient-bootstrap 95% CI).

| **Estimate** | **Population** | **No. of patients** | **Estimate (95% CI)** | **P** |
|---|---|---|---|---|
| Heart-rate coupling, beta | Continuous or discontinuous | 140 | -0.04 (-0.22 to 0.14) | - |
| Heart-rate coupling, beta | Full sample | 298 | 0.05 (-0.08 to 0.18) | - |
| Good outcome, OR per SD | Continuous or discontinuous | 245 | 0.65 (0.37-1.01) | .06 |
| Good outcome, OR per SD | Full sample | 499 | 0.77 (0.55-1.05) | .10 |
| Better CPC (1-5), proportional-odds OR per SD | Continuous or discontinuous | 245 | 0.71 (0.50-1.01) | - |
| Good outcome, OR per SD, heart-rate fluctuation added (descriptive) | Continuous or discontinuous | 130 | 0.54 (0.13-1.19) | - |

Heart-rate fluctuation was unavailable for 246 of 544 patients (no ECG file, 186; low template correlation, 42; implausible RR intervals, 18).

Table S16G. False calls of each primary reading in post-arrest coma by the American Clinical Neurophysiology Society continuity grade of the window (descriptive). No./No.; % (exact 95% CI). Every positive call is false.

| Analysis | Continuous | Discontinuous | Burst suppression | Suppressed | All |
|---|---|---|---|---|---|
| Claassen 2019 | 9/132; 6.8 (3.2-12.5) | 12/108; 11.1 (5.9-18.6) | 12/174; 6.9 (3.6-11.7) | 7/75; 9.3 (3.8-18.3) | 40/489; 8.2 (5.9-11.0) |
| Goldfine 2011 | 10/124; 8.1 (3.9-14.3) | 17/101; 16.8 (10.1-25.6) | 22/157; 14.0 (9.0-20.4) | 5/64; 7.8 (2.6-17.3) | 54/446; 12.1 (9.2-15.5) |
| Edlow 2017 | 0/130; 0.0 (0.0-2.8) | 2/105; 1.9 (0.2-6.7) | 0/174; 0.0 (0.0-2.1) | 0/75; 0.0 (0.0-4.8) | 2/484; 0.4 (0.1-1.5) |
| Guger 2017 | 0/109; 0.0 (0.0-3.3) | 0/73; 0.0 (0.0-4.9) | 0/162; 0.0 (0.0-2.3) | 2/74; 2.7 (0.3-9.4) | 2/418; 0.5 (0.1-1.7) |
| du Bois 2026 | 40/126; 31.7 (23.7-40.6) | 23/104; 22.1 (14.6-31.3) | 40/162; 24.7 (18.3-32.1) | 26/72; 36.1 (25.1-48.3) | 129/464; 27.8 (23.8-32.1) |
| Guger 2018 | 17/128; 13.3 (7.9-20.4) | 12/103; 11.7 (6.2-19.5) | 24/174; 13.8 (9.0-19.8) | 11/76; 14.5 (7.5-24.4) | 64/481; 13.3 (10.4-16.7) |
| Gibson 2016 | 7/129; 5.4 (2.2-10.9) | 6/106; 5.7 (2.1-11.9) | 6/173; 3.5 (1.3-7.4) | 3/75; 4.0 (0.8-11.2) | 22/483; 4.6 (2.9-6.8) |
| Sokoliuk 2021 (covert processing) | 8/112; 7.1 (3.1-13.6) | 8/91; 8.8 (3.9-16.6) | 11/165; 6.7 (3.4-11.6) | 5/71; 7.0 (2.3-15.7) | 32/439; 7.3 (5.0-10.1) |

Table S16H. False calls of each primary reading in post-arrest coma by the American Clinical Neurophysiology Society continuity grade of the window, on all 544 patients with a usable window (descriptive; Table S16G gives the 490 with complete data). No./No.; % (exact 95% CI). Every positive call is false; patients without a grade count only in the total.

| Analysis | Continuous | Discontinuous | Burst suppression | Suppressed | All |
|---|---|---|---|---|---|
| Claassen 2019 | 11/146, 7.5 (3.8-13.1) | 12/119, 10.1 (5.3-17.0) | 13/192, 6.8 (3.7-11.3) | 7/86, 8.1 (3.3-16.1) | 43/543, 7.9 (5.8-10.5) |
| Goldfine 2011 | 10/135, 7.4 (3.6-13.2) | 18/110, 16.4 (10.0-24.6) | 24/174, 13.8 (9.0-19.8) | 5/75, 6.7 (2.2-14.9) | 57/494, 11.5 (8.9-14.7) |
| Edlow 2017 | 0/144, 0.0 (0.0-2.5) | 2/116, 1.7 (0.2-6.1) | 0/192, 0.0 (0.0-1.9) | 0/86, 0.0 (0.0-4.2) | 2/538, 0.4 (0.0-1.3) |
| Guger 2017 | 0/118, 0.0 (0.0-3.1) | 0/82, 0.0 (0.0-4.4) | 0/179, 0.0 (0.0-2.0) | 2/85, 2.4 (0.3-8.2) | 2/464, 0.4 (0.1-1.5) |
| du Bois 2026 | 42/138, 30.4 (22.9-38.8) | 23/113, 20.4 (13.4-29.0) | 46/179, 25.7 (19.5-32.8) | 29/83, 34.9 (24.8-46.2) | 140/513, 27.3 (23.5-31.4) |
| Guger 2018 | 18/141, 12.8 (7.7-19.4) | 14/114, 12.3 (6.9-19.7) | 28/192, 14.6 (9.9-20.4) | 15/87, 17.2 (10.0-26.8) | 75/534, 14.0 (11.2-17.3) |
| Gibson 2016 | 7/141, 5.0 (2.0-10.0) | 6/117, 5.1 (1.9-10.8) | 6/191, 3.1 (1.2-6.7) | 5/86, 5.8 (1.9-13.0) | 24/535, 4.5 (2.9-6.6) |
| Sokoliuk 2021 (covert processing) | 8/123, 6.5 (2.8-12.4) | 8/101, 7.9 (3.5-15.0) | 13/182, 7.1 (3.9-11.9) | 5/82, 6.1 (2.0-13.7) | 34/488, 7.0 (4.9-9.6) |

Table S17. Replication: the primary readings fixed on the development patients, applied unchanged to the clinical EEG of 730 further patients listed after the readings were fixed and never analyzed before; every positive call is false with respect to commands. Tests fixed before the draw: the 3 analyses above 5% in development, one-sided exact binomial for a rate above 5% (Holm over 3); the other 4, one-sided exact binomial for a rate below 10% (Holm over 4). Reproduced: the replication rate lies inside the 95% prediction interval around the development cross-fitted rate (the centre of the interval; the reading chosen on one half of the development patients and applied to the other). Development: primary reading, all development patients. Rerun with the replication's window placement (14 of the 481 windows had no fitting overlay), the Claassen 2019 development windows were called in 26/467 (5.6%; 95% CI, 3.7%-8.1%) against 33/467 at the original placement (exact McNemar P = .40) and 25/730 in the replication (Fisher P = .08); these data cannot tell whether window placement explains the difference.

| Analysis | Development, k/n, % (95% CI) | Development cross-fitted, k/n, % | Replication, k/n, % (95% CI) | Test | Holm P | 95% prediction interval, % | Reproduced |
|---|---|---|---|---|---|---|---|
| Claassen 2019 | 33/481, 6.9 (4.8-9.5) | 33/481, 6.9 | 25/730, 3.4 (2.2-5.0) | Below 10% | <.001 | 4.0-9.8 | No |
| Goldfine 2011 | 164/703, 23.3 (20.2-26.6) | 164/703, 23.3 | 165/722, 22.9 (19.8-26.1) | Above 5% | <.001 | 18.9-27.7 | Yes |
| Edlow 2017 | 3/796, 0.4 (0.1-1.1) | 4/796, 0.5 | 2/718, 0.3 (0.0-1.0) | Below 10% | <.001 | 0.0-1.2 | Yes |
| Guger 2017 | 8/550, 1.5 (0.6-2.8) | 8/550, 1.5 | 2/561, 0.4 (0.0-1.3) | Below 10% | <.001 | 0.0-2.9 | Yes |
| du Bois 2026 | 157/707, 22.2 (19.2-25.5) | 157/707, 22.2 | 181/730, 24.8 (21.7-28.1) | Above 5% | <.001 | 17.9-26.5 | Yes |
| Guger 2018 | 117/850, 13.8 (11.5-16.3) | 129/841, 15.3 | 84/670, 12.5 (10.1-15.3) | Above 5% | <.001 | 11.7-19.0 | Yes |
| Gibson 2016 | 25/683, 3.7 (2.4-5.4) | 28/683, 4.1 | 37/706, 5.2 (3.7-7.2) | Below 10% | <.001 | 2.0-6.2 | Yes |

| Analysis | Development, k/n, % (95% CI) | Development cross-fitted, k/n, % | Replication, k/n, % (95% CI) | Test | Holm P | 95% prediction interval, % | Reproduced |
|---|---|---|---|---|---|---|---|
| Guger 2018, two-run rule | 206/841, 24.5 (21.6-27.5) | - | 152/652, 23.3 (20.1-26.8) | - | - | - | - |

Table S18. The du Bois 2026 decision pathway on recordings without study commands. Advancement (the primary call): peak accuracy above 70% or a one-tailed paired t test P < .05, the published criterion for advancing to brain-computer interface training. Significant run: the t test alone, as the publication defines it. Permutation: 100 label permutations at the fold-mean peak time with the configuration fixed, P < .05, which the publication describes as an evaluation against chance without stating that it gates advancement. As a gate it is lenient: the observed accuracy is the best over time points and configurations, and each permuted accuracy is not. Every window's permutation and corrected rows matched on window start, peak time and peak accuracy.

| Dataset | Advancement | Significant run | Permutation | Advancement and permutation |
|---|---|---|---|---|
| Clinical EEG | 157/707, 22.2 (19.2-25.5) | 151/707, 21.4 (18.4-24.6) | 36/707, 5.1 (3.6-7.0) | 16/707, 2.3 (1.3-3.6) |
| Prolonged DoC | 9/38, 23.7 (11.4-40.2) | 8/38, 21.1 (9.6-37.3) | 4/38, 10.5 (2.9-24.8) | 3/38, 7.9 (1.7-21.4) |
| Post-arrest coma, 490 patients | 129/464, 27.8 (23.8-32.1) | 127/464, 27.4 (23.4-31.7) | 8/464, 1.7 (0.7-3.4) | 5/464, 1.1 (0.4-2.5) |
| Post-arrest coma, every patient with a window | 140/513, 27.3 (23.5-31.4) | 138/513, 26.9 (23.1-31.0) | 10/513, 1.9 (0.9-3.6) | 5/513, 1.0 (0.3-2.3) |
| Replication set | 181/730, 24.8 (21.7-28.1) | 177/730, 24.2 (21.2-27.5) | 43/730, 5.9 (4.3-7.9) | 13/730, 1.8 (1.0-3.0) |

Table S19. Three levels of chance for each published call rule: the error rate the rule states (or, where none is stated, implies); the rate of the reimplemented rule on white noise, which has no EEG structure; and the rate on clinical EEG without commands (primary reading). A white-noise rate above the stated rate points to the analysis design; a clinical EEG rate above the white-noise rate points to properties of EEG. P against own rate: one-sided exact binomial of the clinical EEG count against the stated (or implied) rate. P against white noise: two-sided Fisher exact test, clinical EEG against 500 white-noise recordings. P against own rate is descriptive: it uses the selected reading's full-sample count, whose P value the selection makes uninterpretable (Supplementary Methods 4); the replication tests (Table S17) are confirmatory. The du Bois 2026 implied rate of 6% is a simplified reference, not a bound on the complete rule: 5% for the t test plus at most 1% for the 70% threshold, assuming independent trials, one configuration and one time point; it does not cover the search over configurations and peak times. Guger 2018 states no error rate; its implied rate is the threshold's chance rate. Chennu 2013: its 20-block reading on the 89 development recordings that hold it (Supplementary Methods 18); Sokoliuk 2021 tests covert processing, not command-following.

| **Analysis** | **Call rule** | **Stated (implied) rate, %** | **White noise, 60** | **White noise, 500** | **Clinical EEG, k/n, % (95% CI)** | **P against own rate** | **P against white noise** |
|---|---|---|---|---|---|---|---|
| Claassen 2019 | AUC above 0.5 and permutation P < .05 | 5.0 | 3/60, 5.0 (1.0-13.9) | - | 33/481, 6.9 (4.8-9.5) | .04 | - |

| Analysis | Call rule | Stated (implied) rate, % | White noise, 60 | White noise, 500 | Clinical EEG, k/n, % (95% CI) | P against own rate | P against white noise |
|---|---|---|---|---|---|---|---|
| Goldfine 2011 | Spectral difference P <= .05 by jackknife, false discovery rate .05, contiguity | 5.0 | 0/60, 0.0 (0.0-6.0) | 0/500, 0.0 (0.0-0.7) | 164/703, 23.3 (20.2-26.6) | <.001 | <.001 |
| Cruse 2011 | Accuracy above 50% with the 99% binomial interval above 50% | 0.5 | 1/60, 1.7 (0.0-8.9) | - | 1/147, 0.7 (0.0-3.7) | .52 | - |
| Cruse 2012 | Either of 2 permutation comparisons P < .025 | 5.0 | 9/60, 15.0 (7.1-26.6) | - | 5/210, 2.4 (0.8-5.5) | .98 | - |
| Edlow 2017 | Permutation P < .05 | 5.0 | 0/60, 0.0 (0.0-6.0) | - | 3/796, 0.4 (0.1-1.1) | >.99 | - |
| Guger 2017 | Accuracy above the binomial 95% limit | 2.5 | 0/60, 0.0 (0.0-6.0) | - | 8/550, 1.5 (0.6-2.8) | .97 | - |
| du Bois 2026 | Peak accuracy above 70% or paired t test P < .05 | 5.0 (6.0, simplified reference) | 20/60, 33.3 (21.7-46.7) | 192/500, 38.4 (34.1-42.8) | 157/707, 22.2 (19.2-25.5) | <.001 | <.001 |
| Guger 2018 | Accuracy above 23% | None stated; at least 11.95 by chance (independent splits) | 9/60, 15.0 (7.1-26.6) | 76/500, 15.2 (12.2-18.7) | 117/850, 13.8 (11.5-16.3) | .06 | .47 |
| Gibson 2016 | Cluster permutation P < .05 for the target-versus-nontarget P3b | 5.0 | 1/60, 1.7 (0.0-8.9) | - | 25/683, 3.7 (2.4-5.4) | .96 | - |

| Analysis | Call rule | Stated (implied) rate, % | White noise, 60 | White noise, 500 | Clinical EEG, k/n, % (95% CI) | P against own rate | P against white noise |
|---|---|---|---|---|---|---|---|
| Sokoliuk 2021 (covert processing) | Surrogate test of phrase and sentence tracking P < .05 | 5.0 | 2/60, 3.3 (0.4-11.5) | - | 41/565, 7.3 (5.3-9.7) | .01 | - |

Table S20. The clinical EEG reading of each analysis applied unchanged to post-arrest coma (490 patients), whether the post-arrest selection picked the same reading, and the usability of the primary reading on clinical EEG with worst-case bounds (every unusable recording counted negative, or positive).

| Analysis | Post-arrest coma, clinical EEG reading, k/n, % (95% CI) | Same reading selected | Usable/attempted | Worst-case range, % |
|---|---|---|---|---|
| Claassen 2019 | 40/489, 8.2 (5.9-11.0) | Yes | 481/481 | 6.9 to 6.9 |
| Goldfine 2011 | 54/446, 12.1 (9.2-15.5) | Yes | 703/708 | 23.2 to 23.9 |
| Cruse 2011 | - | No | 147/159 | 0.6 to 8.2 |
| Cruse 2012 | - | No | 210/224 | 2.2 to 8.5 |
| Edlow 2017 | 2/484, 0.4 (0.1-1.5) | Yes | 796/803 | 0.4 to 1.2 |
| Guger 2017 | 2/418, 0.5 (0.1-1.7) | Yes | 550/704 | 1.1 to 23.0 |
| du Bois 2026 | 129/464, 27.8 (23.8-32.1) | Yes | 707/708 | 22.2 to 22.3 |
| Guger 2018 | 64/481, 13.3 (10.4-16.7) | Yes | 850/905 | 12.9 to 19.0 |
| Gibson 2016 | 28/483, 5.8 (3.9-8.3) | No | 683/703 | 3.6 to 6.4 |
| Sokoliuk 2021 (covert processing) | 32/439, 7.3 (5.0-10.1) | Yes | 565/595 | 6.9 to 11.9 |

Table S21. Range of clinical EEG false-call rates across readings consistent with each publication, three ways: every text-consistent reading; technically sound readings, which exclude readings that fit spatial filters on all trials before cross-validation, that use random cross-validation folds on a block design, or that take the best time point of an accuracy course on the data used for the call where the publication does not state that summary; and corrected readings. Flags were assigned by rule from each reading's definition after the per-reading rates had been seen. Rates in %; number of readings in parentheses. Each reading's flags are in Table S21B in Supplementary Data S2.

| Analysis | Text-consistent | Technically sound | Corrected |
|---|---|---|---|
| Claassen 2019 | 6.9 to 6.9 (1) | 6.9 to 6.9 (1) | - |
| Goldfine 2011 | 23.3 to 65.5 (9) | 23.3 to 65.5 (9) | 23.3 to 23.3 (1) |
| Cruse 2011 | 0.0 to 8.3 (16) | 0.0 to 8.3 (16) | - |
| Cruse 2012 | 2.4 to 2.9 (2) | 2.4 to 2.9 (2) | - |
| Edlow 2017 | 0.4 to 30.4 (4) | 0.4 to 0.5 (2) | - |
| Guger 2017 | 1.5 to 84.2 (4) | 1.5 to 1.5 (1) | - |
| du Bois 2026 | 22.2 to 90.0 (10) | 22.2 to 82.6 (7) | 22.2 to 84.6 (2) |
| Guger 2018 | 13.8 to 53.5 (6) | 13.8 to 20.2 (4) | - |
| Gibson 2016 | 3.7 to 9.1 (6) | 3.7 to 9.1 (6) | - |
| Sokoliuk 2021 (covert processing) | 7.3 to 53.6 (4) | 7.3 to 53.6 (4) | - |

Table S22. Illustrative share of positive calls that would be false, by assumed prevalence of covert command-following and false-call rate, assuming perfect detection and that rates measured without commands applied to patients who hear commands without following them. Not a measured quantity.

| Assumed prevalence, % | False-call rate 5.0% | False-call rate 13.8% | False-call rate 22.2% | False-call rate 23.3% | False-call rate 24.5% |
|---|---|---|---|---|---|
| 10 | 31.0 | 55.4 | 66.6 | 67.7 | 68.8 |
| 25 | 13.0 | 29.3 | 40.0 | 41.1 | 42.4 |
| 40 | 7.0 | 17.1 | 25.0 | 25.9 | 26.9 |

Table S23. Patient-level decision summary of each primary reading under its published call rule: false calls on clinical EEG at the published number of runs (k), detection of healthy volunteers' motor imagery and calls on their label-shuffled sessions (same reading, one-source sessions), and simulated detection, which was measured for size-corrected comparators (Table S14B in Supplementary Data S2), not at the published rule. ERP detection used auditory counting, a different paradigm (Table S13). Other positive-control evidence: patient data, or healthy sessions read with the primary reading itself. Edlow 2017 was the only primary reading that detected most true responders (50/51) while calling none of 51 shuffled sessions (upper 95% confidence limit, 7.0%): favorable observed performance in a limited control, with sessions assembled from trial excerpts, which does not show a false-call probability below 5%. The k = 1 rows use every patient with at least one usable window, so their denominators can differ from the first-window rates in Table S7A (motor imagery: 8/559 here, 8/550 there).

| Analysis | k | False calls at k, k/n, % (95% CI) | Healthy detection | Healthy shuffled calls | Simulated detection | Other positive-control evidence |
|---|---|---|---|---|---|---|
| Claassen 2019 | 2 | independent days 8.6; same session 7.1 | Not measured in one-source sessions | - | Not measured at the published rule | None |
| Goldfine 2011 | 1 | 164/703, 23.3 (20.2-26.6) | 51/51, 100.0 (93.0-100.0) | 14/51, 27.5 (15.9-41.7) | Not measured at the published rule | Stroke motor imagery on its recorded design: 98.4% of sessions called, 2.5% with sham labels; task performance not verified (Table S33) |

| Analysis | k | False calls at k, k/n, % (95% CI) | Healthy detection | Healthy shuffled calls | Simulated detection | Other positive-control evidence |
|---|---|---|---|---|---|---|
| Cruse 2011 | 1 | 1/147, 0.7 (0.0-3.7) | Not measured in one-source sessions | - | Not measured at the published rule | None |
| Cruse 2012 | 1 | 5/210, 2.4 (0.8-5.5) | Not measured in one-source sessions | - | Not measured at the published rule | Healthy motor imagery, original assembly, same reading: 43/47 detected, 8/49 shuffled sessions called (Table S13C in Supplementary Data S2) |
| Edlow 2017 | 1 | 3/796, 0.4 (0.1-1.1) | 50/51, 98.0 (89.6-100.0) | 0/51, 0.0 (0.0-7.0) | Not measured at the published rule | None |
| Guger 2017 | 1 | 8/559, 1.4 (0.6-2.8) | Not measured in one-source sessions | - | Not measured at the published rule | Acute stroke, instructed: 9/47 called, 3.2% of shuffled sessions; task performance not verified (Table S28) |
| du Bois 2026 | 2 | 30/95, 31.6 (22.4-41.9) | 41/51, 80.4 (66.9-90.2) | 15/51, 29.4 (17.5-43.8) | Not measured at the published rule | Stroke motor imagery on its recorded design: 86.9% of sessions called, 46.7% with sham labels; task performance not verified (Table S33) |

| Analysis | k | False calls at k, k/n, % (95% CI) | Healthy detection | Healthy shuffled calls | Simulated detection | Other positive-control evidence |
|---|---|---|---|---|---|---|
| Guger 2018 | 2 | 206/841, 24.5 (21.6-27.5) | Not measured in one-source sessions | - | Not measured at the published rule | Tactile attention in healthy volunteers: 36/38 sessions called, 9/38 with an unattended finger as the label (Table S32). In amyotrophic lateral sclerosis visual speller data, 9 of 48 patients passed (34 required; Supplementary Results 12) the entry check for a Pz response, so the rule was not run there |
| Gibson 2016 | 1 | 25/683, 3.7 (2.4-5.4) | Not measured in one-source sessions | - | Not measured at the published rule | None |
| Sokoliuk 2021 (covert processing) | 1 | 41/565, 7.3 (5.3-9.7) | Not measured in one-source sessions | - | Not measured at the published rule | None |

Table S24. Independent implementation. The du Bois 2026 and Guger 2018 analyses were written a second time from the publication texts alone, without access to this study's code, and run on the same 200 clinical EEG development windows with the same sham schedules as this study's primary readings (Guger 2018 on the windows where both returned a result). Bars set before the run: the independent rate inside the range of readings consistent with the publication, and kappa of at least 0.6. False calls on recordings without commands are chance events specific to an implementation, so agreement on which windows are called can be low even when rates are similar.

| Analysis | Windows | This study, k/n, % (95% CI) | Independent, k/n, % (95% CI) | Kappa | Agreement, % | Range of consistent readings, % | Independent rate inside range |
|---|---|---|---|---|---|---|---|
| du Bois 2026 | 200 | 53/200, 26.5 (20.5-33.2) | 192/200, 96.0 (92.3-98.3) | 0.02 | 29.5 | 22.2 to 90.0 | No |
| Guger 2018 | 178 | 15/178, 8.4 (4.8-13.5) | 31/178, 17.4 (12.2-23.8) | 0.26 | 83.1 | 13.8 to 53.5 | Yes |

Table S26. Post-arrest coma: patients called positive without commands, by outcome (poor: Cerebral Performance Category 3 to 5, 305 patients; good: 1 or 2, 185). Primary reading, first overlay of the window nearest 24 hours after return of circulation. How clinicians would act on such a call is not known; the share describes exposure, not harm.

| Analysis | Poor outcome, k/n, % (95% CI) | Good outcome, k/n, % (95% CI) |
|---|---|---|
| Claassen 2019 | 29/304, 9.5 (6.5-13.4) | 11/185, 5.9 (3.0-10.4) |
| Goldfine 2011 | 40/273, 14.7 (10.7-19.4) | 14/173, 8.1 (4.5-13.2) |
| du Bois 2026 | 73/287, 25.4 (20.5-30.9) | 56/177, 31.6 (24.9-39.0) |
| Edlow 2017 | 2/300, 0.7 (0.1-2.4) | 0/184, 0.0 (0.0-2.0) |
| Guger 2017 | 2/247, 0.8 (0.1-2.9) | 0/171, 0.0 (0.0-2.1) |
| Gibson 2016 | 8/301, 2.7 (1.2-5.2) | 14/182, 7.7 (4.3-12.6) |
| Guger 2018 | 44/300, 14.7 (10.9-19.2) | 20/181, 11.0 (6.9-16.5) |
| Sokoliuk 2021 (covert processing) | 24/271, 8.9 (5.8-12.9) | 8/168, 4.8 (2.1-9.2) |
| Any of Goldfine 2011, du Bois 2026 and Guger 2018 (patients assessed by all 3) | 127/270, 47.0 (41.0-53.2) | 79/171, 46.2 (38.6-54.0) |

Table S27. Published positive rates with delivered commands against the rate of the same call rule on EEG recorded without study commands, at the published number of assessments. du Bois 2026: advancement after Phase I (either of 2 sessions); no-command rate of the two-session criterion on clinical EEG (primary) and on prolonged-DoC EEG (secondary); one-sided Fisher exact test. Claassen 2019: EEG brain activation; the 2-recording rate of the Claassen 2019 rule assuming independent recordings, in post-arrest coma (primary) and on clinical EEG (secondary); one-sided exact binomial test. Because that rate is estimated, the primary Claassen P is also .001 to .34 across the 95% CI of the no-command rate, and .01 against the 1-recording rate (identical recordings). Holm adjustment over the 4 primary comparisons. A rate above the no-command rate is signal beyond the rule's chance behavior; a rate within it cannot be told apart from chance behavior by these data. Neither says which published patients were true or false positives. UWS indicates unresponsive wakefulness syndrome; MCS, minimally conscious state; LIS, locked-in syndrome.

| Cohort | Published, k/n, % (95% CI) | No-command rate (primary) | P | Holm P | No-command rate (secondary) | P (secondary) |
|---|---|---|---|---|---|---|
| du Bois UWS | 8/14, 57.1 (28.9-82.3) | 30/95, 31.6 (22.4-41.9) | .06 | .06 | 18/38, 47.4 (31.0-64.2) | .38 |
| du Bois MCS | 13/17, 76.5 (50.1-93.2) | 30/95, 31.6 (22.4-41.9) | <.001 | .002 | 18/38, 47.4 (31.0-64.2) | .04 |
| du Bois LIS | 10/11, 90.9 (58.7-99.8) | 30/95, 31.6 (22.4-41.9) | <.001 | <.001 | 18/38, 47.4 (31.0-64.2) | .01 |
| Claassen 2019 | 16/104, 15.4 (9.1-23.8) | 9.3 | .03 | .06 | 8.6 | .02 |

Table S28. Positive control in patients with acute stroke instructed to imagine hand movements (open data, 47 patients 1 to 30 days after stroke; 3 duplicate recordings excluded): the Guger 2017, 40-trial variant (published run: 60 trials), on each patient's own 40 left and right hand imagery trials, common spatial patterns with linear discriminant analysis, 10 x 10-fold cross-validation; a call is mean accuracy above 66.2%, the binomial bound at 40 trials by the formula that gives 63.19% at 60. False calls: 20 random permutations of each patient's labels; rate with a patient-cluster bootstrap CI that relabels resampled patients. An occipital-only run checked that the hand-specific video did not drive accuracy (median 49.5% vs 52.0% sensorimotor). Mean accuracy was 54.6% on sensorimotor channels and 52.2% on all channels; the dataset's authors reported 55.57% with the same method.[38] The lower-limb motor imagery set of 27 patients was not used: in its first 3 patients, idle trials desynchronized against fixation like imagery trials (patient 1: accuracy 90.0%, sensorimotor 68.8%, imagery vs idle P = .08, idle vs fixation P = .02; patient 2: accuracy 63.0%, sensorimotor 58.0%, imagery vs idle P = .07, idle vs fixation P = .04; patient 3: accuracy 71.2%, sensorimotor 67.5%, imagery vs idle P = .10, idle vs fixation P = .03). Task performance was not verified, so calls with true labels are the share called positive after the instruction, not sensitivity in verified responders or in disorders of consciousness.

| Channels | Called positive, true labels, k/n, % (95% CI) | False calls, shuffled labels, % (95% CI) | Patients with at least 1 false call in 20 shuffles | Called positive minus false calls, points (95% CI) |
|---|---|---|---|---|
| Sensorimotor channels (primary) | 9/47, 19.1 (9.1-33.3) | 3.2 (2.1-4.4) | 21/47, 44.7 (30.2-59.9) | 16.0 (5.1-27.3) |

| Channels | Called positive, true labels, k/n, % (95% CI) | False calls, shuffled labels, % (95% CI) | Patients with at least 1 false call in 20 shuffles | Called positive minus false calls, points (95% CI) |
|---|---|---|---|---|
| All channels | 7/47, 14.9 (6.2-28.3) | 2.4 (1.6-3.4) | 19/47, 40.4 (26.4-55.7) | 12.4 (3.5-23.2) |

Table S30. The second du Bois 2026 implementation, our departures from it and the publication's permutation evaluation, on the 200 clinical EEG development windows of Table S24 (first overlay, same schedules). The second implementation (Supplementary Methods 16) follows more of the publication's stated choices (3 filter pairs per band, mutual information with 3 quantization levels, 40-ms step, accuracy at each time point, raw-curve peak within 150 ms of the smoothed peak), uses every recorded channel and draws random stratified folds, a fold rule the publication does not state. A: each departure of our primary reading applied alone to the second implementation, and cumulatively; gap share: the share of the difference in rates between it and our primary reading closed by that change alone; kappa: agreement on which windows are called, against our primary reading. Block-wise folds alone: exact McNemar 109 vs 2, P < .001. After every listed change, the training scheme (one classifier per time point vs one per fold on pooled training times) and the spatial-filter covariance, which were not varied, still differ. B: folds x peak rule x configuration search (fixed configuration, chosen before any window was run: the motor channel set, the bands from mu to low gamma, a 2-s window and 7 features). C: the publication's 100-permutation evaluation (P < .05), alone and with the advancement criterion; synthetic recordings as in Supplementary Methods 10, with slow amplitude drift added.

A. Single and cumulative changes

| Reading | Positive calls, k/n, % (95% CI) | Gap share | Kappa with our primary reading |
|---|---|---|---|
| Second implementation as written (random folds, refined peak, 84-configuration search) | 192/200, 96.0 (92.3-98.3) | - | 0.02 |
| One change: block-wise folds (leave one block out) | 85/200, 42.5 (35.6-49.7) | 0.77 | -0.08 |
| One change: 17 standard sites (all but Fp1 and Fp2) | 190/200, 95.0 (91.0-97.6) | 0.01 | 0.01 |
| One change: our peak rule (smoothed maximum, no refinement; paired t at the smoothed peak) | 164/200, 82.0 (76.0-87.1) | 0.20 | -0.01 |
| One change: 100-ms step | 184/200, 92.0 (87.3-95.4) | 0.06 | -0.03 |
| One change: our band edges | 191/200, 95.5 (91.6-97.9) | 0.01 | 0.01 |
| One change: 2 filter pairs per band | 188/200, 94.0 (89.8-96.9) | 0.03 | 0.00 |
| One change: Gaussian mutual information | 189/200, 94.5 (90.4-97.2) | 0.02 | 0.01 |
| One change: Ledoit-Wolf shrinkage | 189/200, 94.5 (90.4-97.2) | 0.02 | 0.01 |
| Cumulative to L1: + block-wise folds (leave one block out) | 85/200, 42.5 (35.6-49.7) | - | -0.08 |
| Cumulative to L2: + 17 standard sites (all but Fp1 and Fp2) | 83/200, 41.5 (34.6-48.7) | - | -0.02 |
| Cumulative to L3: + our peak rule (smoothed maximum, no refinement; paired t at the smoothed peak) | 86/200, 43.0 (36.0-50.2) | - | 0.03 |
| Cumulative to L4: + 100-ms step | 73/200, 36.5 (29.8-43.6) | - | 0.08 |
| Cumulative to L5: + our band edges | 70/200, 35.0 (28.4-42.0) | - | 0.06 |
| Cumulative to L6: + 2 filter pairs per band | 68/200, 34.0 (27.5-41.0) | - | 0.05 |
| Cumulative to L7: + Gaussian mutual information | 68/200, 34.0 (27.5-41.0) | - | 0.07 |
| Cumulative to L8: + Ledoit-Wolf shrinkage | 81/200, 40.5 (33.6-47.7) | - | 0.10 |

| Reading | Positive calls, k/n, % (95% CI) | Gap share | Kappa with our primary reading |
|---|---|---|---|
| Our implementation, block-wise folds (primary reading) | 53/200, 26.5 (20.5-33.2) | 1.00 | 1.00 |
| Our implementation, random folds | 174/200, 87.0 (81.5-91.3) | - | 0.03 |

B. Factorial

| Folds | Peak rule | Configuration | Positive calls, k/n, % (95% CI) |
|---|---|---|---|
| Block-wise | Ours | Fixed in advance | 40/200, 20.0 (14.7-26.2) |
| Block-wise | Ours | 84-configuration search | 91/200, 45.5 (38.5-52.7) |
| Block-wise | Refined (published) | Fixed in advance | 28/200, 14.0 (9.5-19.6) |
| Block-wise | Refined (published) | 84-configuration search | 85/200, 42.5 (35.6-49.7) |
| Random | Ours | Fixed in advance | 70/200, 35.0 (28.4-42.0) |
| Random | Ours | 84-configuration search | 164/200, 82.0 (76.0-87.1) |
| Random | Refined (published) | Fixed in advance | 85/200, 42.5 (35.6-49.7) |
| Random | Refined (published) | 84-configuration search | 192/200, 96.0 (92.3-98.3) |

C. Permutation evaluation

| Permutation null | Win dow s | Random folds | Block-wise folds | Advancement and permutation, random folds | Advancement and permutation, block-wise folds |
|---|---|---|---|---|---|
| Configuration fixed after selection, P at the selected peak (literal) | 200 | 200/200, 100.0 (98.2-100.0) | 112/200, 56.0 (48.8-63.0) | 192/200, 96.0 (92.3-98.3) | 61/200, 30.5 (24.2-37.4) |
| Configuration search repeated in each permutation | 200 | 156/200, 78.0 (71.6-83.5) | 12/200, 6.0 (3.1-10.2) | 156/200, 78.0 (71.6-83.5) | 12/200, 6.0 (3.1-10.2) |
| Search repeated, synthetic 1/f EEG without drift | 30 | 2/30, 6.7 (0.8-22.1) | 0/30, 0.0 (0.0-11.6) | 2/30, 6.7 (0.8-22.1) | 0/30, 0.0 (0.0-11.6) |
| Search repeated, synthetic 1/f EEG with strong drift | 30 | 29/30, 96.7 (82.8-99.9) | 8/30, 26.7 (12.3-45.9) | 29/30, 96.7 (82.8-99.9) | 8/30, 26.7 (12.3-45.9) |

Table S31. Goldfine 2011 with a variance that respects dependence within commands. The primary reading takes its jackknife over single tapered estimates (5 tapers x 3-s epochs), treating them as independent; the command-level jackknife deletes all estimates of one command at a time. Variance inflation: the command-level jackknife variance of the standardized spectral difference, whose expected value for independent estimates is 1, median over channels of the band mean. The resolution element is 2 Hz (time-bandwidth 3 on 3-s epochs), the width of the >2 Hz contiguity criterion. Clinical EEG: the 342 development recordings held locally, first overlay; the primary reading reproduced the stored call in 342 of 342. Median variance inflation was 5.19 in windows the primary reading called and 3.63 in the others. Synthetic: 100 command-free 1/f recordings per drift level (Supplementary Methods 10, slow amplitude drift added).

| **Reading** | **Clinical EEG, k/n, % (95% CI)** | **Synthetic, no drift** | **Synthetic, drift 0.5** | **Synthetic, drift 1.0** |
|---|---|---|---|---|
| Sample-level jackknife (our primary reading) | 79/342, 23.1 (18.7-27.9) | 0/100, 0.0 (0.0-3.6) | 2/100, 2.0 (0.2-7.0) | 29/100, 29.0 (20.4-38.9) |
| Sample-level jackknife, one frequency bin per resolution element | 59/342, 17.3 (13.4-21.7) | 0/100, 0.0 (0.0-3.6) | 2/100, 2.0 (0.2-7.0) | 21/100, 21.0 (13.5-30.3) |
| Command-level jackknife, normal reference | 2/342, 0.6 (0.1-2.1) | 0/100, 0.0 (0.0-3.6) | 0/100, 0.0 (0.0-3.6) | 0/100, 0.0 (0.0-3.6) |
| Command-level jackknife, t reference | 2/342, 0.6 (0.1-2.1) | 0/100, 0.0 (0.0-3.6) | 0/100, 0.0 (0.0-3.6) | 0/100, 0.0 (0.0-3.6) |
| Command-level jackknife, t reference, one bin per resolution element | 2/342, 0.6 (0.1-2.1) | 0/100, 0.0 (0.0-3.6) | 0/100, 0.0 (0.0-3.6) | 0/100, 0.0 (0.0-3.6) |

| Reading | Clinical EEG, k/n, % (95% CI) | Synthetic, no drift | Synthetic, drift 0.5 | Synthetic, drift 1.0 |
|---|---|---|---|---|
| Variance inflation, median (IQR); share | 3.92 (2.88-6.18); 93.9% at least 2 | 1.05 (1.04-1.06); 0.0% at least 2 | 1.54 (1.42-1.81); 11.0% at least 2 | 3.68 (2.83-5.08); 98.0% at least 2 |

Table S32. Tactile P300 positive control with genuine attention (open Braille brain-computer interface data, 19 blind or sighted adults x 2 sessions; one participant was used only to build the loader).[36] Each fingertip rests on 1 of 8 Braille cells; each selection cycle raises every cell 10 times in random order, and the participant counts the target cell's activations. Guger 2018 ran on its own 8-stimulus layout, unchanged except that a stimulus is a fingertip; run A is cycles 1 to 6 and run B cycles 7 to 12, and a session is called if either run is (the per-patient rule of the replication). True: the attended finger as the label. Sham: the same recordings labeled with an unattended finger, by a per-session derangement of the 8 fingers. Randomization test: the run's targets permuted across cycles, 200 draws. Sham calls of the primary reading by hand: 1 of 7 sessions in which the sham label mostly stayed on the attended hand and 8 of 31 in which it moved to the other hand. The published 3 of 3 bar (upper confidence limit reaching 100%) is not met by 36 of 38 (upper limit 99.4%). Exact CIs over sessions, not adjusted for the 2 sessions of each participant.

| Reading | True label, sessions called | Sham label, sessions called | Participants called, true | Participants called, sham | McNemar, true only vs sham only |
|---|---|---|---|---|---|
| Published rule, our primary reading | 36/38, 94.7 (82.3-99.4) | 9/38, 23.7 (11.4-40.2) | 19/19 | 8/19 | 27 vs 0, P < .001 |
| Published rule, default reading | 35/38, 92.1 (78.6-98.3) | 13/38, 34.2 (19.6-51.4) | 19/19 | 10/19 | 22 vs 0, P < .001 |
| Independent implementation | 37/38, 97.4 (86.2-99.9) | 13/38, 34.2 (19.6-51.4) | 19/19 | 9/19 | 24 vs 0, P < .001 |
| Randomization test (targets permuted across cycles) | 33/38, 86.8 (71.9-95.6) | 2/38, 5.3 (0.6-17.7) | 18/19 | 2/19 | 31 vs 0, P < .001 |

Table S33. Stroke motor imagery on its recorded design (open data, 24 patients 3 to 14 months after stroke, sessions at 3 time points, 4 runs each; one patient was used only to build the loader).[37] Each trial is 7 s of rest with a fixation cross, 1 s of preparation and 6 s of imagery of the affected hand or of a robotic sixth finger, the imagery type in random order. The analyses ran on each session's own trials and timing: Goldfine 2011, imagery against the preceding rest; du Bois 2026, affected-hand against sixth-finger imagery, the session's runs joined with 0.25-s cross-fades (random folds are legitimate here because the imagery type was randomized). Sham (Goldfine 2011): rest periods only, each split into halves labeled rest and task, half of each run's rest periods in each order. Sham (du Bois 2026): the trial labels redrawn at random. Exchanged (Goldfine 2011): exactly half of each run's trial labels swapped, a comparator that keeps imagery in both classes and is sensitive to slow drift. 20 draws of each per session. Rates with patient-cluster bootstrap 95% CIs. Byte-identical sessions, found by file hash, excluded when the rows were merged, before any merged result was read: 3 (S21_pre, identical to S22_pre (other patient); S22_pre, identical to S21_pre (other patient); S2_follow, identical to S2_post (same patient)). No Goldfine 2011 call with true labels rested on posterior sites only. Task performance was not verified.

| Reading | Sessions / patients | Called with true labels, % (95% CI) | Patients with at least 1 call, % | Sham, % (95% CI) | Exchanged labels, % (95% CI) |
|---|---|---|---|---|---|
| Goldfine 2011, sample-level jackknife (our primary reading) | 61 / 23 | 98.4 (95.0-100.0) | 100.0 | 2.5 (1.2-4.4) | 12.0 (7.9-16.8) |

| Reading | Sessions / patients | Called with true labels, % (95% CI) | Patients with at least 1 call, % | Sham, % (95% CI) | Exchanged labels, % (95% CI) |
|---|---|---|---|---|---|
| Goldfine 2011, sample-level, one bin per resolution element | 61 / 23 | 95.1 (85.0-100.0) | 95.7 | 1.0 (0.4-1.7) | 7.8 (4.6-11.7) |
| Goldfine 2011, command-level jackknife, normal reference | 61 / 23 | 93.4 (82.5-100.0) | 95.7 | 0.0 (0.0-0.0) | 0.6 (0.1-1.2) |
| Goldfine 2011, command-level jackknife, t reference | 61 / 23 | 93.4 (82.5-100.0) | 95.7 | 0.0 (0.0-0.0) | 0.3 (0.0-0.8) |
| Goldfine 2011, command-level, t reference, one bin per resolution element | 61 / 23 | 93.4 (82.5-100.0) | 95.7 | 0.0 (0.0-0.0) | 0.2 (0.0-0.4) |
| du Bois 2026, second implementation | 61 / 23 | 93.4 (86.7-98.4) | 95.7 | 68.0 (65.4-70.4) | - |
| du Bois 2026, our implementation (random folds) | 61 / 23 | 86.9 (73.8-96.8) | 91.3 | 46.7 (43.2-50.2) | - |

Table S34. Detection at thresholds targeting a patient-level false-call rate, set within each design. Each analysis's evidence statistic was thresholded on recordings without commands from the same design, targeting a 5% chance that a patient assessed k times is called positive by chance: per-assessment level $1 - 0.95^{(1/k)}$, the (1 - level) quantile of the null statistic, a call strictly above it. Thresholds were cross-fitted by patient (two halves by a hash of the patient identifier; each half's null judged by the other half's threshold, and each task unit by the threshold of the half its own participant or patient does not fall in under the same hash) and applied to genuine responses; a threshold is never carried across designs. Cells: single-assessment detection (achieved held-out false calls pooled over the two halves; 95% CI from a bootstrap of participants or patients, 2,000 resamples with both thresholds re-estimated in each, so it reflects runs and sessions clustered within participants and the estimation of each threshold), %; target per-assessment levels 5.0%, 2.5%, 1.0% and 0.5% for k = 1, 2, 5 and 10. ↑, achieved rate above its target, descriptive; †, target finer than the null can resolve (a threshold set on the n null units of one half cannot reach a held-out rate below 1/(n + 1)); detection in marked cells is optimistic. Achieved rates were not tested against their targets. The per-assessment level assumes independent assessments; with positive association between a patient's assessments, the patient-level rate is lower. Published rule: detection (false-call rate of the same rule on the same null, where measured). Randomization: detection by the randomization-based version at p at or below the per-assessment level, k = 1 / 2 / 5 / 10, on randomized-order sessions (volunteers) or by the across-cycle permutation test (tactile runs). Tactile runs: only 38 null runs per half, so the 5% quantile at k = 1 is coarse. k: 2 (acute intensive care, a median of 2 recordings[3]), 5 (prolonged disorders of consciousness[41]) and 10 (daily sessions[32^]).

| Analysis | Genuine task | Null | Task units / null units | k = 1 | k = 2 | k = 5 | k = 10 | Published rule | Randomization, k = 1 / 2 / 5 / 10 |
|---|---|---|---|---|---|---|---|---|---|
| Goldfine 2011 | Healthy volunteers, motor imagery (original assembly) | Clinical EEG without commands | 51 / 703 | 100.0 (4.8; 4.1-6.5) | 100.0 (2.6; 2.0-3.8)↑ | 100.0 (1.0; 0.7-2.0) | 96.1 (0.4; 0.3-1.3) | 100.0 | 96 / 96 / 94 / 94 |
| du Bois 2026 (our implementation, paired t) | Healthy volunteers, motor imagery (original assembly) | Clinical EEG without commands | 51 / 707 | 21.6 (4.4; 3.7-6.5) | 5.9 (3.0; 2.0-4.0)↑ | 2.0 (1.0; 0.7-2.4) | 2.0 (0.4; 0.3-1.3) | 96.1 | 86 / 84 / 82 / 75 |
| Guger 2018 | Healthy volunteers, auditory counting | Clinical EEG without stimuli | 13 / 850 | 61.5 (6.4; 4.8-8.0)↑ | 30.8 (2.5; 2.0-5.2) | 7.7 (1.8; 0.9-2.9)↑ | 0.0 (1.5; 0.4-2.5)↑ | 69.2 | 31 / 23 / 23 / 23 |
| Guger 2018 | Tactile attention, runs (Table S32) | Sham-labeled runs | 76 / 76 | 61.8 (10.5; 2.6-18.4)↑ | 47.4 (2.6; 2.6-15.8)† | 30.3 (2.6; 2.6-17.1)† | 30.3 (2.6; 2.6-15.8)† | 81.6 (13.2) | 72 / 70 / 62 / 51 |

| Analysis | Genuine task | Null | Task units / null units | k = 1 | k = 2 | k = 5 | k = 10 | Published rule | Randomization, k = 1 / 2 / 5 / 10 |
|---|---|---|---|---|---|---|---|---|---|
| Goldfine 2011, sample-level | Stroke motor imagery (Table S33) | Rest-only sham | 61 / 1220 | 96.7 (4.9; 4.2-6.8) | 96.7 (2.6; 2.0-4.0)↑ | 95.1 (1.1; 0.7-2.0)↑ | 95.1 (0.5; 0.3-1.4) | 98.4 (2.5) | - |
| Goldfine 2011, command-level | Stroke motor imagery (Table S33) | Rest-only sham | 61 / 1220 | 98.4 (5.6; 4.5-7.1)↑ | 96.7 (2.3; 1.8-3.5) | 95.1 (1.5; 0.7-2.3)↑ | 95.1 (0.4; 0.2-1.7) | 93.4 (0.0) | - |
| du Bois 2026 (our implementation, paired t) | Stroke motor imagery (Table S33) | Labels redrawn | 61 / 1220 | 39.3 (5.7; 4.3-6.6)↑ | 27.9 (2.6; 1.8-3.1)↑ | 16.4 (1.1; 0.6-1.8)↑ | 9.8 (0.7; 0.3-1.3)↑ | 86.9 (46.7) | - |

Table S35. The command-level Goldfine 2011 variance, fixed before testing, in windows it was not developed on (Supplementary Methods 17). The frozen record (primary reading: command-level jackknife, normal reference; test: one-sided exact binomial of the count against 5%, Holm over the replication and post-arrest sets) was committed before any of these windows was run. Each window is the patient's first published Goldfine 2011 overlay; the published (sample-level) reading was rerun on the same features and reproduced the stored call in every usable window. Left out: replication, 8 windows without usable trials; post-arrest, 49 without usable trials and 1 without a fitting overlay, as in the primary analysis. McNemar: windows called by the published reading only vs the command-level reading only. Strata are descriptive: post-arrest, the window's ACNS continuity grade; development, the window's MORGOTH background (awake, sleep, mixed, or ICU-typical: burst suppression, periodic or diffuse slowing).

| **Set** | **Published jackknife** | **Command-level, normal** | **Command-level, t** | **Command-level, t, one bin per resolution element** | **P for a rate below 5%** | **McNemar** |
|---|---|---|---|---|---|---|
| Development, held locally | 79/342, 23.1 (18.7-27.9) | 2/342, 0.6 (0.1-2.1) | 2/342, 0.6 (0.1-2.1) | 2/342, 0.6 (0.1-2.1) | - | - |
| Replication | 165/722, 22.9 (19.8-26.1) | 6/722, 0.8 (0.3-1.8) | 4/722, 0.6 (0.2-1.4) | 3/722, 0.4 (0.1-1.2) | P < .001 (Holm < .001) | 159 vs 0 |
| Post-arrest coma | 57/494, 11.5 (8.9-14.7) | 12/494, 2.4 (1.3-4.2) | 9/494, 1.8 (0.8-3.4) | 8/494, 1.6 (0.7-3.2) | P = .003 (Holm = .003) | 45 vs 0 |

| Stratum | Published jackknife | Command-level, normal |
|---|---|---|
| Post-arrest, burst suppression | 24/174, 13.8 (9.0-19.8) | 6/174, 3.4 (1.3-7.4) |
| Post-arrest, continuous | 10/135, 7.4 (3.6-13.2) | 1/135, 0.7 (0.0-4.1) |
| Post-arrest, discontinuous | 18/110, 16.4 (10.0-24.6) | 5/110, 4.5 (1.5-10.3) |
| Post-arrest, suppressed | 5/75, 6.7 (2.2-14.9) | 0/75, 0.0 (0.0-4.8) |
| Development, awake | 23/112, 20.5 (13.5-29.2) | 0/112, 0.0 (0.0-3.2) |
| Development, ICU-typical | 2/17, 11.8 (1.5-36.4) | 0/17, 0.0 (0.0-19.5) |
| Development, mixed | 18/63, 28.6 (17.9-41.3) | 2/63, 3.2 (0.4-11.0) |
| Development, sleep | 36/150, 24.0 (17.4-31.6) | 0/150, 0.0 (0.0-2.4) |

Table S36. Cluster-aware inference for the genuine-task controls. Percentile 95% CIs from 2000 resamples of whole patients (stroke) or participants (tactile). Stroke: true-label detection by reading, and the paired change in detection from the published to the command-level Goldfine 2011 jackknife on the same 61 sessions: 4.9 points (0.0-12.3); 3 sessions called by the published reading only and 0 by the command-level reading only (exact McNemar P = .25). Tactile: session calls (either of runs A and B) with the attended finger (true) and an unattended finger (sham) as the label, their difference, the per-run sham rate, participants with at least one call, and an exact sign test over participants of true minus sham sessions called.

| **Stroke reading** | **Sessions** | **Patients** | **Called with true labels, % (95% CI)** |
|---|---|---|---|
| Goldfine 2011, published jackknife | 61 | 23 | 98.4 (95.1-100.0) |
| Goldfine 2011, command-level jackknife | 61 | 23 | 93.4 (83.3-100.0) |
| Goldfine 2011, command-level, t reference | 61 | 23 | 93.4 (83.3-100.0) |
| du Bois 2026, second implementation | 61 | 23 | 93.4 (86.8-98.4) |
| du Bois 2026, our implementation (random folds) | 61 | 23 | 86.9 (74.6-96.8) |

| Tactile reading | True, % (95% CI) | Sham, % (95% CI) | Difference, points (95% CI) | Sham per run, % (95% CI) | Participants called, true / sham | Sign test |
|---|---|---|---|---|---|---|
| Published threshold, primary reading | 94.7 (86.8-100.0) | 23.7 (10.5-36.8) | 71.1 (55.3-84.2) | 13.2 (6.6-21.1) | 19 / 8 | 17 vs 0 (2 ties), P < .001 |
| Published threshold, default reading | 92.1 (84.2-100.0) | 34.2 (18.4-50.0) | 57.9 (39.5-73.7) | 18.4 (10.5-27.6) | 19 / 10 | 15 vs 0 (4 ties), P < .001 |
| Independent implementation | 97.4 (92.1-100.0) | 34.2 (18.4-52.6) | 63.2 (44.7-78.9) | 22.4 (11.8-35.5) | 19 / 9 | 15 vs 0 (4 ties), P < .001 |
| Randomization test | 86.8 (73.7-97.4) | 5.3 (0.0-13.2) | 81.6 (68.4-94.7) | 2.6 (0.0-6.6) | 18 / 2 | 18 vs 0 (1 tie), P < .001 |

Table S37. Time-matched specificity of Goldfine 2011: left- against right-hand imagery at the same time after the cue (one 3-s epoch, 0.5-3.5 s after imagery onset), without rest, in 47 patients with acute stroke (Table S28 data; sub-05, sub-15, sub-49 excluded as duplicates). Four pseudo-runs of 10 consecutive trials; the Goldfine 2011 decision with the published (sample-level) or the command-level jackknife on the same features. Null: exactly half of the 20 consecutive left / right pairs with their labels swapped, 20 draws per patient; rate with a patient-cluster bootstrap CI. Contralateral: a call with any FDR-surviving bin at C3 or C4 in 8-24 Hz with lower power for the contralateral hand's imagery; posterior only: every surviving bin at O1, O2, P7, P8, T5 or T6. Reading fixed beforehand: sensorimotor-specific if at least half of the true-label calls are contralateral and at most a quarter posterior only; null calls were as often contralateral and never posterior only, so the rule does not separate true from null calls here. P against null: one-sided binomial of the patients called against the mean null rate. The cues are hand-specific videos, so the contrast is matched in time, not in stimulus. Task performance was not verified.

| **Reading** | **Patients called, k/n, % (95% CI)** | **Null calls, % (95% CI)** | **P against null** | **True calls contralateral** | **Null calls contralateral** | **True calls posterior only** | **Sensorimotor-specific (fixed rule)** |
|---|---|---|---|---|---|---|---|
| Published jackknife | 5/47, 10.6 (3.5-23.1) | 7.7 (4.3-11.5) | P = .29 | 5/5 | 34/72 | 0/5 | Yes |
| Command-level jackknife | 2/47, 4.3 (0.5-14.5) | 0.5 (0.0-1.2) | P = .03 | 1/2 | 1/5 | 0/2 | Yes |

Table S38. Patient-level calibration measured directly on repeated null runs. Clinical EEG without commands, analysis split, published schedules: every overlay (run) of a patient's window; patients with a usable statistic on each of the first k runs (others excluded, in parentheses). Direct threshold: the 95th percentile ('higher') of each half's patient maxima over k runs; held-out patient-level false calls: patients of the other half whose maximum exceeds it, pooled over halves (exact CI). Independence-based threshold: the per-assessment quantile (level 1 - 0.95^(1/k)) of the first-run statistics of the same patients, so it can differ from Table S34, which uses every patient's first run; at k = 5 and 10 the two halves' independence-based thresholds differed widely. Detection: single-assessment detection of the healthy volunteers (Guger 2018: auditory counting) at each threshold. Runs are windows of one recording.

| Analysis | k | Patients (excluded) | Direct: held-out patient-level false calls | Direct: detection, % | Independence: held-out patient-level false calls | Independence: detection, % |
|---|---|---|---|---|---|---|
| Guger 2018 | 2 | 806 (77) | 41/806, 5.1 (3.7-6.8) | 38.5 | 39/806, 4.8 (3.5-6.6) | 30.8 |
| Guger 2018 | 5 | 661 (222) | 32/661, 4.8 (3.3-6.8) | 7.7 | 41/661, 6.2 (4.5-8.3) | 7.7 |
| Guger 2018 | 10 | 266 (617) | 17/266, 6.4 (3.8-10.0) | 7.7 | 17/266, 6.4 (3.8-10.0) | 0.0 |
| du Bois 2026 paired t test | 2 | 95 (612) | 7/95, 7.4 (3.0-14.6) | 5.9 | 3/95, 3.2 (0.7-9.0) | 3.9 |

Table S39. Analyses run later at full length, and Claassen 2019 on the six sites of the prolonged-DoC recordings. A: schedules longer than the 30-minute replication and post-arrest windows (Chennu 2013 2,550 s, Cruse 2011 2,186 s and Cruse 2012 1,891 s, with lead and tail), run on one longer clean window per patient where the recording holds one (Supplementary Methods 18), and Sokoliuk 2021 on the replication set's own windows. Chennu 2013 at its stated 20 blocks on the clinical development recordings that hold them, its reading chosen by the rule of Supplementary Methods 4; every reading was fixed before the new windows were analyzed. Post-arrest: the patients whose segments were read, in patient-identifier order (C). First overlay; P: one-sided exact binomial against 5%, descriptive and outside the Holm families.

| Analysis | Recordings | Reading | Windows run | False calls, k/n, % (95% CI) | P against 5% | Note |
|---|---|---|---|---|---|---|
| Chennu 2013 | Clinical EEG, development set | Chosen here, fewer calls of 2 eligible | 89 of 364 recordings of at least 2,550 s | 0/89, 0.0 (0.0-4.1) | >.99 | Other eligible reading 2/89, 2.2 (0.3-7.9); cross-fitted held out 2/89, 2.2 (0.3-7.9) |
| Chennu 2013 | Clinical EEG, replication set | Chosen on development | 231 long windows | 0/231, 0.0 (0.0-1.6) | >.99 | - |
| Cruse 2011 | Clinical EEG, replication set | Development primary | 398 long windows | 7/365, 1.9 (0.8-3.9) | >.99 | 33 without a usable result |
| Cruse 2012 | Clinical EEG, replication set | Development primary | 540 long windows | 17/495, 3.4 (2.0-5.4) | .96 | 45 without a usable result |
| Sokoliuk 2021 (covert processing) | Clinical EEG, replication set | Development primary | 730 30-minute windows | 58/693, 8.4 (6.4-10.7) | <.001 | 37 without a usable result |
| Chennu 2013 | Coma after cardiac arrest | Chosen on development | 198 long windows | 0/198, 0.0 (0.0-1.8) | >.99 | - |
| Cruse 2011 | Coma after cardiac arrest | Development primary | 216 long windows | 5/205, 2.4 (0.8-5.6) | .98 | 11 without a usable result |

| Analysis | Recordings | Reading | Windows run | False calls, k/n, % (95% CI) | P against 5% | Note |
|---|---|---|---|---|---|---|
| Cruse 2012 | Coma after cardiac arrest | Development primary | 231 long windows | 6/223, 2.7 (1.0-5.8) | .97 | 8 without a usable result |

B: Claassen 2019 on the six sites F3, F4, C3, C4, O1 and O2, the reduced montage of every whole-head analysis on the prolonged-DoC recordings: the released pipeline with its current source density rebuilt over the six sites (24 features instead of 76) and its classifier, cross-validation, permutation test and call unchanged. On clinical EEG it ran on the same windows, schedules and permutation seeds as the 19-site results, so the pairs differ only in montage. Pairs called by both / by 19 sites only / by 6 sites only; exact McNemar test.

| Recordings | Test | 19 sites, k/n, % (95% CI) | 6 sites, k/n, % (95% CI) | Called: both / 19 only / 6 only | McNemar P |
|---|---|---|---|---|---|
| Clinical EEG, development set (481 paired recordings) | Published test | 33/481, 6.9 (4.8-9.5) | 40/481, 8.3 (6.0-11.2) | 9 / 24 / 31 | .42 |
| Clinical EEG, development set (481 paired recordings) | Randomization-based test | 21/481, 4.4 (2.7-6.6) | 26/481, 5.4 (3.6-7.8) | 5 / 16 / 21 | .51 |
| Prolonged DoC | Published test | - | 1/38, 2.6 (0.1-13.8) | - | - |
| Prolonged DoC | Randomization-based test | - | 0/38, 0.0 (0.0-9.3) | - | - |

C: Post-arrest patients whose segments were read for the long windows, in patient-identifier order (0284 to 0621), by hospital; no outcome was read. Long windows: 198 at 2,550 s, 18 at 2,186 s and 15 at 1,891 s.

| Hospital | Read | With a long clean window | Not read |
|---|---|---|---|
| A | 107 | 105 | 139 |
| B | 51 | 49 | 54 |
| D | 26 | 26 | 34 |
| E | 28 | 27 | 41 |
| F | 24 | 24 | 40 |
| All | 236 | 231 | 308 |

### Supplementary Figures

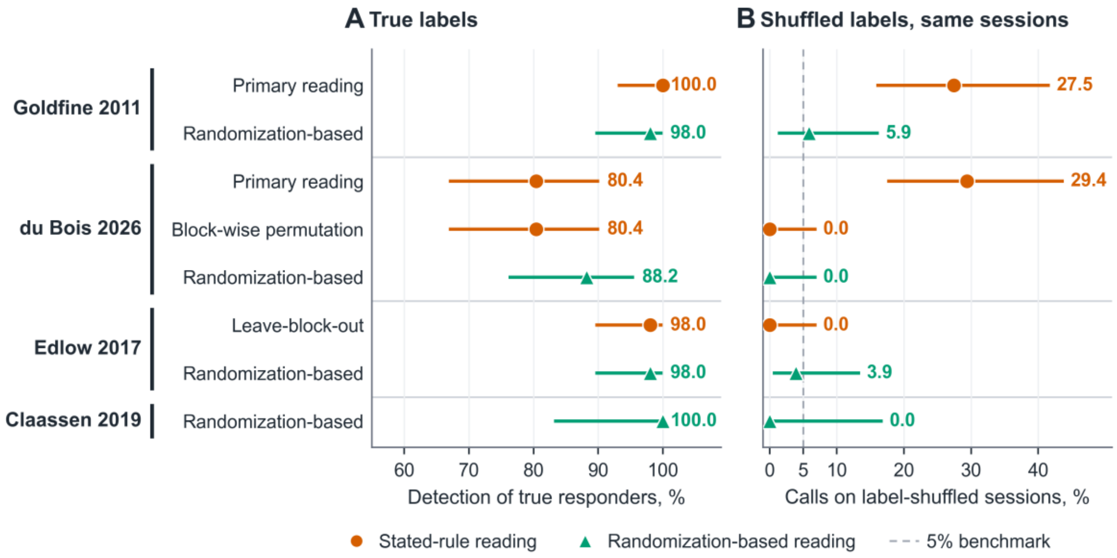


**Supplementary Figure S1. Detection and False Calls in Healthy Volunteers.** One row per reading, grouped by analysis. A, detection of true responders; B, calls on the same sessions with shuffled labels; exact 95% CIs, value printed to the right of each interval. Circles: stated-rule readings; triangles: randomization-based readings. Dashed line: 5% benchmark. Sessions were assembled from separate trial excerpts and contain no slow drift across blocks. This figure was Figure 3 of the earlier version.

#### Supplementary References

References to publications cited in the main text are given by author and year in square brackets and carry the numbers of the main-text reference list. References e1 to e24 are cited only in this supplement.


e1. Benjamini Y, Hochberg Y. Controlling the false discovery rate: a practical and powerful approach to multiple testing. *J R Stat Soc Series B Stat Methodol*. 1995;57(1):289-300. doi:10.1111/j.2517-6161.1995.tb02031.x

e2. Benjamini Y, Yekutieli D. The control of the false discovery rate in multiple testing under dependency. *Ann Stat*. 2001;29(4):1165-1188. doi:10.1214/aos/1013699998

e3. Thomson DJ. Spectrum estimation and harmonic analysis. *Proc IEEE*. 1982;70(9):1055-1096. doi:10.1109/PROC.1982.12433

e4. Bokil H, Andrews P, Kulkarni JE, Mehta S, Mitra PP. Chronux: a platform for analyzing neural signals. *J Neurosci Methods*. 2010;192(1):146-151. doi:10.1016/j.jneumeth.2010.06.020

e5. Hjorth B. An on-line transformation of EEG scalp potentials into orthogonal source derivations. *Electroencephalogr Clin Neurophysiol*. 1975;39(5):526-530. doi:10.1016/0013-4694(75)90056-5 e6. Ramoser H, Müller-Gerking J, Pfurtscheller G. Optimal spatial filtering of single trial EEG during imagined hand movement. *IEEE Trans Rehabil Eng*. 2000;8(4):441-446. doi:10.1109/86.895946

e7. Combrisson E, Jerbi K. Exceeding chance level by chance: the caveat of theoretical chance levels in brain signal classification and statistical assessment of decoding accuracy. *J Neurosci Methods*. 2015;250:126-136. doi:10.1016/j.jneumeth.2015.01.010

e8. Maris E, Oostenveld R. Nonparametric statistical testing of EEG- and MEG-data. *J Neurosci Methods*. 2007;164(1):177-190. doi:10.1016/j.jneumeth.2007.03.024

e9. Cohen J. A coefficient of agreement for nominal scales. *Educ Psychol Meas*. 1960;20(1):37-46. doi:10.1177/001316446002000104

e10. Holm S. A simple sequentially rejective multiple test procedure. *Scand J Stat*. 1979;6(2):65-70.

e11. Clopper CJ, Pearson ES. The use of confidence or fiducial limits illustrated in the case of the binomial. *Biometrika*. 1934;26(4):404-413. doi:10.1093/biomet/26.4.404

e12. Efron B. Bootstrap methods: another look at the jackknife. *Ann Stat*. 1979;7(1):1-26. doi:10.1214/aos/1176344552

e13. Armitage P. Tests for linear trends in proportions and frequencies. *Biometrics*. 1955;11(3):375-386. doi:10.2307/3001775 e14. Donoghue T, Haller M, Peterson EJ, et al. Parameterizing neural power spectra into periodic and aperiodic components. *Nat Neurosci*. 2020;23(12):1655-1665. doi:10.1038/s41593-020-00744-x

e15. Hodges JL, Lehmann EL. Estimates of location based on rank tests. *Ann Math Stat*. 1963;34(2):598-611. doi:10.1214/aoms/1177704172

e16. Theiler J, Eubank S, Longtin A, Galdrikian B, Farmer JD. Testing for nonlinearity in time series: the method of surrogate data. *Physica D*. 1992;58(1-4):77-94. doi:10.1016/0167-2789(92)90102-S

e17. Purdon PL, Weisskoff RM. Effect of temporal autocorrelation due to physiological noise and stimulus paradigm on voxel-level false-positive rates in fMRI. *Hum Brain Mapp*. 1998;6(4):239-249. doi:10.1002/(SICI)1097-0193(1998)6:4<239::AID-HBM4>3.0.CO;2-4

e18. Eklund A, Nichols TE, Knutsson H. Cluster failure: why fMRI inferences for spatial extent have inflated false-positive rates. *Proc Natl Acad Sci U S A*. 2016;113(28):7900-7905. doi:10.1073/pnas.1602413113

e19. MacKinnon JG, White H. Some heteroskedasticity-consistent covariance matrix estimators with improved finite sample properties. *J Econom*. 1985;29(3):305-325. doi:10.1016/0304-4076(85)90158-7

e20. Bossuyt PM, Reitsma JB, Bruns DE, et al. STARD 2015: an updated list of essential items for reporting diagnostic accuracy studies. *BMJ*. 2015;351:h5527. doi:10.1136/bmj.h5527 e21. von Elm E, Altman DG, Egger M, Pocock SJ, Gøtzsche PC, Vandenbroucke JP; STROBE Initiative. The Strengthening the Reporting of Observational Studies in Epidemiology (STROBE) statement: guidelines for reporting observational studies. *Lancet*. 2007;370(9596):1453-1457. doi:10.1016/S0140-6736(07)61602-X

e22. Westhall E, Rossetti AO, van Rootselaar AF, et al. Standardized EEG interpretation accurately predicts prognosis after cardiac arrest. *Neurology*. 2016;86(16):1482-1490. doi:10.1212/WNL.0000000000002462

e23. Mainsah B, Fleeting C, Balmat T, Sellers E, Collins L. bigP3BCI: an open, diverse and machine learning ready P300-based brain-computer interface dataset (version 1.0.0). PhysioNet. 2025. doi:10.13026/0byy-ry86

e24. Jamalabadi H, Alizadeh S, Schönauer M, Leibold C, Gais S. Classification based hypothesis testing in neuroscience: below-chance level classification rates and overlooked statistical properties of linear parametric classifiers. *Hum Brain Mapp*. 2016;37(5):1842-1855. doi:10.1002/hbm.23140
